# Validity and reliability of the Japanese version of the Movement Behaviour Questionnaire–Child (MBQ-C) for assessing active play, screen time, and sleep in preschool children

**DOI:** 10.64898/2026.09.28.26364096

**Authors:** Keisuke Komura, Mitsuya Yamakita, Akira Kyan, Shigeru Inoue, Mayumi Nagano, Rie Takenaga, Raoul Breugelmans, Eric Hajime Jego, Stewart G. Trost

## Abstract

**Background:** Valid and feasible measures of preschoolers’ 24-hour movement behaviours are needed for population-level research. We developed a Japanese version of the Movement Behaviour Questionnaire–Child (MBQ-C) and evaluated its measurement properties and practicality.

**Methods:** After translation and cross-cultural adaptation following the COSMIN procedure, including cognitive interviews, we evaluated the questionnaire in a cross-sectional validation study of Japanese children aged 3–6 years. Concurrent validity was assessed against hip-worn accelerometry and a 7-day diary; we also examined test–retest reliability, measurement error, agreement between response formats and modes, and practicality (completion time, burden, and clarity). Analyses used Spearman correlations, intraclass correlation coefficients, the SEM, smallest detectable change (SDC), and Wilcoxon signed-rank tests.

**Results:** Of 208 children, 170 provided valid accelerometer and diary data for the concurrent-validity analysis, and 187 completed the MBQ-C for the questionnaire-based analyses. Concurrent validity was weak for active and energetic play (Spearman ρ = 0.12–0.26), strong for total screen time (0.78–0.85), and moderate for total sleep (0.48–0.58). Test–retest reliability was moderate to good (ICC, 0.66–0.81), and within-child variability was lowest for sleep (CV 3%) and highest for the physical activity items (21–33%). The open- and closed-ended versions showed no significant differences; paper–electronic agreement was good for screen time and sleep but poor for active play. Completion took approximately 6–9 minutes, with low reported burden. Findings were robust in sensitivity analyses. At the group level, SDC for a mean was SDCind/√N: 119.8/√N, 74.0/√N, 112.6/√N, and 92.5/√N min/day for active play, energetic play, total screen time, and total sleep, respectively (N = number of children surveyed). The systematic differences between the two test–retest occasions were +15.7, +6.5, +4.0, and −5.4 min/day, respectively.

**Conclusions:** The Japanese MBQ-C is a practical option for assessing preschoolers’ active play, screen time, and sleep at the group level. By quantifying its group-level measurement error, this study provides benchmarks for interpreting group-level differences and change, supporting the use of the MBQ-C in surveillance and evaluation of programmes delivered at scale.

## BACKGROUND

Healthy growth and development in early childhood are associated with an appropriate balance among the 24-hour movement behaviours of physical activity, sedentary behaviour, and sleep [1]. On this basis, the World Health Organization has issued guidelines specifying how much time young children should spend on physical activity, sedentary behaviour (including screen time), and sleep across the 24-hour day [2].

As these guidelines have become more widely disseminated, the need has grown for feasible measures that can assess young children’s 24-hour movement behaviours validly and reliably [1]. Device-based methods such as accelerometry are objective but costly [3] and require specialised processing [4], limiting their use in large-scale surveys. Parent proxy-report questionnaires, in contrast, are inexpensive, suited to large-scale surveys, and can capture the type of behaviour, such as screen-time content [5]. Many such instruments exist, but a recent systematic review of proxy-report instruments for children aged 0–5 years concluded that none could comprehensively assess the 24-hour movement behaviours with established validity and reliability [6].

To address this gap, an Australian research group developed the Movement Behaviour Questionnaire (MBQ), a brief parent-report questionnaire for young children [7]. Its child version, the MBQ-C, is intended for children from the onset of independent walking to 5 years of age. It comprises nine items, imposes a low response burden, and yields indicators aligned with the guidelines. In the original validation against accelerometry and a 24-hour behaviour diary, the MBQ-C was reported to be a valid and reliable parent proxy measure of preschoolers’ movement behaviours, suitable for large-scale research [8]. A Chinese version was similarly supported [9]. However, the measurement properties of the MBQ-C have not yet been examined in Japanese preschool children. In addition, the original developers noted that additional measurement properties, such as responsiveness to change and the smallest detectable change (SDC), warrant examination in future MBQ research [8]. Evaluating these properties in a Japanese version can further clarify the measurement characteristics of the MBQ-C and support international comparison.

Therefore, this study aimed to develop a Japanese version of the MBQ-C and, as the primary aim, to evaluate its concurrent validity and test–retest reliability in Japanese preschool children. As secondary aims, we examined measurement error; agreement between response formats (open-ended and closed-ended versions) and between response modes (paper and electronic versions); and practicality.

## METHODS

### 2.1 Study Design

This study comprised two sequential phases: (1) translation and cross-cultural adaptation of the MBQ-C into Japanese, including cognitive interviews, conducted between July 2024 and April 2025; and (2) a cross-sectional validation study evaluating the measurement properties and practicality of the Japanese version, conducted in May–June and October–December 2025. Data collection was shared with a separate validation study of the Sasakawa Sports Foundation Preschoolers’ Activity Questionnaire (SSF-PAQ), completed by the same participants on the same occasions and reported separately [10]. The study was reported in accordance with the STROBE statement [11], and the completed checklist is provided in Additional file 1 (Supplementary Table S1). Both phases were approved by the Meijo University Human Research Ethics Committee (approval nos. 2024-17-2 and 2024-43), and informed consent was obtained from a parent or guardian of each child (online for the cognitive interviews in the translation phase, and in writing for the validation study).

### 2.2 Translation and Cross-Cultural Adaptation

#### 2.2.1 Translation Process

The MBQ-C was translated and cross-culturally adapted following the COSMIN procedure [12], in six steps (Fig. 1). In Step 1, two Japanese native speakers with high English proficiency (one non-specialist and one content specialist) independently produced forward translations. In Step 2, the specialist translator reconciled the two versions, and the original questionnaire developer clarified the intended meaning of the source items. In Step 3, two English native speakers with high Japanese proficiency and no content expertise independently back-translated the reconciled version, and the review team examined discrepancies in conceptual equivalence. In Step 4, the Japanese version was revised based on the developer’s review of the source text and back-translations. In Steps 5 and 6, the revised version underwent two rounds of pilot testing with cognitive interviews, each followed by revision, back translation, and developer review.

**Figure 1.**
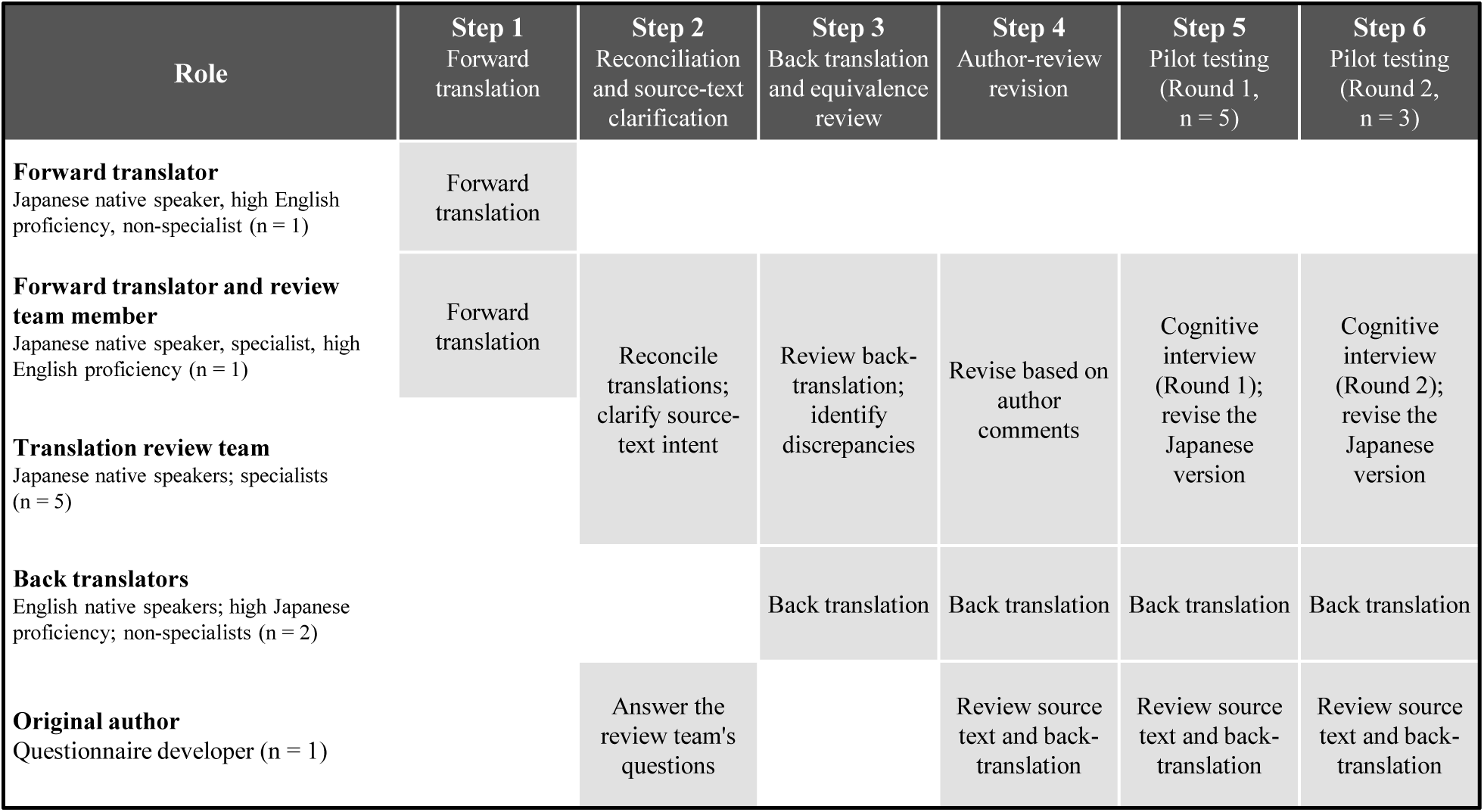
Translation process and roles. The Movement Behaviour Questionnaire-Child (MBQ-C) was translated into Japanese following the COSMIN translation procedure. Rows indicate the roles involved, and shaded cells indicate the activities performed by each role at each step. Detailed confirmations and revisions made during the process, together with the COSMIN checklist items addressed, are presented in Supplementary Table S10. Abbreviations: MBQ-C, Movement Behaviour Questionnaire-Child; COSMIN, COnsensus-based Standards for the selection of health Measurement INstruments.

#### 2.2.2 Cognitive Interviews

Cognitive interviews assessing comprehension and cultural appropriateness were conducted in two rounds with parents of children aged 3–6 years. Because the original development study noted an over-representation of highly educated respondents as a limitation [7], we sought socioeconomic diversity among the interviewees. Using a preliminary online form on educational attainment and household income, we selected interviewees with diverse socioeconomic backgrounds. Following the COSMIN Study Design checklist [12], which requires at least seven participants for a very good sample-size rating in qualitative content-validity studies, we aimed to recruit at least seven. Recruitment flyers were distributed through kindergartens and nursery centers to parents of 134 children aged 3–6 years; 9 provided consent and received 2,000 JPY (approximately 12.36 USD). Each interview was conducted one-on-one via Zoom (approximately 30 minutes) with screen sharing, based on a pre-developed interview guide (Supplementary Table S2). The interviews combined the think-aloud and verbal probing techniques [13]. In think-aloud, respondents verbalise their thoughts while answering; in verbal probing, the interviewer asks follow-up questions to confirm comprehension and identify difficulties.

Interviewers completed about three hours of prior training (a mock interview with feedback). Zoom audio was transcribed using AiNote (LINE WORKS Corp.) and corrected against the recordings. The transcripts were coded by two raters against a codebook grounded in the question-and-answer model of Willis and Tourangeau [13, 14], comprising nine categories (Supplementary Table S3). Inter-rater agreement was first assessed on one participant: Gwet’s AC1 [15] was 0.795, improving to 0.836 (almost perfect) after discussion and codebook refinement. The remaining data were then coded independently by the two raters, with discrepancies resolved by discussion. Findings from each round informed revisions, which were back-translated and reviewed by the developer before finalisation.

### 2.3 Validation Study

#### 2.3.1 Participants and Group Allocation

The required sample size was determined for the primary aim of evaluating concurrent validity. In the original MBQ-C validation study [8], Spearman correlations between the MBQ-C active play and energetic play items and accelerometer-derived total physical activity (Total PA) and moderate-to-vigorous physical activity (MVPA) ranged from 0.25 to 0.39. Assuming ρ = 0.30 (α = 0.05, power = 0.80; pwr package in R), 85 children were required per response format, giving 170 valid datasets for the open- and closed-ended versions. In a previous study of 3–6-year-olds using one week of 24-hour accelerometry plus a diary [16], approximately 79% of consenting participants provided valid accelerometer data and 27% of those given the recruitment documents through their center returned a consent form. Assuming 30% consent, distributing documents to 710 children would yield 213 participants; even with up to 20% missing data, this would provide 170 valid datasets.

Aiming for 213 consenting participants, we recruited through 11 conveniently sampled kindergartens, nursery centers, and centers for early childhood education and care (ECEC centers), attended by 705 children aged 3–6 years. However, one preschool withdrew shortly before the study, so recruitment documents were distributed through the remaining 10 preschools (623 children) to parents, and 208 consent forms were obtained (consent rate, 33%). An incentive of up to 3,000 JPY was provided upon completion (1,000 JPY each for accelerometer wear, the diary, and the questionnaire on both occasions). Participants were allocated to seven groups (Groups 1–7), with grades balanced across groups (Supplementary Table S4). Allocation prioritised concurrent validity while also enabling comparisons of test–retest reliability, open-versus closed-ended formats, and paper versus electronic modes.

#### 2.3.2 Study Protocol

Through the preschools, each family received a kit containing an accelerometer, a 7-day diary, the MBQ-C for Days 7 and 10, a QR code to register for automated reminders through a social-media messaging app (LINE, widely used in Japan) sending wear and diary reminders (daily at 07:00 and 20:00), and magnet clips for attaching the diary to the refrigerator. From Day 1 to Day 7, the child wore the accelerometer on the hip, including during sleep. Over the same period, the parent recorded the child’s sleep, screen time, accelerometer non-wear time, and arrival and departure times at the center in the diary each day, and completed the MBQ-C on Days 7 and 10. After the 10-day study, the kit was returned to the center and collected or shipped to the laboratory.

#### 2.3.3 Physical Activity Assessment Using Accelerometry

Physical activity was assessed with a triaxial accelerometer (ActiGraph wGT3X-BT). Children were asked to wear the accelerometer on the right hip with an elastic belt for seven consecutive days, including during sleep, removing it only for bathing and water-based activities. The device was initialised at 100 Hz with idle sleep mode enabled and local time, recording from 00:00 on Day 1. After return, the raw data were exported in .gt3x format using ActiLife v6.13.6.

For young children, raw-data machine-learning methods classify physical activity more accurately than conventional counts-based methods [17, 18]. We therefore used the Preschool Hip Random Forest Free-Living Lag–Lead classifier [17] in the R package actimetric [19], which estimates physical activity and sleep from raw data. However, a processing issue affecting 100-Hz data starting at 00:00 was identified; after consulting the classifier developer (a co-author, S.G.T.), we processed the .gt3x files directly with the underlying R script (since corrected in actimetric).

The classifier categorises physical activity in 15-second epochs into five categories: sedentary (SED), light activity and games (L_ACT_G), moderate-to-vigorous activity and games (MV_ACT_G), walking, and running [17]. Non-wear time was determined separately using the SD_VM algorithm of Ahmadi et al. [20]: (1) windows where the vector-magnitude standard deviation was below 13 mg for at least 30 minutes were classed as non-wear, and (2) wear periods shorter than 30 minutes and under 30% of the surrounding non-wear length were reclassified as non-wear. actimetric also provides a sleep category, but its estimation uses a wrist-worn algorithm based on z-axis angle variation [21, 22].

Because the present data were hip-worn, sleep was instead determined using an algorithm developed for hip data in this age group [23], combined with a sleep diary (Section 2.3.5).

Total PA was calculated as the sum of the time spent in L_ACT_G, MV_ACT_G, walking, and running, and MVPA as the sum of the time spent in MV_ACT_G, walking, and running. As an additional comparison metric for the play items, mean Euclidean Norm Minus One (ENMO; mg) [24] was calculated from raw acceleration over the same valid wear epochs (07:00–21:00), excluding non-wear. Daily means were averaged separately for weekdays and weekends/holidays and combined using 5:2 weighting; associations with the Day-7 play items were assessed using Spearman correlations.

#### 2.3.4 Screen Time and Sleep Diary

Diary information on screen time and sleep was collected with a 24-hour behaviour diary kept by the parent over seven days. This diary was based on the diary used in the original MBQ-C validation study [8], which was adapted from the screen-time diary used by Mendoza et al. [25] (Supplementary Material S5).

Parents were asked to record, in 15-minute units, the child’s sleep, passive screen time, interactive screen time, and accelerometer non-wear time. For passive and interactive screen time, the time spent in a standing position was also recorded. In addition, for each day, the parent recorded whether the child had spent that day in the usual way for that day of the week and rated how accurately the diary had been completed (Supplementary Material S5).

When entering the diary, an arrow recorded in the middle of a 15-minute cell was assigned to the nearer 15-minute time point; when positioned exactly midway, it was assigned to the time point that lengthened the corresponding behaviour. Total screen time was calculated as the union of passive sitting screen time, passive standing screen time, interactive sitting screen time, and interactive standing screen time; overlapping or continuous periods were merged to avoid double counting. Sitting screen time was calculated as the sum of passive sitting screen time and interactive sitting screen time.

#### 2.3.5 Sleep Assessment Integrating Diary and Accelerometer Data

Night-sleep episodes separated by an awake gap under 30 minutes were treated as one continuous period. When both diary endpoints were recorded, they defined the diary-based night-sleep interval; when one was missing, it was imputed from the mean time for the same child within the same next-day category. The next-day category classified each night according to whether the following day was a weekday or a weekend/holiday, because night sleep extends into the next morning. Because times cross midnight, a circular mean was used (or the single available time when only one day applied). If an endpoint remained missing after imputation, no interval was created for that day.

Accelerometer-derived bed-rest endpoints were identified using the R package PhysActBedRest, which implements the sleep-detection algorithm for hip-worn accelerometers in young children developed by Tracy et al. [23]. Before the Tracy determination, non-wear epochs identified by the criteria of Ahmadi et al. [20] were excluded.

For each diary-based interval, onset and offset were corrected separately. For night sleep, the search range was ±30 minutes of the diary endpoint, based on Werner et al.[26], who reported diary– accelerometer differences of about 28 minutes for onset and 24 for offset. If a same-type Tracy endpoint occurred within ±30 minutes, it was adopted. Otherwise, the local wear ratio within ±30 minutes was calculated, and only when ≥90% was the range expanded to ±90 minutes to prioritise the accelerometer endpoint, adopting a same-type Tracy endpoint if detected. When none was found within ±90 minutes, or the wear ratio was <90%, the diary endpoint was retained. For day sleep, the nearest same-type Tracy endpoint was adopted only when detected within ±15 minutes of the diary endpoint; otherwise, the diary endpoint was retained.

The 15-second-epoch classification was then updated: epochs within a corrected sleep interval were classed as sleep, and machine-learning sleep epochs outside these intervals were reclassified as sedentary. Daily sleep duration was calculated from these corrected sleep intervals. The full processing workflow and worked examples of the interval correction and epoch-level reclassification are provided in Supplementary Figure S6.

#### 2.3.6 Practicality Indicators

Three practicality indicators were added to the MBQ-C: completion time (respondents recorded the time at the start and end of the items), perceived response burden (four-point scale), and perceived clarity (five-point scale).

#### 2.3.7 Data Cleaning and Processing

A valid accelerometer day required at least 600 minutes of wear between 07:00 and 21:00, set as the main waking period from a national survey of Japanese preschoolers (mean wake time 06:56–07:28 and bedtime 21:12–21:24 across weekdays and weekends) [27]. The main analysis included participants with at least three valid weekdays and at least one valid weekend/holiday. Including multiple weekdays plus at least one weekend day follows previous studies for reliably estimating habitual physical activity [28, 29], and the ≥600-minutes/day criterion reflects the 10-hours/day standard common in accelerometer studies of young children [30].

We excluded participants who returned the kit unused (n = 16), those with missing data due to device failure (n = 2), and those not meeting the valid-day criteria (n = 15). We also noticed a few records with implausibly high MVPA relative to step counts. The Mahalanobis distance was calculated using mean steps and mean MVPA, and participants above the 99.9th percentile of the χ² distribution (df = 2) were excluded as potentially invalid (n = 4; Supplementary Figure S7).

For accelerometer variables, diary screen time, and corrected sleep, daily values were summarised as separate weekday and weekend/holiday means, with a weighted weekly average of (weekday × 5 + weekend/holiday × 2)/7. Following the original scoring, MBQ-C active play and screen time (with weekday and weekend items) were similarly weighted; total sleep was the sum of typical night and day sleep.

Open-ended responses were converted to minutes per day from the reported hours and minutes; closed-ended responses used the midpoint of the selected category. Extreme values were truncated using the thresholds reported in the original study [8]: 480 (total active play), 360 (energetic play), 600 (passive and interactive screen time), and 360 (daytime sleep) min/day. Values exceeding these thresholds were set to the corresponding upper limits rather than treated as invalid.

#### 2.3.8 Statistical Analysis

The main validity analysis included participants with valid accelerometer data who completed the MBQ-C on Day 7. Each analysis used complete cases, and the number of participants is reported for each. Participant characteristics were summarised as means and standard deviations for continuous variables and as frequencies and percentages for categorical variables.

Concurrent validity was evaluated using Spearman rank correlation coefficients (ρ) between the Day-7 MBQ-C values and the corresponding criterion measures. The 95% confidence intervals (CIs) were calculated using Fisher’s z transformation as implemented in the DescTools package [31]. For descriptive interpretation, ρ was classified as negligible (< 0.10), weak (0.10–0.39), moderate (0.40– 0.69), strong (0.70–0.89), or very strong (≥ 0.90) [32]. Systematic differences from the criterion were examined with the Wilcoxon signed-rank test, except for active and energetic play, because parent-reported play time and accelerometer-based movement intensity are not measured on the same scale, making a systematic difference uninformative.

Test–retest reliability (Day 7 vs Day 10, same version on both occasions) was assessed using intraclass correlation coefficients (ICCs) and 95% CIs from a single-measurement, absolute-agreement, two-way mixed-effects model (’irr’ package [33]). ICCs were interpreted as poor (<0.5), moderate (0.5–0.75), good (0.75–0.90), and excellent (>0.90) [34]. Agreement between response formats (open- vs closed-ended) and between modes (paper vs electronic) was assessed using the Spearman ρ, the ICC, and the paired Wilcoxon signed-rank test for systematic differences.

Measurement error was characterised by the within-child coefficient of variation, the standard error of measurement (SEM), and the SDC (also termed the minimal detectable change at the 95% level, MDC95). SEM was estimated from a linear mixed-effects model with random effects for participant and measurement occasion, as the square root of the sum of the measurement-occasion and residual variance components. The individual-level SDC (SDCind) was calculated as 1.96 × √2 × SEM [35].

Because random measurement error partly averages out when a group mean is analysed, the SDC for a group mean is smaller than for an individual by a factor of √N; we therefore also report the group-level SDC, SDCgroup = SDCind/√N, where N is the number of children surveyed [36]. Agreement between the MBQ-C and the criterion measures for total screen time and total sleep was further examined using Bland–Altman analysis (bias, 95% limits of agreement, and proportional bias). Practicality indicators were summarised descriptively.

We conducted three sensitivity analyses. First, because many international studies define preschoolers as aged 3–5 years, we re-examined estimates after restricting the sample to ages 3–5 years (<72 months). Second, we varied the valid-day wear-time criterion: a stricter definition (≥960 minutes/day over 24 hours) and a less strict one (≥360 minutes/day, 07:00–21:00), because at least 6 hours of wear on any 3 days has been reported to give acceptable reliability in young children [37]. Third, we recomputed each child’s weighted weekly criterion means from three day subsets: (i) days reported as usual, (ii) days rated as accurately recorded, and (iii) both. These analyses were motivated by the MBQ-C asking respondents about a typical day, whereas the measurement week may include atypical days, and by the dependence of diary-derived measures on recording quality.

All analyses were performed in R (version 4.5.0).

## RESULTS

### 3.1 Translation and Cross-Cultural Adaptation

Of the parents who provided consent, one could not be contacted, and eight completed the cognitive interviews in Steps 5 and 6 (Supplementary Figure S8); their characteristics are shown in Supplementary Table S9. Among the eight interviewees, four (50%) were university graduates and seven (88%) reported a household income below the national average for households with children, 8.205 million JPY [38]. The key revisions made through this process, which included clarifying the target child for parent report, adjusting the wording used to ask about duration, rendering “movement behaviours” with an age-appropriate Japanese expression, and visually emphasising the “typical day” framing, are summarised in Supplementary Table S10, and the finalised open-ended and closed-ended Japanese versions are provided in Supplementary Material S11 and S12.

### 3.2 Validation Study

#### 3.2.1 Participants

Of the 208 children whose parents provided consent, 187 completed the MBQ-C; the remaining 21 did not return a questionnaire, most undergoing no measurement (participant flow, Supplementary Figure S13). In the accelerometer-based stream, 171 children had valid accelerometer data after exclusions for non-measurement, device failure, insufficient valid days, and MVPA outliers, and 170 of these also completed the MBQ-C on Day 7 and were included in the main concurrent-validity analysis. Their characteristics are summarised in Table 1 (mean age, 4.95 years; 53% girls; mother as respondent in 89%). The questionnaire-based analyses (reliability, measurement error, format and mode agreement, practicality) do not require accelerometer data; each used only the respondents with the required data, so the number analysed varied and is reported for each (characteristics in Supplementary Table S14).

**Table 1.**
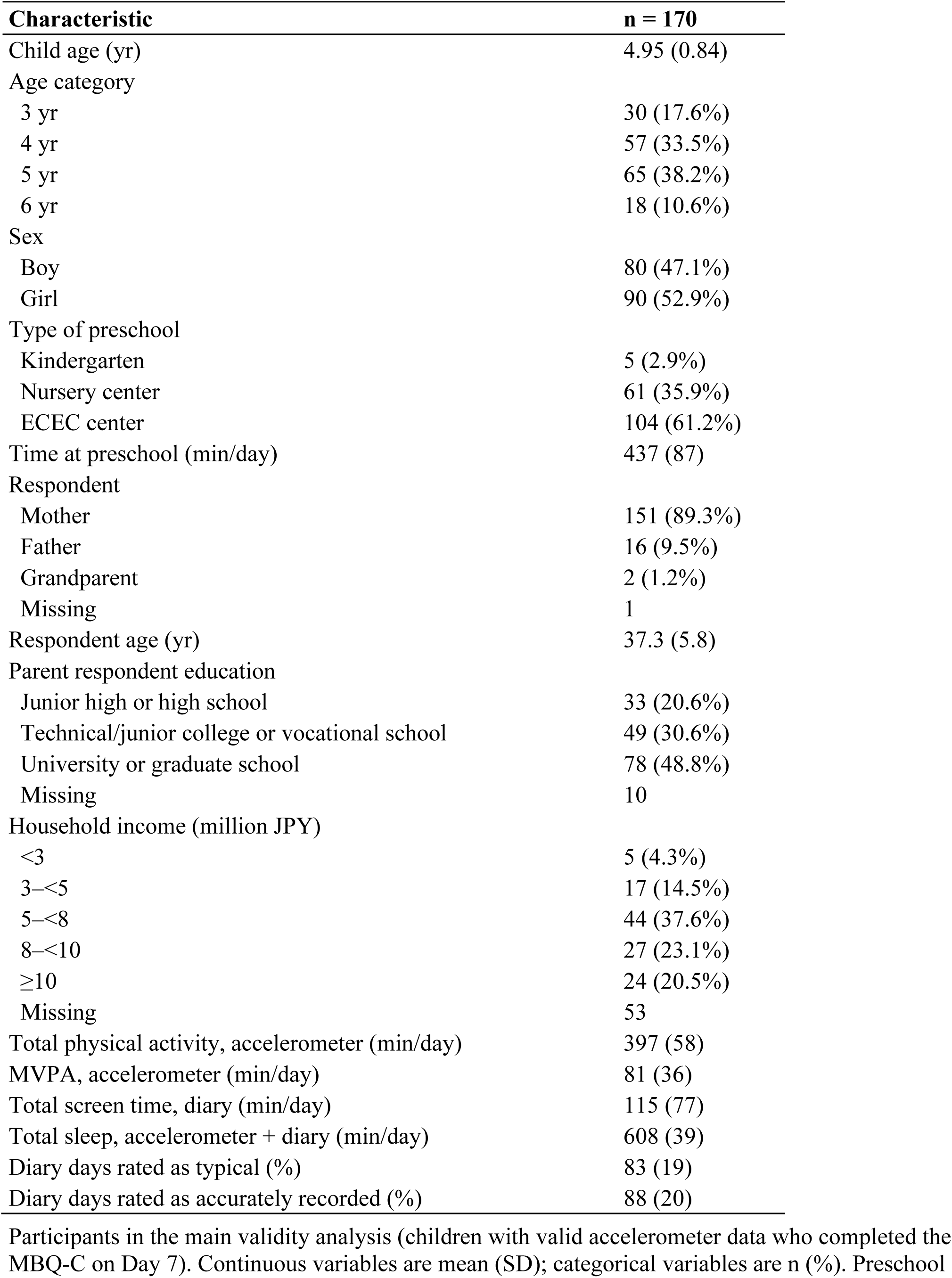
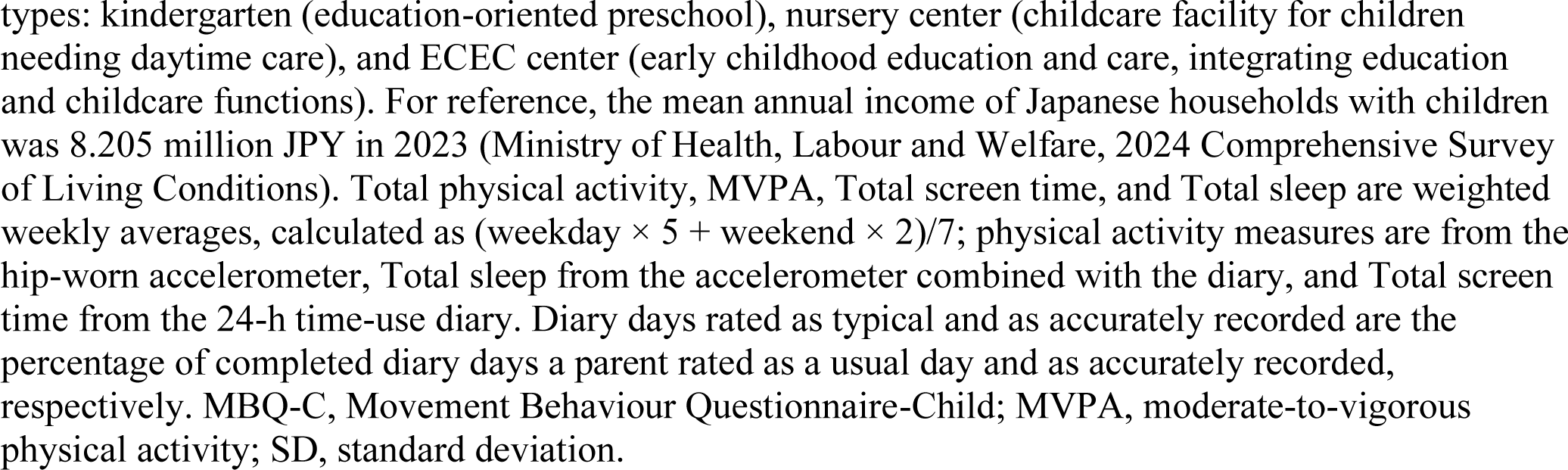
Participant characteristics.

| **Characteristic** | **n = 170** |
|---|---|
| Child age (yr) | 4.95 (0.84) |
| Age category |  |
| 3 yr | 30 (17.6%) |
| 4 yr | 57 (33.5%) |
| 5 yr | 65 (38.2%) |
| 6 yr | 18 (10.6%) |
| Sex |  |
| Boy | 80 (47.1%) |
| Girl | 90 (52.9%) |
| Type of preschool |  |
| Kindergarten | 5 (2.9%) |
| Nursery center | 61 (35.9%) |
| ECEC center | 104 (61.2%) |
| Time at preschool (min/day) | 437 (87) |
| Respondent |  |
| Mother | 151 (89.3%) |
| Father | 16 (9.5%) |
| Grandparent | 2 (1.2%) |
| Missing | 1 |
| Respondent age (yr) | 37.3 (5.8) |
| Parent respondent education |  |
| Junior high or high school | 33 (20.6%) |
| Technical/junior college or vocational school | 49 (30.6%) |
| University or graduate school | 78 (48.8%) |
| Missing | 10 |
| Household income (million JPY) |  |
| <3 | 5 (4.3%) |
| 3–<5 | 17 (14.5%) |
| 5–<8 | 44 (37.6%) |
| 8–<10 | 27 (23.1%) |
| ≥10 | 24 (20.5%) |
| Missing | 53 |
| Total physical activity, accelerometer (min/day) | 397 (58) |
| MVPA, accelerometer (min/day) | 81 (36) |
| Total screen time, diary (min/day) | 115 (77) |
| Total sleep, accelerometer + diary (min/day) | 608 (39) |
| Diary days rated as typical (%) | 83 (19) |
| Diary days rated as accurately recorded (%) | 88 (20) |

Participants in the main validity analysis (children with valid accelerometer data who completed the MBQ-C on Day 7). Continuous variables are mean (SD); categorical variables are n (%). Preschool types: kindergarten (education-oriented preschool), nursery center (childcare facility for children needing daytime care), and ECEC center (early childhood education and care, integrating education and childcare functions). For reference, the mean annual income of Japanese households with children was 8.205 million JPY in 2023 (Ministry of Health, Labour and Welfare, 2024 Comprehensive Survey of Living Conditions). Total physical activity, MVPA, Total screen time, and Total sleep are weighted weekly averages, calculated as (weekday × 5 + weekend × 2)/7; physical activity measures are from the hip-worn accelerometer, Total sleep from the accelerometer combined with the diary, and Total screen time from the 24-h time-use diary. Diary days rated as typical and as accurately recorded are the percentage of completed diary days a parent rated as a usual day and as accurately recorded, respectively. MBQ-C, Movement Behaviour Questionnaire-Child; MVPA, moderate-to-vigorous physical activity; SD, standard deviation.

#### 3.2.2 Concurrent Validity

Concurrent validity differed across behaviours (Table 2). Validity was weak for active play (open-ended ρ = 0.15 [95% CI, –0.08 to 0.36]; closed-ended ρ = 0.26 [0.06–0.44]) and energetic play (0.22 [0.00–0.43] and 0.12 [–0.09 to 0.32]). In contrast, total screen time showed strong validity (0.85 [0.77– 0.90] and 0.78 [0.69–0.85]), and total sleep showed moderate validity (0.58 [0.40–0.71] and 0.48 [0.31–0.63]). This pattern was consistent across the paper and electronic modes. Detailed results by indicator and day type are reported in Supplementary Table S15. Sleep showed lower validity on weekends than on weekdays, most clearly for night sleep, which declined from a moderate weekday range (ρ = 0.61–0.64) to a weak weekend range (0.20–0.28); differences for the other behaviours were smaller or less consistent.

**Table 2.**
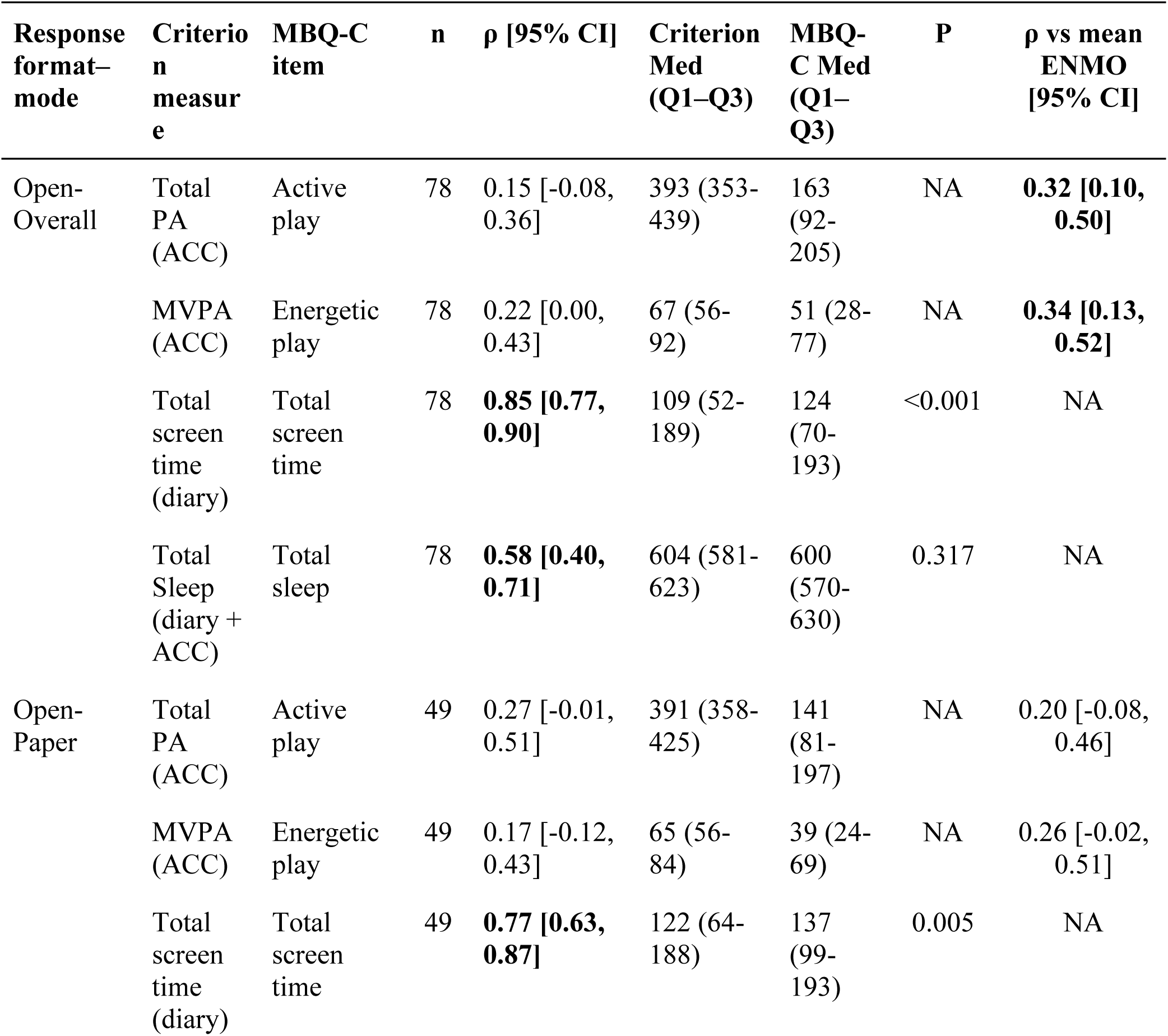
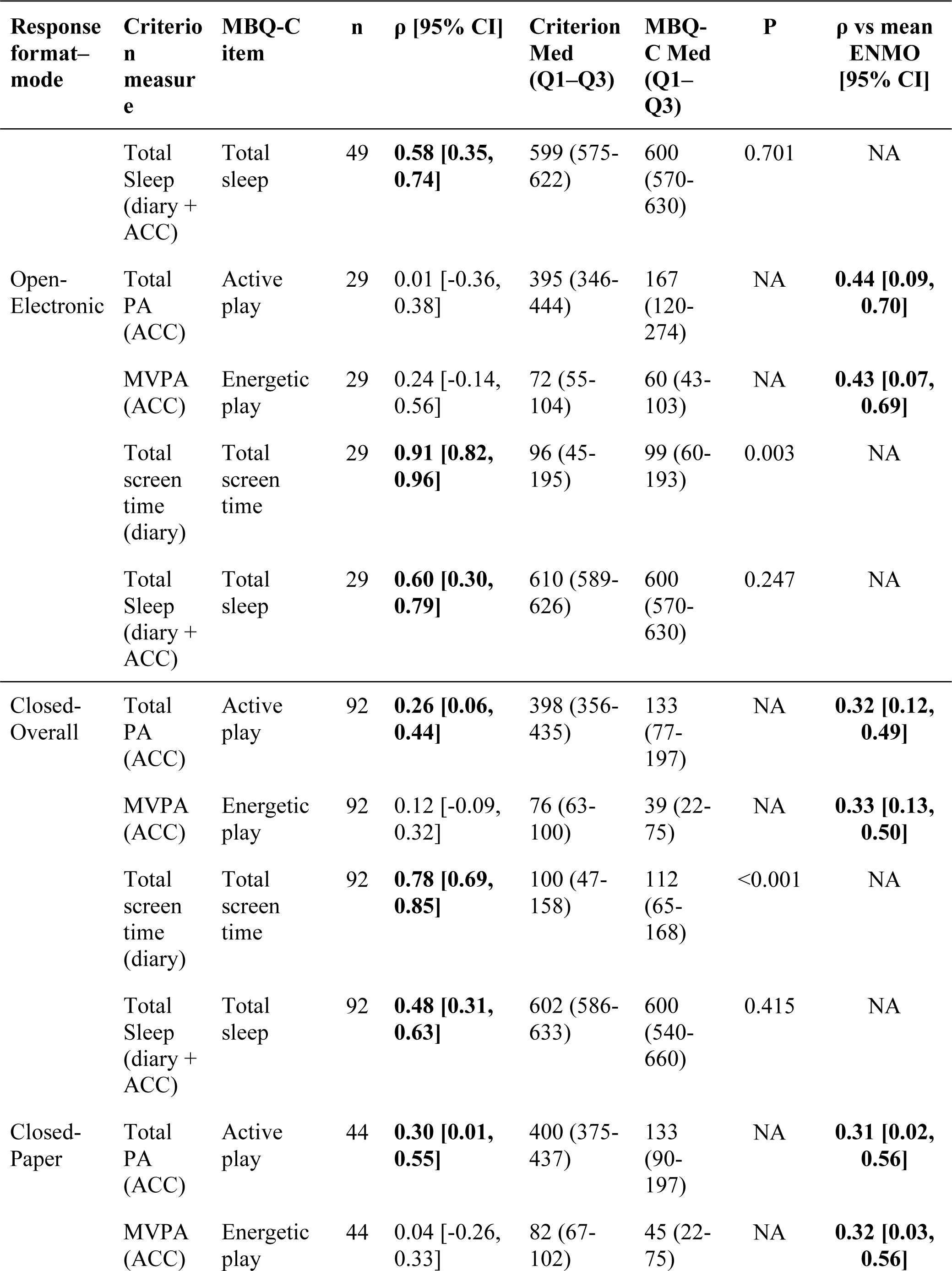
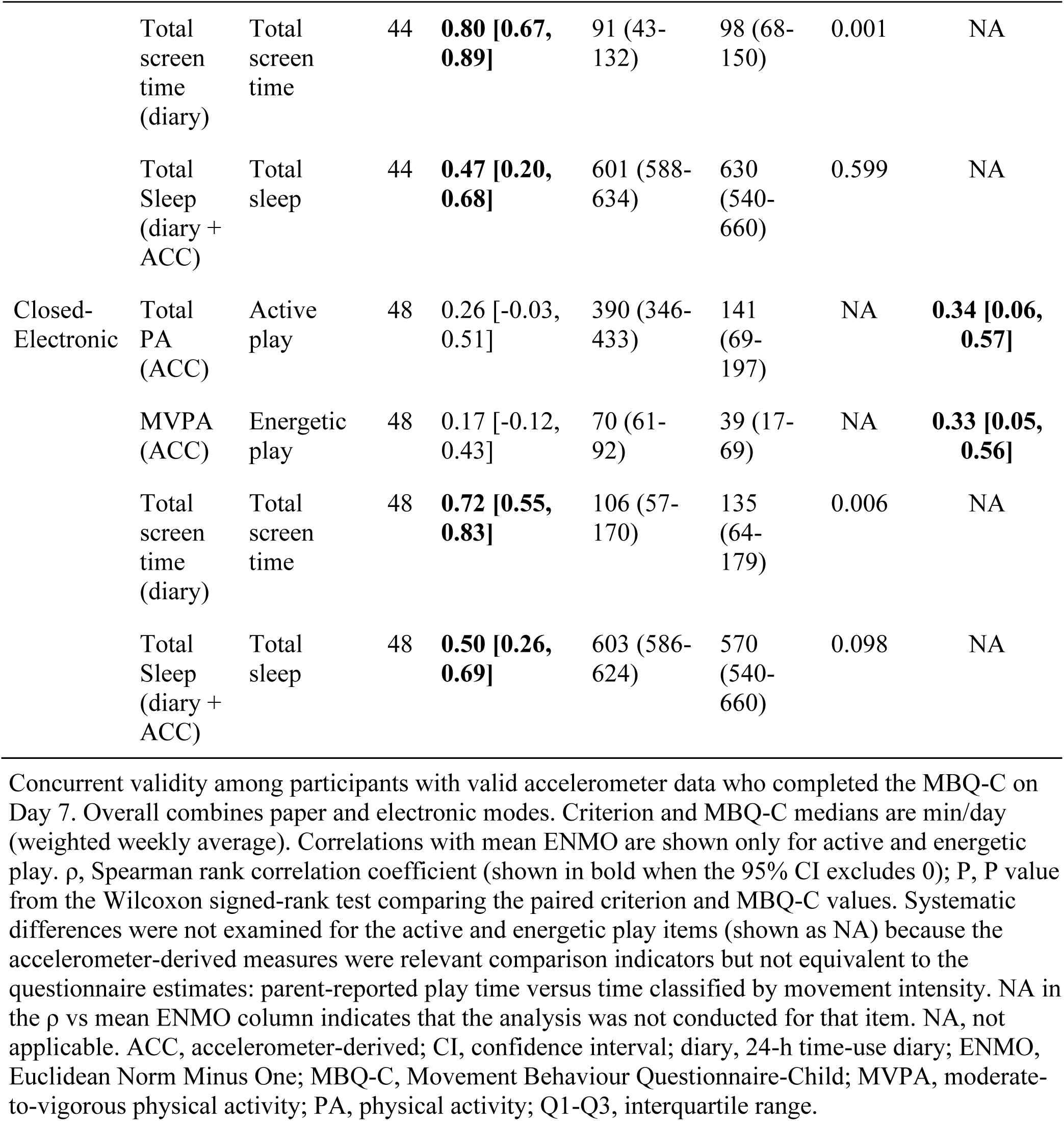
Validity of the Japanese MBQ-C by response format and mode.

| **Response format–mode** | **Criterion measure** | **MBQ-C item** | **n** | **ρ [95% CI]** | **Criterion Med (Q1–Q3)** | **MBQ-C Med (Q1–Q3)** | **P** | **ρ vs mean ENMO [95% CI]** |
|---|---|---|---|---|---|---|---|---|
| Open-Overall | Total PA (ACC) | Active play | 78 | 0.15 [-0.08, 0.36] | 393 (353-439) | 163 (92-205) | NA | **0.32 [0.10, 0.50]** |
|  | MVPA (ACC) | Energetic play | 78 | 0.22 [0.00, 0.43] | 67 (56-92) | 51 (28-77) | NA | **0.34 [0.13, 0.52]** |
|  | Total screen time (diary) | Total screen time | 78 | **0.85 [0.77, 0.90]** | 109 (52-189) | 124 (70-193) | <0.001 | NA |
|  | Total Sleep (diary + ACC) | Total sleep | 78 | **0.58 [0.40, 0.71]** | 604 (581-623) | 600 (570-630) | 0.317 | NA |
| Open-Paper | Total PA (ACC) | Active play | 49 | 0.27 [-0.01, 0.51] | 391 (358-425) | 141 (81-197) | NA | 0.20 [-0.08, 0.46] |
|  | MVPA (ACC) | Energetic play | 49 | 0.17 [-0.12, 0.43] | 65 (56-84) | 39 (24-69) | NA | 0.26 [-0.02, 0.51] |
|  | Total screen time (diary) | Total screen time | 49 | **0.77 [0.63, 0.87]** | 122 (64-188) | 137 (99-193) | 0.005 | NA |
| **Response format–mode** | **Criterion measure** | **MBQ-C item** | **n** | **ρ [95% CI]** | **Criterion Med (Q1–Q3)** | **MBQ-C Med (Q1–Q3)** | **P** | **ρ vs mean ENMO [95% CI]** |
|  | Total Sleep (diary + ACC) | Total sleep | 49 | **0.58 [0.35, 0.74]** | 599 (575-622) | 600 (570-630) | 0.701 | NA |
| Open-Electronic | Total PA (ACC) | Active play | 29 | 0.01 [-0.36, 0.38] | 395 (346-444) | 167 (120-274) | NA | **0.44 [0.09, 0.70]** |
|  | MVPA (ACC) | Energetic play | 29 | 0.24 [-0.14, 0.56] | 72 (55-104) | 60 (43-103) | NA | **0.43 [0.07, 0.69]** |
|  | Total screen time (diary) | Total screen time | 29 | **0.91 [0.82, 0.96]** | 96 (45-195) | 99 (60-193) | 0.003 | NA |
|  | Total Sleep (diary + ACC) | Total sleep | 29 | **0.60 [0.30, 0.79]** | 610 (589-626) | 600 (570-630) | 0.247 | NA |
| Closed-Overall | Total PA (ACC) | Active play | 92 | **0.26 [0.06, 0.44]** | 398 (356-435) | 133 (77-197) | NA | **0.32 [0.12, 0.49]** |
|  | MVPA (ACC) | Energetic play | 92 | 0.12 [-0.09, 0.32] | 76 (63-100) | 39 (22-75) | NA | **0.33 [0.13, 0.50]** |
|  | Total screen time (diary) | Total screen time | 92 | **0.78 [0.69, 0.85]** | 100 (47-158) | 112 (65-168) | <0.001 | NA |
|  | Total Sleep (diary + ACC) | Total sleep | 92 | **0.48 [0.31, 0.63]** | 602 (586-633) | 600 (540-660) | 0.415 | NA |
| Closed-Paper | Total PA (ACC) | Active play | 44 | **0.30 [0.01, 0.55]** | 400 (375-437) | 133 (90-197) | NA | **0.31 [0.02, 0.56]** |
|  | MVPA (ACC) | Energetic play | 44 | 0.04 [-0.26, 0.33] | 82 (67-102) | 45 (22-75) | NA | **0.32 [0.03, 0.56]** |
|  | Total screen time (diary) | Total screen time | 44 | **0.80 [0.67, 0.89]** | 91 (43-132) | 98 (68-150) | 0.001 | NA |
|  | Total Sleep (diary + ACC) | Total sleep | 44 | **0.47 [0.20, 0.68]** | 601 (588-634) | 630 (540-660) | 0.599 | NA |
| Closed-Electronic | Total PA (ACC) | Active play | 48 | 0.26 [-0.03, 0.51] | 390 (346-433) | 141 (69-197) | NA | **0.34 [0.06, 0.57]** |
|  | MVPA (ACC) | Energetic play | 48 | 0.17 [-0.12, 0.43] | 70 (61-92) | 39 (17-69) | NA | **0.33 [0.05, 0.56]** |
|  | Total screen time (diary) | Total screen time | 48 | **0.72 [0.55, 0.83]** | 106 (57-170) | 135 (64-179) | 0.006 | NA |
|  | Total Sleep (diary + ACC) | Total sleep | 48 | **0.50 [0.26, 0.69]** | 603 (586-624) | 570 (540-660) | 0.098 | NA |

Concurrent validity among participants with valid accelerometer data who completed the MBQ-C on Day 7. Overall combines paper and electronic modes. Criterion and MBQ-C medians are min/day (weighted weekly average). Correlations with mean ENMO are shown only for active and energetic play. ρ, Spearman rank correlation coefficient (shown in bold when the 95% CI excludes 0); P, P value from the Wilcoxon signed-rank test comparing the paired criterion and MBQ-C values. Systematic differences were not examined for the active and energetic play items (shown as NA) because the accelerometer-derived measures were relevant comparison indicators but not equivalent to the questionnaire estimates: parent-reported play time versus time classified by movement intensity. NA in the ρ vs mean ENMO column indicates that the analysis was not conducted for that item. NA, not applicable. ACC, accelerometer-derived; CI, confidence interval; diary, 24-h time-use diary; ENMO, Euclidean Norm Minus One; MBQ-C, Movement Behaviour Questionnaire-Child; MVPA, moderate-to-vigorous physical activity; PA, physical activity; Q1-Q3, interquartile range.

#### 3.2.3 Test–retest Reliability

Test–retest reliability was moderate to good for the overall open- and closed-ended versions (ICC, 0.66–0.81; Table 3). By behaviour, ICCs were 0.71–0.79 for active play, 0.66–0.69 for energetic play, 0.79–0.81 for total screen time, and 0.68–0.72 for total sleep.

**Table 3.** Test-retest reliability of the Japanese MBQ-C by response format and mode.

| **Response format–mode** | **MBQ-C item** | **n** | **ICC [95% CI]** | **Day 7 Med (Q1–Q3)** | **Day 10 Med (Q1–Q3)** |
|---|---|---|---|---|---|
| Open-Overall | Active play | 59 | **0.79 [0.67, 0.87]** | 171 (105-229) | 146 (86-236) |
|  | Energetic play | 59 | **0.66 [0.49, 0.78]** | 57 (29-81) | 56 (26-101) |
|  | Total screen time | 59 | **0.79 [0.67, 0.87]** | 137 (74-208) | 141 (69-219) |
|  | Total sleep | 59 | **0.68 [0.51, 0.79]** | 600 (560-630) | 600 (570-630) |
|  | Sleep routine | 59 | **0.81 [0.70, 0.88]** | 6 (4-7) | 6 (4-7) |
| Open-Paper | Active play | 29 | **0.76 [0.54, 0.88]** | 159 (77-206) | 129 (63-206) |
|  | Energetic play | 29 | 0.47 [0.12, 0.71] | 51 (24-69) | 39 (17-77) |
|  | Total screen time | 29 | **0.82 [0.65, 0.91]** | 169 (137-214) | 161 (119-219) |
|  | Total sleep | 29 | **0.74 [0.52, 0.87]** | 600 (570-650) | 600 (570-630) |
|  | Sleep routine | 29 | **0.77 [0.57, 0.89]** | 6 (3-7) | 6 (3-7) |
| Open-Electronic | Active play | 30 | **0.81 [0.64, 0.91]** | 176 (121-274) | 161 (121-240) |
|  | Energetic play | 30 | **0.75 [0.54, 0.87]** | 60 (43-98) | 61 (32-103) |
|  | Total screen time | 30 | **0.76 [0.55, 0.88]** | 90 (57-175) | 87 (47-212) |
|  | Total sleep | 30 | **0.63 [0.35, 0.80]** | 600 (548-630) | 600 (570-630) |
|  | Sleep routine | 30 | **0.87 [0.75, 0.94]** | 7 (4-7) | 6.5 (5-7) |
| Closed-Overall | Active play | 61 | **0.71 [0.53, 0.82]** | 133 (64-197) | 90 (58-150) |
|  | Energetic play | 61 | **0.69 [0.49, 0.82]** | 45 (22-75) | 29 (18-54) |
|  | Total screen time | 61 | **0.81 [0.70, 0.88]** | 133 (75-176) | 118 (69-166) |
|  | Total sleep | 61 | **0.72 [0.58, 0.82]** | 630 (540-660) | 630 (540-660) |
|  | Sleep routine | 61 | **0.83 [0.73, 0.90]** | 4 (2-4) | 3 (2-4) |
| Closed-Paper | Active play | 30 | **0.63 [0.32, 0.81]** | 141 (90-197) | 107 (48-146) |
|  | Energetic play | 30 | **0.58 [0.26, 0.78]** | 52 (24-80) | 42 (19-60) |
|  | Total screen time | 30 | **0.78 [0.59, 0.89]** | 114 (87-163) | 137 (84-160) |
|  | Total sleep | 30 | **0.76 [0.56, 0.88]** | 630 (540-660) | 630 (540-660) |
|  | Sleep routine | 30 | **0.86 [0.73, 0.93]** | 4 (2.2-4) | 3.5 (1-4) |
| Closed-Electronic | Active play | 31 | **0.79 [0.61, 0.89]** | 133 (55-195) | 90 (58-163) |
|  | Energetic play | 31 | **0.80 [0.60, 0.90]** | 32 (10-63) | 22 (13-39) |
|  | Total screen time | 31 | **0.85 [0.67, 0.93]** | 141 (70-182) | 112 (58-166) |
|  | Total sleep | 31 | **0.67 [0.42, 0.83]** | 570 (540-660) | 630 (540-660) |
|  | Sleep routine | 31 | **0.80 [0.62, 0.90]** | 3 (2-4) | 3 (2-4) |

Test-retest reliability among participants who completed the same MBQ-C version and mode on Day 7 and Day 10. Overall pools paper and electronic modes within each format. Values are min/day (weighted weekly average), except Sleep routine. ICC, intraclass correlation coefficient, estimated with a single-measurement, absolute-agreement, two-way mixed-effects model; values ≥0.50 (moderate or higher) are shown in bold. Sleep routine: open = nights/week (0-7); closed = 5 categories (0 = never, 1 = 1-2, 2 = 3-4, 3 = 5-6, 4 = every night); medians are not comparable across formats. CI, confidence interval; MBQ-C, Movement Behaviour Questionnaire-Child; Q1-Q3, interquartile range.

#### 3.2.4 Measurement Error

Within-child measurement error was lowest for sleep, intermediate for screen time, and highest for the play items, with CVs of 2.9%, 17.3%, and 21.3–33.2%, respectively; the SDCind ranged from 74.0 (energetic play) to 119.8 min/day (active play) (Table 4). At the group level, SDCgroup = SDCind/√N; for example, in a survey of 1000 children this corresponds to about 3.8, 2.3, 3.6, and 2.9 min/day for active play, energetic play, total screen time, and total sleep, respectively (Table 4). The mean difference between occasions (Day 7 to Day 10) was +15.7, +6.5, +4.0, and −5.4 min/day, respectively (Table 4).

**Table 4.** Measurement error in repeated assessments of the Japanese version of the MBQ-C.

| **Response format** | **MBQ-C item** | **n** | **Day 7 Mean (SD)** | **Day 10 Mean (SD)** | **Difference Mean (SD)** | **CV (%)** | **SEM** | **SDCind** | **SDCgroup** |
|---|---|---|---|---|---|---|---|---|---|
| Overall (all versions) | Active play | 120 | 155.2 (91.8) | 139.5 (92.0) | 15.7 (59.3) | 21.3 | 43.2 | 119.8 | 119.8/√N |
|  | Energetic play | 120 | 58.2 (48.0) | 51.7 (46.7) | 6.5 (37.3) | 33.2 | 26.7 | 74.0 | 74.0/√N |
|  | Total screen time | 120 | 143.8 (85.3) | 139.8 (95.1) | 4.0 (57.5) | 17.3 | 40.6 | 112.6 | 112.6/√N |
|  | Total sleep | 120 | 601.6 (63.2) | 607.0 (59.7) | -5.4 (47.1) | 2.9 | 33.4 | 92.5 | 92.5/√N |
| Open (overall) | Active play | 59 | 179.9 (103.7) | 168.5 (107.4) | 11.4 (68.6) | 18.3 | 48.8 | 135.3 | 135.3/√N |
|  | Energetic play | 59 | 67.7 (56.8) | 65.7 (56.2) | 2.0 (46.9) | 37.6 | 32.9 | 91.3 | 91.3/√N |
|  | Total screen time | 59 | 145.4 (80.7) | 151.7 (105.0) | -6.3 (61.4) | 18.1 | 43.3 | 119.9 | 119.9/√N |
|  | Total sleep | 59 | 596.6 (58.2) | 599.4 (49.3) | -2.8 (43.6) | 2.9 | 30.6 | 84.9 | 84.9/√N |
| Closed (overall) | Active play | 61 | 131.3 (71.6) | 111.4 (63.4) | 19.9 (48.9) | 24.2 | 37.1 | 102.8 | 102.8/√N |
|  | Energetic play | 61 | 49.0 (35.9) | 38.1 (29.9) | 10.9 (24.4) | 29.0 | 18.8 | 52.0 | 52.0/√N |
|  | Total screen time | 61 | 142.3 (90.3) | 128.3 (83.7) | 13.9 (52.1) | 16.6 | 37.9 | 104.9 | 104.9/√N |
|  | Total sleep | 61 | 606.4 (67.8) | 614.3 (67.9) | -7.9 (50.5) | 2.9 | 35.8 | 99.3 | 99.3/√N |

Repeated MBQ-C assessments on Day 7 and Day 10 in the same response format and mode. Overall (all versions) pools all test-retest groups; Open (overall) and Closed (overall) pool paper and electronic modes within each format. Values are min/day. Difference is Day 7 minus Day 10. CV is the mean within-child coefficient of variation; SDCind = 1.96 × √2 × SEM is the individual-level smallest detectable change, and SDCgroup = SDCind/√N is the smallest detectable change in the mean of N participants. CV, coefficient of variation; MBQ-C, Movement Behaviour Questionnaire-Child; SDCgroup, group-level smallest detectable change; SDCind, individual-level smallest detectable change; SD, standard deviation; SEM, standard error of measurement.

#### 3.2.5 Agreement between Formats and Modes

The open- and closed-ended versions showed moderate to good agreement (ICC, 0.57–0.82) with no systematic differences (all Wilcoxon P > 0.4; Table 5). Agreement between the paper and electronic modes was poor for active play (ICC, 0.37) and energetic play (0.38) but good for total screen time (0.86) and total sleep (0.78). In the Bland–Altman analysis, the MBQ-C overestimated total screen time (bias, 19.1 min/day for the open-ended and 20.5 min/day for the closed-ended version) and showed a small bias for total sleep (−5.9 and −2.9 min/day). The 95% limits of agreement were wide (e.g., −103 to 141 min/day for open-ended screen time), and the proportional bias was statistically significant in all four comparisons (all P < 0.05; Supplementary Figure S16).

**Table 5.** Agreement of the Japanese version of the MBQ-C across response formats and modes.

| **Comparison** | **MBQ-C item** | **n** | **ρ [95% CI]** | **ICC [95% CI]** | **Med 1 (Q1–Q3)** | **Med 2 (Q1–Q3)** | **P** |
|---|---|---|---|---|---|---|---|
| Open vs Closed (overall) | Active play | 41 | **0.74 [0.54, 0.87]** | **0.68 [0.47, 0.81]** | 113 (77-180) | 150 (90-197) | 0.494 |
|  | Energetic play | 41 | **0.61 [0.34, 0.78]** | **0.57 [0.31, 0.74]** | 30 (16-60) | 39 (22-75) | 0.560 |
|  | Total screen time | 41 | **0.85 [0.71, 0.92]** | **0.82 [0.68, 0.90]** | 107 (60-150) | 90 (60-144) | 0.884 |
|  | Total sleep | 41 | **0.61 [0.34, 0.78]** | **0.59 [0.34, 0.76]** | 600 (570-630) | 630 (540-660) | 0.870 |
| Paper vs Electronic (open) | Active play | 23 | **0.59 [0.20, 0.82]** | 0.37 [-0.01, 0.67] | 171 (93-201) | 120 (79-167) | 0.061 |
|  | Energetic play | 23 | **0.45 [0.02, 0.73]** | 0.38 [-0.01, 0.67] | 39 (30-63) | 43 (30-58) | 0.244 |
|  | Total screen time | 23 | **0.86 [0.66, 0.95]** | **0.86 [0.70, 0.94]** | 103 (45-147) | 103 (54-160) | 0.320 |
|  | Total sleep | 23 | **0.74 [0.43, 0.90]** | **0.78 [0.55, 0.90]** | 600 (570-600) | 600 (570-605) | 0.832 |

Agreement between response formats (open-ended vs closed-ended) and modes (paper on Day 7 vs electronic on Day 10, open-ended version). Values are weighted weekly averages (min/day). Med 1 and Med 2 are the first and second format or mode named in the Comparison column. ρ with a 95% CI excluding 0, and ICC ≥0.50, are shown in bold. P is the Wilcoxon signed-rank test of the difference between Med 1 and Med 2. CI, confidence interval; ICC, intraclass correlation coefficient; MBQ-C, Movement Behaviour Questionnaire-Child; Q1-Q3, interquartile range; ρ, Spearman rank correlation coefficient.

#### 3.2.6 Practicality

The mean completion time ranged from 5.9 minutes (closed-ended electronic) to 8.9 minutes (open-ended paper) and was shorter in the electronic mode than on paper (Fig. 2). Across the groups, 78% of parents rated the response burden as low (“hardly” or “not very” burdensome), and 60% rated the items as clear (“very” or “somewhat clear”).

**Figure 2.**
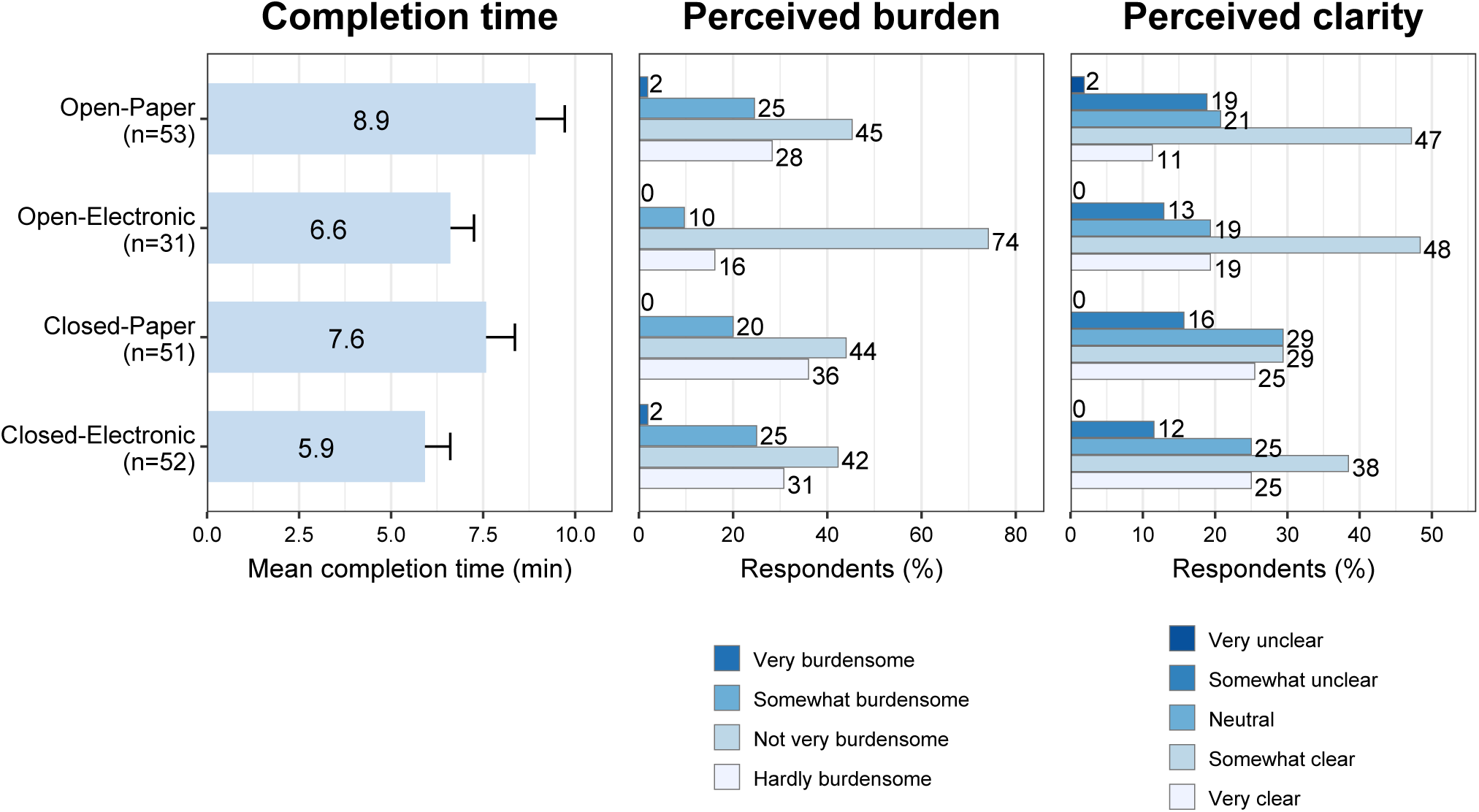
Practicality of the Japanese version of the MBQ-C. The left panel shows the mean time taken to complete the MBQ-C in minutes, with error bars representing the standard error of the mean. The center and right panels show perceived burden and perceived clarity. The number of respondents per group is shown in the left panel. In the Closed-Paper group, completion time and perceived burden were based on 50 respondents, as one respondent did not complete these items (clarity, n = 51). MBQ-C, Movement Behaviour Questionnaire-Child.

#### 3.2.7 Sensitivity Analyses

Restricting the sample to children aged 3–5 years, and varying the valid-day wear-time criterion (≥360 minutes/day and ≥960 minutes/day), did not materially change the validity or reliability estimates (Supplementary Table S17 and S18). When each child’s criterion means were recomputed from days reported as usual, the validity coefficients for the play items tended to be slightly higher (e.g., closed-ended energetic play, ρ = 0.12 to 0.23). Restricting instead to accurately recorded diary days left the coefficients essentially unchanged. Across all three day subsets, the rank order of validity across behaviours was unchanged: weak for active and energetic play, strong for total screen time, and moderate for total sleep (Supplementary Table S19).

## DISCUSSION

We developed and validated a Japanese version of the MBQ-C for assessing preschoolers’ active play, screen time, and sleep, and evaluated its measurement properties and practicality. We extend the evidence for the MBQ-C across cultural settings, following the original [8] and Chinese [9] versions. We additionally examined measurement error, mode agreement, completion time, burden, and clarity, rarely reported for proxy-report tools in this age group [4, 6]. To our knowledge, this is the first study to characterise measurement error in the MBQ-C and to examine the extent of agreement across response modes.

The concurrent validity of active play and energetic play was weak (ρ = 0.12–0.26; Table 2), broadly consistent with the original MBQ-C (ρ = 0.25–0.39 against accelerometer-derived physical activity), the Chinese MBQ-C, and other young-child physical-activity questionnaires [4, 6, 8, 9]. Several factors can attenuate parent proxy-reports of young children’s physical activity: activity at childcare that parents cannot observe, the sporadic nature of the behaviour, recall difficulty, and the mismatch between a typical day and the measurement week. Consistent with this, the cognitive interviews showed that parents were sometimes unsure whether to include childcare time in active play, prompting a revision that added an explanatory note (Supplementary Table S10).

Moreover, active play and accelerometer-derived physical activity are related but not equivalent: the MBQ-C asks about behavioural contexts that parents recognise as active or energetic play, whereas the accelerometer estimates total physical activity or MVPA from movement intensity, regardless of type or context. Nonetheless, the weak but non-negligible associations suggest that parent-reported active play may reflect, to some extent, children’s relative levels of accelerometer-derived physical activity. The weak validity of the active-play items should thus be interpreted not as a simple failure to measure physical activity, but in light of the limits of proxy reporting and the conceptual difference between what the questionnaire and the accelerometer capture. Mean acceleration (ENMO), one of the accelerometer-derived comparison metrics used in the original validation [8], showed higher coefficients (overall ρ = 0.32 for active play, and 0.34 and 0.33 for energetic play in the open- and closed-ended versions; Table 2). These values were comparable to those reported for the original MBQ-C. Because ENMO reflects overall movement volume, whereas classifier-derived Total PA and MVPA estimate time spent in activity categories, parent-reported active play may capture children’s general activeness more than device-classified activity time.

This interpretation is supported by the reliability and measurement-error results. Test–retest reliability was moderate to good (ICC = 0.71–0.79 for active play, 0.66–0.69 for energetic play; Table 3), so parents’ responses were not entirely unstable. However, measurement error was larger than for the other behaviours (within-child CV = 21.3% for active play and 33.2% for energetic play; Table 4).

Energetic play showed greater re-administration variability than screen time (17.3%) or sleep (2.9%), reflecting the difficulty of stably estimating habitual higher-intensity play. The individual-level SDC (Table 4) provides the basis for estimating group-level precision. Because random error partly averages out across participants, detectable differences in group means are much smaller than individual-level SDCs [36]. For example, for active play, the detectable difference is about 12 min/day with 100 children and about 4 min/day with 1000 children (Table 4). However, when interpreting longitudinal change, systematic between-occasion differences should also be considered because they do not decrease with larger sample size. In our data, active play showed a systematic difference of about +16 min/day between Day 7 and Day 10 (Table 4), so longitudinal changes should be interpreted in light of both this bias and the group-level random measurement error. Future work should refine item wording and instructions, including how to handle childcare and unobserved periods.

Screen time showed strong validity against the diary-based criterion (ρ = 0.78–0.85; Table 2), comparable to or higher than the original MBQ-C’s correlation with the 24-hour time-use diary (ρ = 0.71–0.79) [8]. The original used a 2-day diary (Days 2 and 5), whereas the present study recorded screen time daily over seven days with morning and evening reminders, so the criterion may have been more stable. Test–retest reliability was also good (ICC = 0.79–0.81; Table 3), and the CV (17.3%) was smaller than for the play items (Table 4), indicating that screen time is relatively easy for parents to recall and corresponds well to the diary. However, the individual-level SDC was large (SDCind = 112.6 min/day), and the Bland–Altman analysis showed wide limits of agreement, overestimation, and proportional bias (Table 4, Supplementary Figure S16). At the group level, screen-time means can be compared to within about 4 min/day in a survey of 1000 children (about 11 min/day for 100; Table 4), so the screen-time items are well suited to surveillance and group comparison.

Sleep showed moderate validity (ρ = 0.48–0.58; Table 2), slightly lower than the original MBQ-C’s total-sleep correlation with the 24-hour time-use diary (ρ = 0.58–0.70) [8]. This likely reflects a more stringent criterion: the original used a 2-day parent-reported diary, so questionnaire and criterion shared the same method (parent report), which tends to inflate validity, whereas the present criterion combined a 7-day diary with accelerometer-based bed-rest determination, reducing this shared-method dependence. The lower coefficient may therefore be a more conservative estimate rather than poorer questionnaire performance.

In terms of measurement error, however, sleep was the most stable. Test–retest reliability for total sleep was moderate (ICC = 0.68–0.72; Table 3), and the CV (2.9%) was clearly smaller than for active play, energetic play, and screen time (Table 4), indicating that sleep is a relatively stable daily routine and easy for parents to report. Nonetheless, the SEM was 33.4 min/day and the individual-level SDC was 92.5 min/day (Table 4). Because total sleep had the smallest within-child variability (CV = 2.9%), its group-level detectable difference was also small (about 3 min/day for a survey of 1000 children and 9 min/day for 100), supporting its use to monitor and rank sleep duration across groups. Asking about weekdays and weekends separately, or including bedtime and wake time, could also capture social jetlag, the weekday–weekend discrepancy in sleep timing [39], which has been associated with greater adiposity in school-aged children [40]. This warrants further development.

Agreement between response formats and modes has implications for large-scale surveys. The open- and closed-ended versions showed moderate-to-good agreement with no systematic differences (Table 5), supporting the original finding that both formats can be used [8] and indicating that the format can be chosen according to the research aim and setting. Completion time tended to be shorter for the closed-ended version, and most parents rated the burden as low and the items as clear (Fig. 2), suggesting that the Japanese MBQ-C can generally be administered with low burden regardless of format.

The response mode requires cautious interpretation. Paper–electronic agreement was a secondary analysis using the open-ended version with a small sample (n = 23), so strong conclusions should be avoided; the ICCs were poor for active play (0.37) and energetic play (0.38) but good for total screen time (0.86) and total sleep (0.78) (Table 5). Because completion time was shorter electronically, parents may have responded more quickly than on paper and entered answers before recalling fully, particularly for the play items. This cannot be confirmed here, so the influence of mode on the response process and the active-play values should be examined in a larger sample. For now, studies in which the play items are a primary outcome should standardise the response mode.

This study has several strengths. First, it followed a rigorous COSMIN-based translation and cross-cultural adaptation process and confirmed comprehensibility through cognitive interviews. Second, the criterion measurement was robust: Total PA and MVPA were estimated with a machine-learning method [17], overcoming a key limitation of cut-points in young children, and sleep was assessed by combining a daily diary with hip-accelerometer bed-rest determination developed for young children [23]. Third, it examined properties not reported for the original MBQ-C—measurement error, paper– electronic agreement, and practicality—more fully characterising its measurement properties.

This study has several limitations. First, the target age and developmental stage do not fully match those of the original: the original MBQ-C targets children from the onset of independent walking to 5 years, whereas this study targeted children aged 3–6 years. In Japan, children may attend kindergarten from 3 years until elementary school entry, so 6-year-olds may still be in preschool settings [41]. The sensitivity analysis restricted to ages 3–5 years gave broadly similar results, supporting robustness (Supplementary Table S17), but the absence of younger, newly walking children leaves it unclear whether the Japanese MBQ-C applies to that group. Second, the paper–electronic comparison was a secondary analysis with a small sample, so between-mode agreement, particularly for the play items, should be regarded as exploratory. Third, although based on the 24-hour movement behaviour framework, the study measured screen time and active play rather than the full 24-hour time-use composition, requiring caution when interpreting results as whole-day behaviour. Fourth, the criteria for total screen time and total sleep relied substantially on the same parent-completed diary, and the MBQ-C is itself parent-reported, so common-method variance may have inflated their apparent validity. Finally, responsiveness to change, which the original developers highlighted as a priority for future MBQ research, could not be evaluated in this cross-sectional study and should be examined in future longitudinal work.

## CONCLUSIONS

The Japanese MBQ-C provides a practical option for public health research and practice to assess preschoolers’ active play, screen time, and sleep at the group level. The findings support its use for surveillance, group-level comparisons, and the evaluation of interventions and programmes delivered at scale. They also provide practical guidance for interpreting MBQ-C measures according to the research aim and survey setting.

## Supporting information

medRxiv_v2_MBQ-C_Japanese_Supplementary_20261120

## LIST OF ABBREVIATIONS

CI: confidence interval
COSMIN: COnsensus-based Standards for the selection of health Measurement INstruments
CV: coefficient of variation
ECEC: early childhood education and care
ENMO: Euclidean Norm Minus One
ICC: intraclass correlation coefficient
JPY: Japanese yen
MBQ-C: Movement Behaviour Questionnaire–Child
MVPA: moderate-to-vigorous physical activity
PA: physical activity
SD: standard deviation
SDC: smallest detectable change
SDCind: individual-level smallest detectable change
SDCgroup: group-level smallest detectable change
SEM: standard error of measurement
SSF-PAQ: Sasakawa Sports Foundation Preschoolers’ Activity Questionnaire
STROBE: Strengthening the Reporting of Observational Studies in Epidemiology.

## DECLARATIONS

### Ethics approval and consent to participate

Both phases of the study were approved by the Meijo University Human Research Ethics Committee (approval nos. 2024-17-2 and 2024-43). Informed consent was obtained from a parent or guardian of each participating child (online for the translation phase and in writing for the validation study). The study was conducted in accordance with the Declaration of Helsinki.

### Consent for publication

Not applicable.

### Availability of data and materials

The datasets used and/or analysed during the current study are available from the corresponding author (K.K.) on reasonable request.

### Authors’ contributions

K.K.: Conceptualization, Funding acquisition, measurement and data analysis, and drafting of the manuscript. M.Y., A.K., S.I., M.N., and R.T.: Methodology as members of the questionnaire review team. M.Y.: Coding of the interview data. A.K.: Support for accelerometer-based data collection. R.B. and E.H.J.: Methodology through back-translation. S.G.T.: Methodological advice on accelerometer-based measurement and analysis. All authors reviewed and edited the manuscript and approved the final version.

## Acknowledgements

We thank Kosuke Mase (Graduate School of Bioagricultural Sciences, Nagoya University) for his assistance with the forward translation of the Japanese MBQ-C. During the preparation of this manuscript, the authors used generative artificial intelligence tools for language editing, proofreading, and assistance in reviewing analysis code. The authors reviewed and edited the content and take full responsibility for the final manuscript.

## Additional file 1: supplementary materials (.pdf)

**Supplementary Table S1.** STROBE checklist for cross-sectional studies.

**Supplementary Table S2.** Interview guide for the cognitive interviews.

**Supplementary Table S3.** Codebook for coding the cognitive interviews.

**Supplementary Table S4.** Allocation of participants to the seven groups.

**Supplementary Material S5.** The 7-day 24-hour time-use diary.

**Supplementary Figure S6.** Determination and correction of sleep periods.

**Supplementary Figure S7.** Mahalanobis distance used to identify MVPA outliers.

**Supplementary Figure S8.** Participant flow for the cognitive interviews.

**Supplementary Table S9.** Participant characteristics for cognitive interview rounds 1 and 2.

**Supplementary Table S10.** Key revisions made through the cognitive interviews.

**Supplementary Material S11.** Japanese version of the MBQ-C, Open Version.

**Supplementary Material S12.** Japanese version of the MBQ-C, Closed-ended Version.

**Supplementary Figure S13.** Participant flow for the validation study.

**Supplementary Table S14.** Characteristics of the participants included in each analysis.

**Supplementary Table S15.** Detailed validity and reliability by indicator and day type.

**Supplementary Figure S16.** Bland–Altman analysis for total screen time and total sleep.

**Supplementary Table S17.** Sensitivity analysis restricted to children aged 3–5 years.

**Supplementary Table S18.** Sensitivity analysis varying the wear-time criterion.

**Supplementary Table S19.** Sensitivity analysis by diary-recording quality.

## References

1. Rollo S, Antsygina O, Tremblay MS. The whole day matters: understanding 24-hour movement guideline adherence and relationships with health indicators across the lifespan. J Sport Health Sci. 2020;9(6):493–510. 10.1016/j.jshs.2020.07.004

2. World Health Organization. Guidelines on physical activity, sedentary behaviour and sleep for children under 5 years of age. Geneva: World Health Organization; 2019.

3. Hidding LM, Chinapaw MJM, van Poppel MNM, Mokkink LB, Altenburg TM. An updated systematic review of childhood physical activity questionnaires. Sports Med. 2018;48(12):2797–2842. 10.1007/s40279-018-0987-0

4. Phillips SM, Summerbell C, Hobbs M, Hesketh KR, Saxena S, Muir C, et al. A systematic review of the validity, reliability, and feasibility of measurement tools used to assess the physical activity and sedentary behaviour of pre-school aged children. Int J Behav Nutr Phys Act. 2021;18(1):141. 10.1186/s12966-021-01132-9

5. Kohl HW 3rd, Fulton JE, Caspersen CJ. Assessment of physical activity among children and adolescents: a review and synthesis. Prev Med. 2000;31(2 Pt 2):S54–S76. 10.1006/pmed.1999.0542

6. Arts J, Gubbels JS, Verhoeff AP, Chinapaw MJM, Lettink A, Altenburg TM. A systematic review of proxy-report questionnaires assessing physical activity, sedentary behavior and/or sleep in young children (aged 0-5 years). Int J Behav Nutr Phys Act. 2022;19(1):18. 10.1186/s12966-022-01251-x

7. Byrne R, Terranova CO, Chai LK, Brookes DSK, Trost SG. Cognitive testing of items measuring movement behaviours in young children aged zero to five years: development of the Movement Behaviour Questionnaires for -Baby (MBQ-B) and -Child (MBQ-C). Children (Basel). 2023;10(9):1554. 10.3390/children10091554

8. Trost SG, Terranova CO, Brookes DSK, Chai LK, Byrne RA. Reliability and validity of rapid assessment tools for measuring 24-hour movement behaviours in children aged 0-5 years: the Movement Behaviour Questionnaire Baby (MBQ-B) and child (MBQ-C). Int J Behav Nutr Phys Act. 2024;21(1):43. 10.1186/s12966-024-01596-5

9. Song H, Lu N, Wang J, Lau PWC, Zhou P. Validity and reliability of the movement behaviour questionnaire child in Chinese preschoolers. Front Pediatr. 2025;13:1544738. 10.3389/fped.2025.1544738

10. Komura K, Nagano M, Takenaga R, Kyan A, Inoue S, Yamakita M. >Validity and reliability of the SSF Preschoolers’ Activity Questionnaire (SSF-PAQ) for assessing active play, screen time, and sleep in Japanese preschool children. medRxiv. 2026. Manuscript in preparation. 10.64898/2026.11.17.26368286

11. von Elm E, Altman DG, Egger M, Pocock SJ, Gøtzsche PC, Vandenbroucke JP; STROBE Initiative. The Strengthening the Reporting of Observational Studies in Epidemiology (STROBE) statement: guidelines for reporting observational studies. Lancet. 2007;370(9596):1453–1457. 10.1016/S0140-6736(07)61602-X

12. Mokkink LB, Prinsen CAC, Patrick DL, Alonso J, Bouter LM, de Vet HCW, et al. COSMIN Study Design checklist for patient-reported outcome measurement instruments. Amsterdam: COSMIN; 2019.

13. Willis GB. Cognitive interviewing: a tool for improving questionnaire design. Thousand Oaks, CA: SAGE Publications; 2004.

14. Tourangeau R. Cognitive sciences and survey methods. In: Jabine TB, Straf ML, Tanur JM, Tourangeau R, editors. Cognitive aspects of survey methodology: building a bridge between disciplines. Washington, DC: National Academy Press; 1984. p. 73–100.

15. Gwet KL. Computing inter-rater reliability and its variance in the presence of high agreement. Br J Math Stat Psychol. 2008;61(Pt 1):29–48. 10.1348/000711006X126600

16. Määttä S, Ray C, Vepsäläinen H, Lehto E, Kaukonen R, Ylönen A, et al. Parental education and pre-school children’s objectively measured sedentary time: the role of co-participation in physical activity. Int J Environ Res Public Health. 2018;15(2):366. 10.3390/ijerph15020366

17. Ahmadi MN, Pavey TG, Trost SG. Machine learning models for classifying physical activity in free-living preschool children. Sensors (Basel). 2020;20(16):4364. 10.3390/s20164364

18. Ahmadi MN, Trost SG. Device-based measurement of physical activity in pre-schoolers: comparison of machine learning and cut point methods. PLoS One. 2022;17(4):e0266970. 10.1371/journal.pone.0266970

19. Migueles JH, Ahmadi M, Trost SG. actimetric: Classifies Accelerometer Data Into Physical Activity Types. R package version 0.1.5. 2025. https://github.com/PhysicalActivityOpenTools/actimetric. Accessed 4 July 2026.

20. Ahmadi MN, Nathan N, Sutherland R, Wolfenden L, Trost SG. Non-wear or sleep? Evaluation of five non-wear detection algorithms for raw accelerometer data. J Sports Sci. 2020;38(4):399–404. 10.1080/02640414.2019.1703301

21. van Hees VT, Sabia S, Anderson KN, Denton SJ, Oliver J, Catt M, et al. A novel, open access method to assess sleep duration using a wrist-worn accelerometer. PLoS One. 2015;10(11):e0142533. 10.1371/journal.pone.0142533

22. van Hees VT, Sabia S, Jones SE, Wood AR, Anderson KN, Kivimäki M, et al. Estimating sleep parameters using an accelerometer without sleep diary. Sci Rep. 2018;8(1):12975. 10.1038/s41598-018-31266-z

23. Tracy JD, Donnelly T, Sommer EC, Heerman WJ, Barkin SL, Buchowski MS. Identifying bedrest using waist-worn triaxial accelerometers in preschool children. PLoS One. 2021;16(1):e0246055. 10.1371/journal.pone.0246055

24. van Hees VT, Gorzelniak L, Dean León EC, Eder M, Pias M, Taherian S, et al. Separating movement and gravity components in an acceleration signal and implications for the assessment of human daily physical activity. PLoS One. 2013;8(4):e61691. 10.1371/journal.pone.0061691

25. Mendoza JA, Baranowski T, Jaramillo S, Fesinmeyer MD, Haaland W, Thompson D, et al. Fit 5 Kids TV reduction program for Latino preschoolers: a cluster randomized controlled trial. Am J Prev Med. 2016;50(5):584–592. 10.1016/j.amepre.2015.09.017

26. Werner H, Molinari L, Guyer C, Jenni OG. Agreement rates between actigraphy, diary, and questionnaire for children’s sleep patterns. Arch Pediatr Adolesc Med. 2008;162(4):350–358. 10.1001/archpedi.162.4.350

27. Sasakawa Sports Foundation. Online survey on out-of-school participation in physical activity among preschool children in Japan. Tokyo: Sasakawa Sports Foundation; 2024. https://www.ssf.or.jp/en/features/japans_data_plus_sports/e0022.html. Accessed 27 June 2026.

28. Addy CL, Trilk JL, Dowda M, Byun W, Pate RR. Assessing preschool children’s physical activity: how many days of accelerometry measurement. Pediatr Exerc Sci. 2014;26(1):103–109. 10.1123/pes.2013-0021

29. Hinkley T, O’Connell E, Okely AD, Crawford D, Hesketh K, Salmon J. Assessing volume of accelerometry data for reliability in preschool children. Med Sci Sports Exerc. 2012;44(12):2436–2441. 10.1249/MSS.0b013e3182661478

30. Breau B, Coyle-Asbil HJ, Vallis LA. The use of accelerometers in young children: a methodological scoping review. J Meas Phys Behav. 2022;5(3):185–201. 10.1123/jmpb.2021-0049

31. Signorell A. DescTools: Tools for Descriptive Statistics. R package version 0.99.60. 2025. https://CRAN.R-project.org/package=DescTools. Accessed 22 Sept 2026.

32. Schober P, Boer C, Schwarte LA. Correlation coefficients: appropriate use and interpretation. Anesth Analg. 2018;126(5):1763–1768. 10.1213/ANE.0000000000002864

33. Gamer M, Lemon J, Fellows I, Singh P. irr: Various Coefficients of Interrater Reliability and Agreement. R package version 0.84.1. 2019. https://CRAN.R-project.org/package=irr. Accessed 22 Sept 2026.

34. Koo TK, Li MY. A guideline of selecting and reporting intraclass correlation coefficients for reliability research. J Chiropr Med. 2016;15(2):155–163. 10.1016/j.jcm.2016.02.012

35. Beaton DE. Understanding the relevance of measured change through studies of responsiveness. Spine (Phila Pa 1976). 2000;25(24):3192–3199. 10.1097/00007632-200012150-00015

36. de Vet HCW, Terwee CB, Mokkink LB, Knol DL. Measurement in Medicine: A Practical Guide. Cambridge: Cambridge University Press; 2011.

37. Bingham DD, Costa S, Clemes SA, Routen AC, Moore HJ, Barber SE. Accelerometer data requirements for reliable estimation of habitual physical activity and sedentary time of children during the early years - a worked example following a stepped approach. J Sports Sci. 2016;34(20):2005–2010. 10.1080/02640414.2016.1149605

38. Ministry of Health, Labour and Welfare. 2024 (Reiwa 6) comprehensive survey of living conditions. Tokyo: Ministry of Health, Labour and Welfare; 2025.

39. Wittmann M, Dinich J, Merrow M, Roenneberg T. Social jetlag: misalignment of biological and social time. Chronobiol Int. 2006;23(1-2):497–509. 10.1080/07420520500545979

40. Stoner L, Beets MW, Brazendale K, Moore JB, Weaver RG. Social jetlag is associated with adiposity in children. Glob Pediatr Health. 2018;5:2333794X18816921. 10.1177/2333794X18816921

41. Ministry of Justice, Japan. School Education Act (Act No. 26 of 1947). Japanese Law Translation Database System. https://www.japaneselawtranslation.go.jp/en/laws/view/4573/en. Accessed 16 June 2026.

