## Supplementary material for "Validity and reliability of the Japanese version of the Movement Behaviour Questionnaire–Child (MBQ-C) for assessing active play, screen time, and sleep in preschool children": medRxiv_v2_MBQ-C_Japanese_Supplementary_20261120

### Additional file 1: supplementary materials

**Affiliations**
1. Department of Liberal Arts, Faculty of Agriculture, Meijo University, Aichi, Japan
2. Graduate School of Environmental and Human Sciences, Meijo University, Aichi, Japan
3. Faculty of Nursing, Yamanashi Prefectural University, Yamanashi, Japan
4. Faculty of Medicine, University of the Ryukyus, Okinawa, Japan
5. Center for Social Common Capital Beyond 2050, Office of Institutional Advancement and Communications, Kyoto University, Kyoto, Japan
6. Department of Preventive Medicine and Public Health, Tokyo Medical University, Tokyo, Japan
7. International College of Arts and Sciences, Fukuoka Women's University, Fukuoka, Japan
8. Sasakawa Sports Foundation, Tokyo, Japan
9. Department of English, Kansai Medical University, Osaka, Japan
10. Medical Education Center, Nihon University School of Medicine, Tokyo, Japan
11. School of Human Movement and Nutrition Sciences, The University of Queensland, Brisbane, Australia

**Corresponding author**
Keisuke Komura, Department of Liberal Arts, Faculty of Agriculture, Meijo University.


#### Index of Additional file 1

#### Supplementary Table S1. STROBE checklist for cross-sectional studies

| No. | STROBE recommendation (abridged) | Status | Where reported / reason |
|---|---|---|---|
| **Title and abstract** | | | |
| 1a | Study design indicated with a common term in title or abstract | Reported | Abstract (Methods: "cross-sectional validation study"; concurrent validity against accelerometry and diary). Title states "validity and reliability". |
| 1b | Balanced, informative summary in the abstract | Reported | Structured Abstract (Background/Methods/Results/Conclusions). |
| **Introduction** | | | |
| 2 | Scientific background and rationale | Reported | Background (24-h movement guidelines; need for feasible valid proxy measures; gap; MBQ-C). |
| 3 | Specific objectives / prespecified aims | Reported | Background, final paragraph (primary aim: concurrent validity and test–retest reliability; secondary aims: measurement error, format/mode agreement, practicality). |
| **Methods** | | | |
| 4 | Key elements of study design, presented early | Reported | Methods 2.1 Study Design (two phases; cross-sectional validation). |
| 5 | Setting, locations, relevant dates incl. recruitment and data collection | Reported | 2.1 (July 2024–Apr 2025; May–June and Oct–Dec 2025); 2.3.1 (centers); 2.3.2 (10-day protocol). |
| 6 | Eligibility criteria; sources and methods of participant selection | Reported | 2.3.1 (convenience-sampled centers, children aged 3–6 years, consent, group allocation); 2.2.2 (cognitive-interview participants). |
| 7 | Clearly define outcomes, exposures, predictors, modifiers | Reported | MBQ-C indices (active/energetic play, screen time, sleep) and criteria defined in 2.3.3–2.3.6; "index vs criterion" framing rather than exposure/outcome (measurement-property study). |
| 8 | Sources of data and details of measurement; comparability of methods | Reported | 2.3.3 (accelerometry, machine-learning classifier), 2.3.4 (diary), 2.3.5 (sleep, accelerometer+diary), 2.3.6 (practicality). Format (open/closed) and mode (paper/electronic) comparability addressed in 2.3.8 and Table 5. |
| 9 | Efforts to address potential sources of bias | Reported | 2.2.2 (socioeconomic diversity of interviewees to counter the original study's education bias); 2.3.2 (twice-daily reminders, refrigerator clips to reduce missing diary entries); 2.3.7 (exclusion of MVPA outliers via Mahalanobis distance); diary-quality day subsets in 2.3.8; proxy-report and mode bias discussed in Discussion (Limitations). |
| 10 | How the study size was determined | Reported | 2.3.1 (a priori sample-size calculation: $\rho$=0.30, $\alpha$=0.05, power=0.80 → 85/format, 170 valid datasets; recruitment yield assumptions). |
| 11 | Handling of quantitative variables; groupings and rationale | Reported | 2.3.7 (data reduction and truncation per the original MBQ-C; weighted weekly averages; open-ended hours/minutes and closed-ended category midpoints); 2.3.8 ($\rho$ and ICC interpretation thresholds). |
| 12a | Statistical methods, including control for confounding | Reported | 2.3.8 (Spearman $\rho$ with 95% CI using Fisher's z transformation (DescTools); ICC two-way absolute-agreement single-measure; Wilcoxon signed-rank; measurement error via linear mixed-effects model; Bland–Altman). |
| 12b | Methods to examine subgroups and interactions | Reported | 2.3.8 (weekday/weekend; open vs closed format; paper vs electronic mode); detailed indicators in Supplementary Table S15. |
| 12c | How missing data were addressed | Reported | 2.3.8 (complete-case analysis per outcome, with n reported for each analysis); 2.3.5 (imputation of missing sleep onset/offset endpoints with within-child circular means). |

Supplementary Table S1. Continued.

| No. | STROBE recommendation (abridged) | Status | Where reported / reason |
|---|---|---|---|
| 12d | Methods accounting for sampling strategy | Reported | 2.3.1 (convenience sampling at center level; allocation to seven groups balancing centers and grades). No probability-sampling weights were applied. |
| 12e | Sensitivity analyses | Reported | 2.3.8 and 3.2.7 (age restriction to 3–5 years; wear-time criteria 360/600/960 min; usual-day and accurate-day diary subsets); Supplementary Table S17–19. |
| **Results** | | | |
| 13a | Numbers of individuals at each stage | Reported | 3.2.1 (208 consented → 170 valid); participant flow in Supplementary Figure S13; cognitive-interview flow in Supplementary Figure S8. |
| 13b | Reasons for non-participation at each stage | Reported | 3.2.1 (non-measurement, device failure, insufficient valid days, MVPA outliers); detailed in Supplementary Figure S13. |
| 13c | Flow diagram | Reported | Supplementary Figure S13 (validation study); Supplementary Figure S8 (cognitive interviews). |
| 14a | Characteristics of participants | Reported | Table 1; 3.2.1 and Supplementary Table S14 (age, sex, center type, respondent); cognitive-interview participants in Supplementary Table S9. |
| 14b | Number with missing data for each variable | Reported | 2.3.8 (complete cases; per-analysis n); analysis-specific n shown in Tables 2–5 and Supplementary Table S14. |
| 15 | Report outcome data / summary measures | Reported | Tables 2–5; 3.2.2–3.2.6 (validity, reliability, measurement error, agreement, practicality). |
| 16a | Estimates and their precision (e.g., 95% CI) | Reported | ρ and ICC with 95% CIs in Tables 2, 3, 5; SEM/SDC in Table 4; Bland–Altman bias and limits of agreement in Supplementary Figure S16. |
| 16b | Category boundaries when continuous variables were categorized | Reported | 2.3.8 (ρ: weak/moderate/strong per Schober 2018; ICC: poor/moderate/good/excellent per Koo & Li 2016); 2.3.6 (practicality response categories). |
| 16c | Translate relative risk into absolute risk if relevant | Not applicable | No risk or effect estimates are reported; this is a correlation/agreement (measurement-property) study without risk measures. |
| 17 | Other analyses (subgroups, interactions, sensitivity) | Reported | 3.2.7 (sensitivity analyses); Supplementary Table S17–19; weekday/weekend and component detail in Supplementary Table S15. |
| **Discussion** | | | |
| 18 | Summarise key results with reference to objectives | Reported | Discussion, paragraph 1. |
| 19 | Limitations, including sources of bias/imprecision and their direction/magnitude | Reported | Discussion, Limitations paragraph (age/developmental mismatch with original; small paper-vs-electronic subsample; partial coverage of the 24-h composition); measurement-error magnitudes (SDC) discussed by behaviour. |
| 20 | Cautious overall interpretation | Reported | Discussion (behaviour-specific interpretation; group-level vs individual-level use; comparison with original and Chinese versions and prior reviews). |
| 21 | Generalisability (external validity) | Reported | Discussion and Conclusions (applicability to large-scale Japanese surveys; caution for children below the studied age range and for individual-level use). |
| **Other information** | | | |
| 22 | Source of funding and role of funders (present study; and original study if applicable) | Reported | Declarations — Funding (DAIKO FOUNDATION FY2024 grant; funder had no role). |

Note. This completed checklist was based on the STROBE Checklist for cross-sectional studies. STROBE, Strengthening the Reporting of Observational Studies in Epidemiology. Recommendations were abridged for presentation.

#### Supplementary Table S2. Interview guide for the cognitive interviews

| Major item | Sub-item | Interviewer instructions / questions |
|---|---|---|
| **Introduction** | **Self-introduction** | Hi [participant name], welcome to the interview today. My name is [name], thank you for your time to participate today. |
| | **Purpose explanation** | The purpose of this interview is to understand how parents interpret and respond to questions about their child's usual behaviour. First, I will display the questionnaire items and response options on the screen for you to read aloud. Then, I will ask you to verbalize your thought process as you decide on an answer. We are not looking for "correct" answers but are more interested in how you understand the questions and decide on responses. Even if you are unsure about an answer, please verbalize your thoughts, including your uncertainty. This may feel unfamiliar initially, but we will practice, and it will become easier as we proceed. Please don't worry. |
| | **Recording confirmation** | This interview will be recorded, but the recordings will only be used for research purposes. No personally identifiable information will be shared. Do I have your permission to record? |
| | **Start recording** | *[Start Recording]* |
| | **Identifying the child for the parent's responses** | Today, please focus on your child aged 3 to 6. If you have more than one, refer to the child from the previous survey. Could you tell me the child's name and age? |
| **Warm-up** | **Practice think-aloud process** | Before starting the main interview, let's practice verbalizing your thoughts while deciding on an answer. Please read the following question aloud, think about your response, and explain your thought process out loud before giving your final answer.<br>*[Displayed on screen in Japanese]* How many rooms are in your house?<br>(If the participant pauses) Please tell me what you're thinking. |
| **Introductory explanation** | **Instructions 1** | I will now display the introductory text for the questionnaire on the screen. Please read it aloud. This is not a question, so after reading, you do not need to verbalize your thought process. Once you've finished reading, I will ask you some questions. Please read the following text aloud.<br>*[Displayed on screen in Japanese]* This survey will ask you questions about your child's movement behaviours (activity, screen time, and sleep) on a typical day. A typical day is a day when your child does things they normally do. |
| | **Questions 1** | Was there anything you found difficult to understand?<br>(If yes) What was difficult? |
| | **Instructions 2** | Please continue reading the following explanation aloud.<br>*[Displayed on screen in Japanese]* For questions about how much time your child spends in these behaviours, please provide an answer to both hours and minutes, e.g., 2 hours 0 minutes, 0 hours 30 minutes. Please respond to all the questions as best as you can. |
| | **Questions 2** | Was there anything you found difficult to understand?<br>(If yes) What was difficult? |
| **Physical activity section Q1A** | **Purpose confirmation** | We will now review the questionnaire items. There are no right or wrong answers. What's most important is understanding how you arrive at your answers. |
| | **Instructions 1** | Here is the first question. Please read it aloud and talk through your thoughts while deciding on your response.<br>*[Displayed on screen in Japanese]* Q1A. Thinking about the past week, on |

Supplementary Table S2. Continued.

| Major item | Sub-item | Interviewer instructions / questions |
|---|---|---|
| | | a TYPICAL WEEKDAY, how much time did your child spend in active play? Active play includes activities such as walking, running, dancing, climbing, playing with balls, riding bikes or scooters, or swimming.<br>hours minutes<br>(If the participant pauses) Please tell me what you're thinking. |
| | **Question 1** | Was there anything you found difficult to understand?<br>(If yes) What was difficult? |
| | **Question 2** | Is it easy to write your response with this answer format? |
| | **Question 3** | Next, please answer the same question using different response options.<br>*[Displayed on screen in Japanese]* [0] 0 min per day, [1] Between 1 and 30 min per day, [2] Between 30 and 60 min per day, [3] Between 1 and 2 hrs per day, [4] Between 2 and 3 hrs per day, [5] Between 3 and 4 hrs per day, [6] More than 4 hrs per day.<br>Did you find an option that fits your answer perfectly, or did it feel slightly different?<br>(If slightly different) What response options would be better? |
| **Physical activity section Q1B-Q2B** | For Q1B–Q2B, repeat the Q1A procedure using each item's wording. | |
| **Screen time section Q3A** | **Instructions 1** | Here's the next question. Read it aloud and tell me what you're thinking.<br>*[Displayed on screen in Japanese]* This section is about your child's screen time. Q3A. Thinking about the past week, on a TYPICAL WEEKDAY, how much time did your child spend watching television programs, videos/internet clips or movies on a television, computer, or portable/mobile device such as iPad, tablet or smartphone?<br>hours minutes<br>(If the participant pauses) Please tell me what you're thinking. |
| | **Question 1** | Was there anything you found difficult to understand?<br>(If yes) What was difficult? |
| | **Question 2** | Now, the same question with different response options.<br>*[Displayed on screen in Japanese]* [0] 0 min per day, [1] Between 1 and 15 min per day, [2] Between 15 and 30 min per day, [3] Between 30 and 60 min per day, [4] Between 1 and 1½ hrs per day, [5] Between 1½ and 2 hrs per day, [6] Between 2 and 3 hrs per day, [7] More than 3 hrs per day.<br>Did you find an option that fits your answer perfectly, or did it feel slightly different?<br>(If slightly different) What response options would be better? |
| **Screen time section Q3B-Q6B** | For Q3B–Q6B, repeat the Q3A procedure using each item's wording. For Q5A, additionally ask: "If screen time outside the home, such as in the car, while shopping, or in a restaurant, is included, would your answer change?" | |
| **Sleep section Q7** | **Instructions 1** | Here's the next question. Read it aloud and tell me what you're thinking.<br>*[Displayed on screen in Japanese]* This section is about your child's sleep. Q7. Thinking about the past week, on a TYPICAL NIGHT, how much time did your child sleep in total during the night?<br>hours minutes<br>(If the participant pauses) Please tell me what you're thinking. |
| | **Question 1** | Was there anything you found difficult to understand?<br>(If yes) What was difficult? |
| | **Question 2** | Now, the same question with different response options.<br>*[Displayed on screen in Japanese]* [1] Less than 6 hrs per night, [2] Between 6 and 8 hrs per night, [3] Between 8 and 10 hrs per night, [4] |

Supplementary Table S2. Continued.

| **Major item** | **Sub-item** | **Interviewer instructions / questions** |
|---|---|---|
| | | Between 10 and 12 hrs per night , [5] Between 12 and 14 hrs per night, [6] More than 14 hrs per night.<br>Did you find an option that fits your answer perfectly, or did it feel slightly different?<br>(If slightly different) What response options would be better? |
| **Sleep section Q8-Q9** | Same instructions and questions as Q7. | |
| **About the response Format** | **Comparison of response Formats** | Which do you prefer: writing the time yourself or choosing from the response options? Why do you prefer that? |
| **Interview close** | **Thank-you gift card** | The interview is now complete. Thank you for your time. A 2,000-yen gift card will be mailed to your provided address. Please return the receipt using the included envelope once you receive it.<br>Thank you so much for participating in the interview today. |

The interview content was designed using the think-aloud method and verbal probing (Willis, 2004). Questionnaire items and response options are shown in English based on the original MBQ-C Open and Closed versions; during the interviews, the Japanese version developed in this study was presented to participants on slides. Bracketed placeholders were replaced during the interview. Q, questionnaire item.

#### Supplementary Table S3. Codebook for coding the cognitive interviews

| Main category | Sub-category | Definition | Example |
|---|---|---|---|
| **1. Comprehension of the question** | **1a. Unclear question intent** | The question's intent is unclear, causing confusion for the respondent. | *"I wasn't sure if they were asking about how often my child exercises or how long they exercise each time."* |
| | **1b. Unclear meaning of terms** | Specific words or phrases are misunderstood or confusing to the respondent. | *"Does 'screen time' mean just phones, or does it also include TV and tablets? I wasn't sure."* |
| **2. Retrieval from memory** | **2a. Difficulty in accurate recall** | Unable to accurately recall the necessary information. | *"I don't really remember how much time my child spent being active last week."* |
| | **2b. Confusion in recall strategies** | Unsure how to recall or respond. | *"Our weekday and weekend schedules are so different, so I didn't know how to answer."* |
| **3. Decision processes** | **3a. Lack of motivation** | Responds without devoting sufficient mental effort to the question. | *"I just gave an answer without thinking deeply about it."* |
| | **3b. Social desirability bias** | Prioritizes socially desirable responses. | *"I said more time to make my child seem more active."* |
| **4. Response processes** | **4. Mapping issues** | Unable to map the answer to the provided options. | *"None of the options fit our situation exactly."* |
| **5. Other issues** | **5. Other issues** | Issues related to the question or response environment that don't fit other categories. | *"The survey was so long, I couldn't stay focused."*<br>*"It was especially cold last week, so my child didn't play outside at all, which isn't usual for us."* |
| **6. No issues** | **6. No issues** | No issues identified. | *"This question was really easy to answer."* |

Based on prior studies (Willis, 2004; Tourangeau, 1984), a codebook with nine categories was created, including seven from the literature and two additional ones: "Other issues" and "No issues."

**Supplementary Table S4. Allocation of participants to the seven groups**

| Group | n | Day 0 | Days 1–7 (T1) | Day 7 (T2) | Day 10 (T3) |
|---|---|---|---|---|---|
| G1 | 34 | Received the accelerometer and diary | Accelerometer monitoring and diary recording | MBQ-C open (paper) | MBQ-C open (paper) |
| G2 | 34 | | | MBQ-C open (electronic) | MBQ-C open (electronic) |
| G3 | 34 | | | MBQ-C closed (paper) | MBQ-C closed (paper) |
| G4 | 34 | | | MBQ-C closed (electronic) | MBQ-C closed (electronic) |
| G5 | 23 | | | MBQ-C closed (paper) | MBQ-C open (paper) |
| G6 | 23 | | | MBQ-C closed (electronic) | MBQ-C open (electronic) |
| G7 | 26 | | | MBQ-C open (paper) | MBQ-C open (electronic) |

| Analysis | n | Questionnaire data | Groups (n) |
|---|---|---|---|
| Concurrent validity (open-ended) | 94 | Day 7 | G1 (34), G2 (34), G7 (26) |
| Concurrent validity (closed-ended) | 114 | Day 7 | G3 (34), G4 (34), G5 (23), G6 (23) |
| Test–retest reliability (open-ended) | 68 | Day 7 and Day 10 | G1 (34), G2 (34) |
| Test–retest reliability (closed-ended) | 68 | Day 7 and Day 10 | G3 (34), G4 (34) |
| Comparison of response formats (open- vs closed-ended) | 46 | Day 7 and Day 10 | G5 (23), G6 (23) |
| Comparison of administration modes (paper vs electronic) | 26 | Day 7 and Day 10 | G7 (26) |

Values are the number of participants allocated to each group at the time of consent (n = 208).

T1, accelerometer-based assessment of physical activity together with diary-based recording of screen time and sleep (Days 1–7); T2, first questionnaire administration (Day 7); T3, second questionnaire administration (Day 10), using either the same questionnaire or one in which the response format or administration mode was changed. The validity analyses used the accelerometer-derived measures from T1 and the questionnaire data from T2; the other analyses used the questionnaire data from T2 and T3.

MBQ-C, Movement Behaviour Questionnaire-Child. Open-ended and closed-ended denote the response format; paper and electronic denote the administration mode.

### Supplementary Material S5. The 7-day 24-hour time-use diary

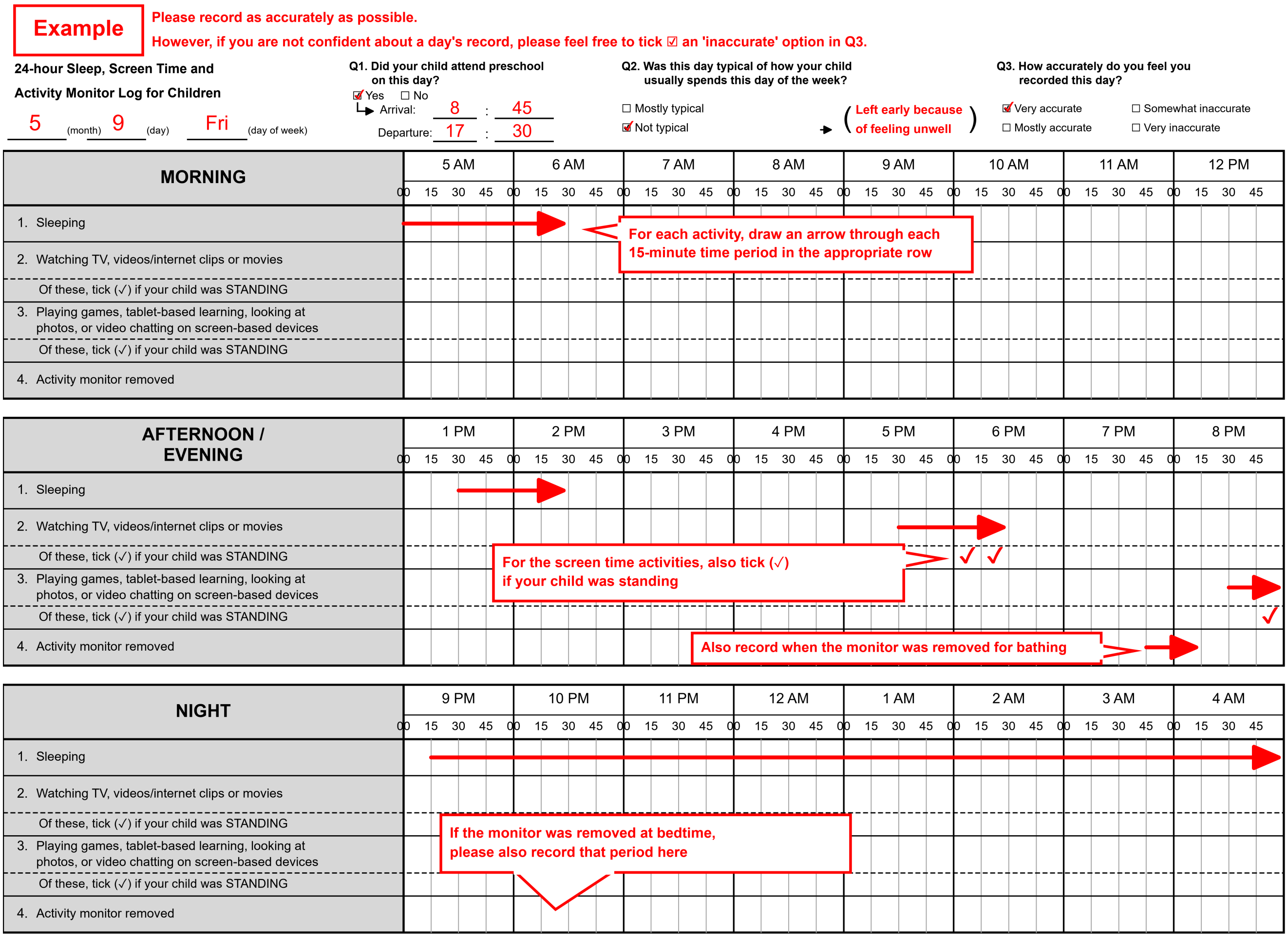

Parents completed a daily diary for seven days, recording their child's sleep, passive screen time, interactive screen time, standing time during screen use, and accelerometer non-wear time in 15-minute intervals. They also reported preschool attendance, whether the day was typical, and the perceived accuracy of the diary record. Reminder messages were sent via LINE OpenChat at 07:00 and 20:00 each day.

#### Supplementary Figure S6. Determination and correction of sleep periods

Determination and correction of sleep periods. Panel A shows the processing workflow for determining and correcting sleep periods. Panel B shows example daily 24-h timelines illustrating sleep-interval correction and the resulting epoch-level reclassification. Panel C shows the timeline legend.

##### Panel A. Processing workflow.

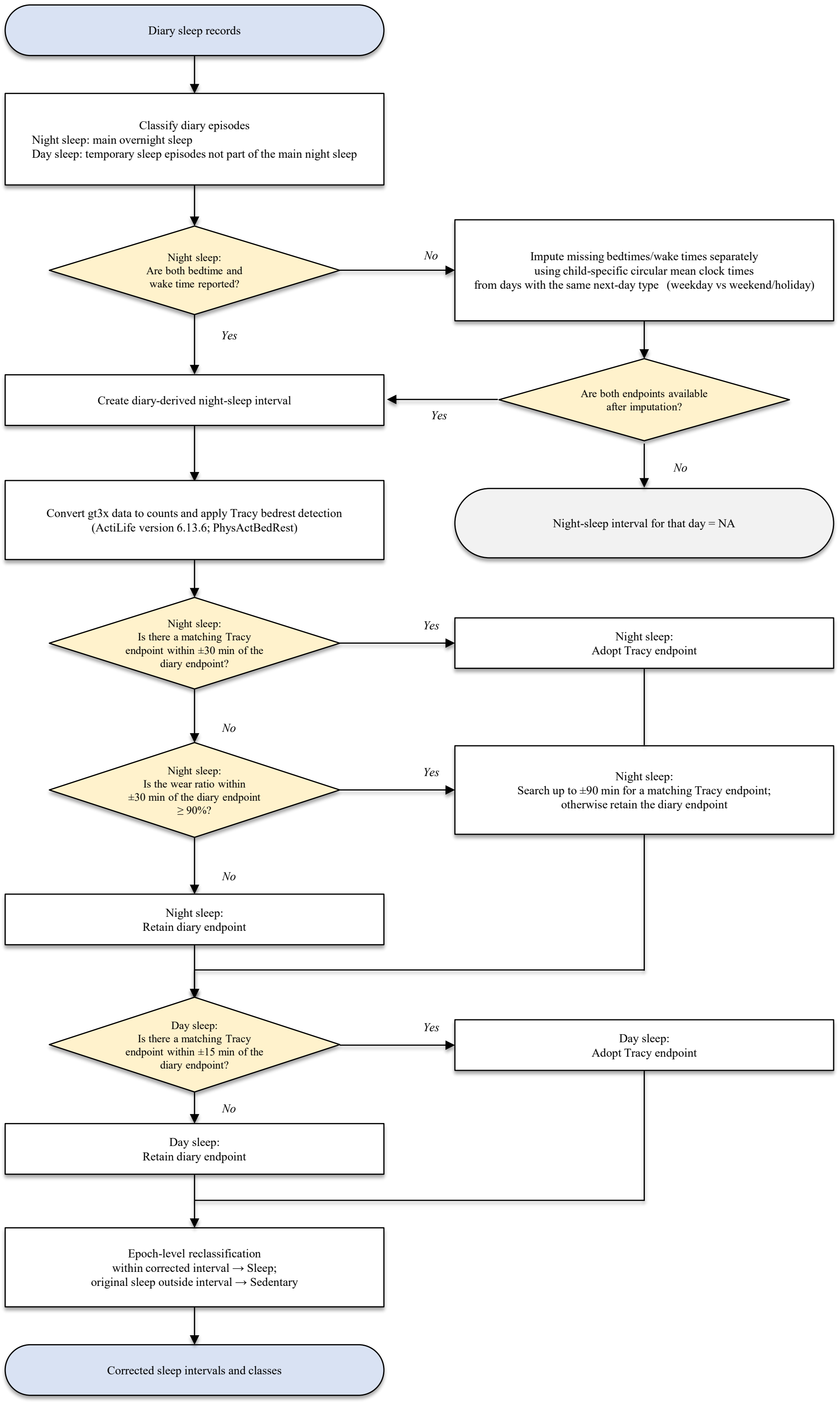

Supplementary Figure S6. Continued.

**Panel B. Sleep-interval and class correction examples.**

Each panel shows one child-day (00:00–24:00) of real participant data. Compare Sleep_Diary with Sleep_Corr to see how the sleep period was adjusted; Sleep_Tracy shows the accelerometer bedrest endpoints used for correction. Sleep_ACC shows the original actimetric sleep category, which was not used as the final sleep classification because the actimetric sleep algorithm was developed for wrist-worn accelerometers. See Panel C for the full legend.

***Pattern 1*** *— Tracy endpoint adopted within ±30 min or extended ±90-min search when local wear ratio was*

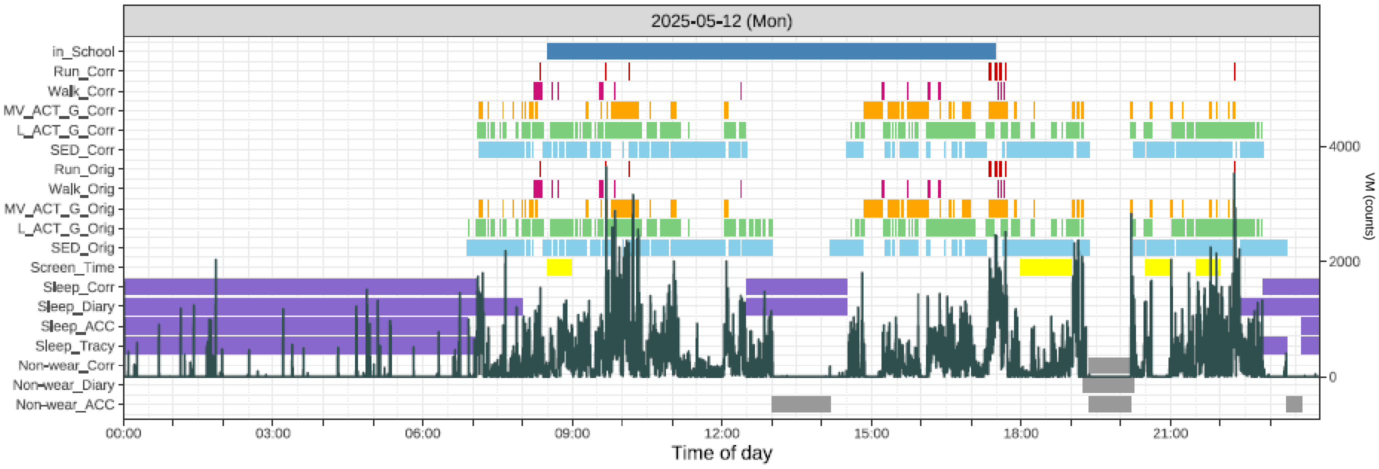

***Pattern 2*** *— Retained diary endpoint: no Tracy endpoint within ±30 min and local ACC wear <90%.*

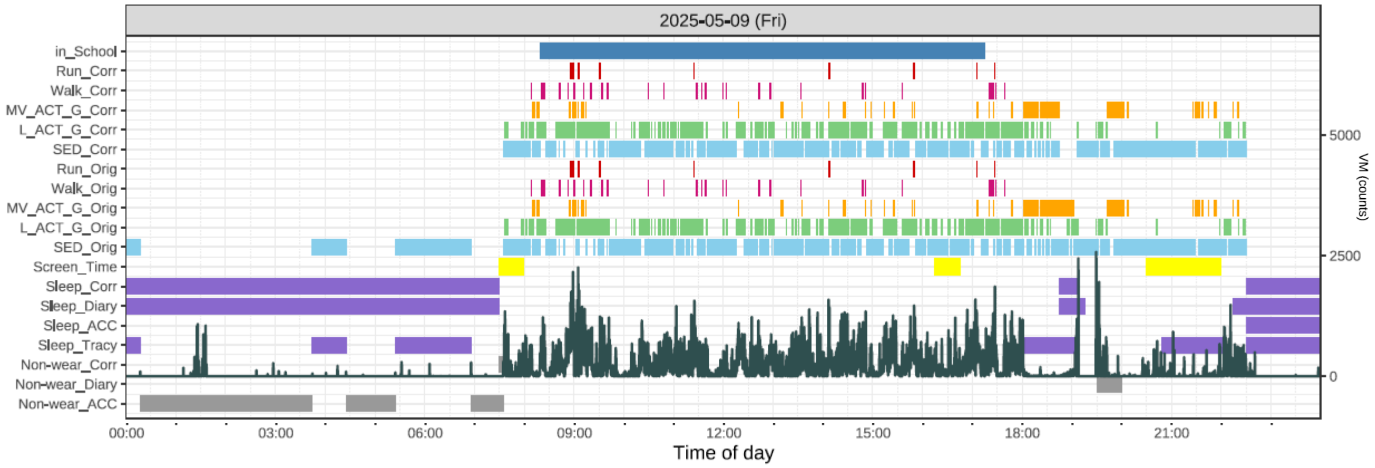

***Pattern 3*** *— Diary endpoint retained; non-wear reclassified as sleep (accelerometer not worn during sleep).*

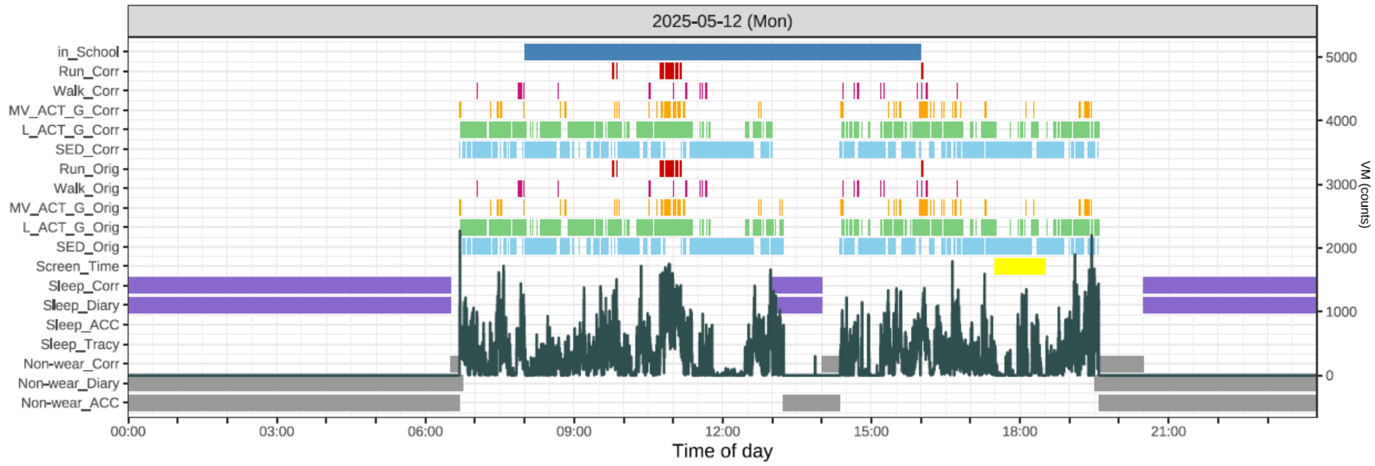

Supplementary Figure S6. Continued.

**Panel C. Legend**

Each timeline panel shows one child-day (00:00-24:00) in 15-s epochs. Rows from top to bottom: in_School; the activity classes Run, Walk, MV_ACT_G, L_ACT_G, SED (each with a "_Corr" and an "_Orig" row); Screen_Time; the sleep rows Sleep_Corr, Sleep_Diary, Sleep_ACC, Sleep_Tracy; and the non-wear rows Non-wear_Corr, Non-wear_Diary, Non-wear_ACC.

**Color and label key**

| | |
|---|---|
| **in_School** | Time at kindergarten / nursery (parent-reported) |
| **Run** | Running |
| **Walk** | Walking |
| **MV_ACT_G** | Moderate-to-vigorous activity and games |
| **L_ACT_G** | Light activity and games |
| **SED** | Sedentary |
| **Screen_Time** | Total screen time (parent-reported diary) |
| **Sleep** | Sleep interval (Corr / Diary / ACC / Tracy rows) |
| **Non-wear** | Non-wear (Corr / Diary / ACC rows) |
| **VM counts** | Vector-magnitude acceleration (background area, right axis) |

**Row meanings**

**"_Orig" rows -** Original machine-learning classification (before sleep correction).
**"_Corr" rows -** After sleep-period correction: epochs inside the corrected sleep interval are set to Sleep, and original Sleep epochs outside it are set to Sedentary.
**Sleep_Diary -** Diary-derived sleep interval.
**Sleep_Tracy -** Accelerometer bedrest endpoints (Tracy et al.; PhysActBedRest).
**Sleep_ACC -** Accelerometer-classified sleep (actimetric output; not used as the final criterion).
**Sleep_Corr -** Final corrected sleep interval (diary interval refined with Tracy endpoints).
**Non-wear_Diary / _ACC / _Corr -** Diary-reported, accelerometer-detected, and final non-wear.

##### Supplementary Figure S7. Mahalanobis distance used to identify MVPA outliers

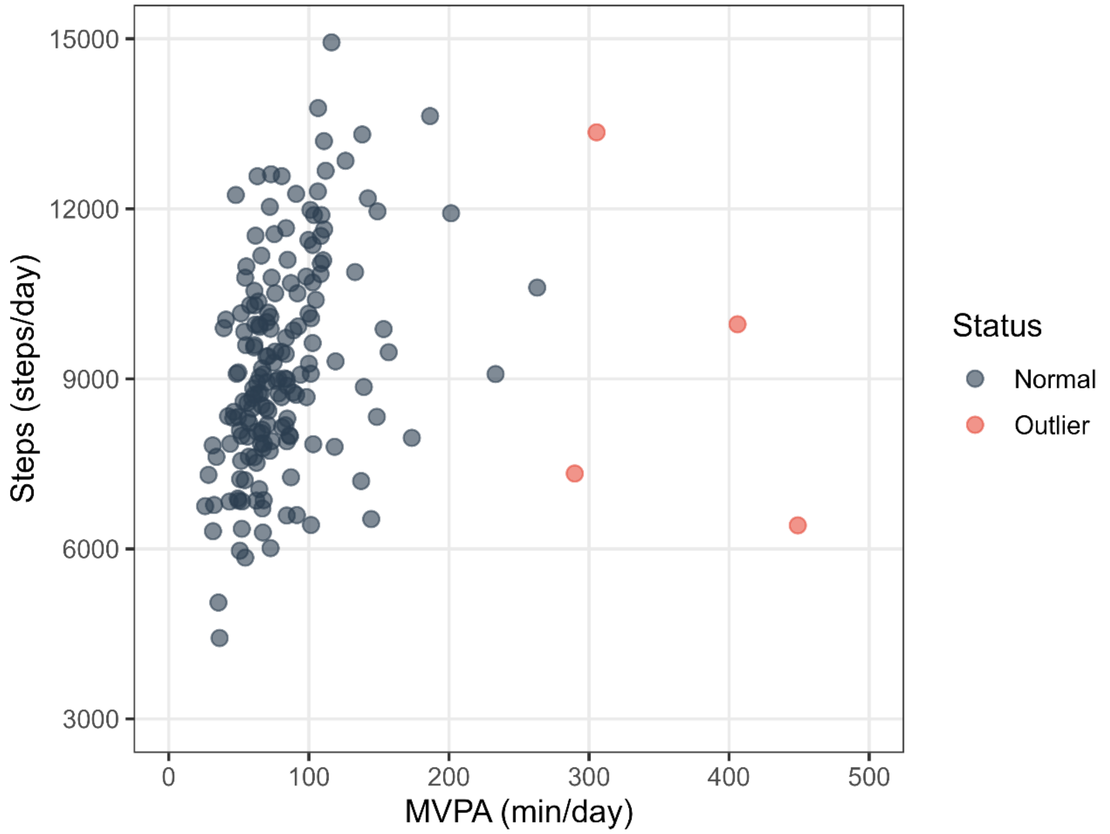

Each point represents one participant, plotted by weighted weekly mean daily moderate-to-vigorous physical activity (MVPA, min/day; x-axis) and weighted weekly mean daily step count (steps/day; y-axis). Weighted weekly means were calculated as (weekday mean × 5 + weekend mean × 2) / 7. Multivariate outliers were identified using Mahalanobis distance ($D^2$) based on MVPA and steps, with a cut-off at the 99.9th percentile of the chi-square distribution with 2 degrees of freedom ($D^2 > 13.82$). Red points indicate excluded outliers, and black points indicate participants retained for analysis. Of 175 participants screened, 4 were excluded as outliers and 171 were retained. The excluded outliers had paired values of MVPA and steps as follows: 449.0 min/day and 6,415 steps/day; 406.0 min/day and 9,963 steps/day; 305.4 min/day and 13,350 steps/day; and 289.8 min/day and 7,330 steps/day, with corresponding $D^2$ values of 51.42, 33.50, 16.85, and 16.76, respectively.

Abbreviations: MVPA, moderate-to-vigorous physical activity; $D^2$, Mahalanobis distance.

**Supplementary Figure S8. Participant flow for the cognitive interviews**

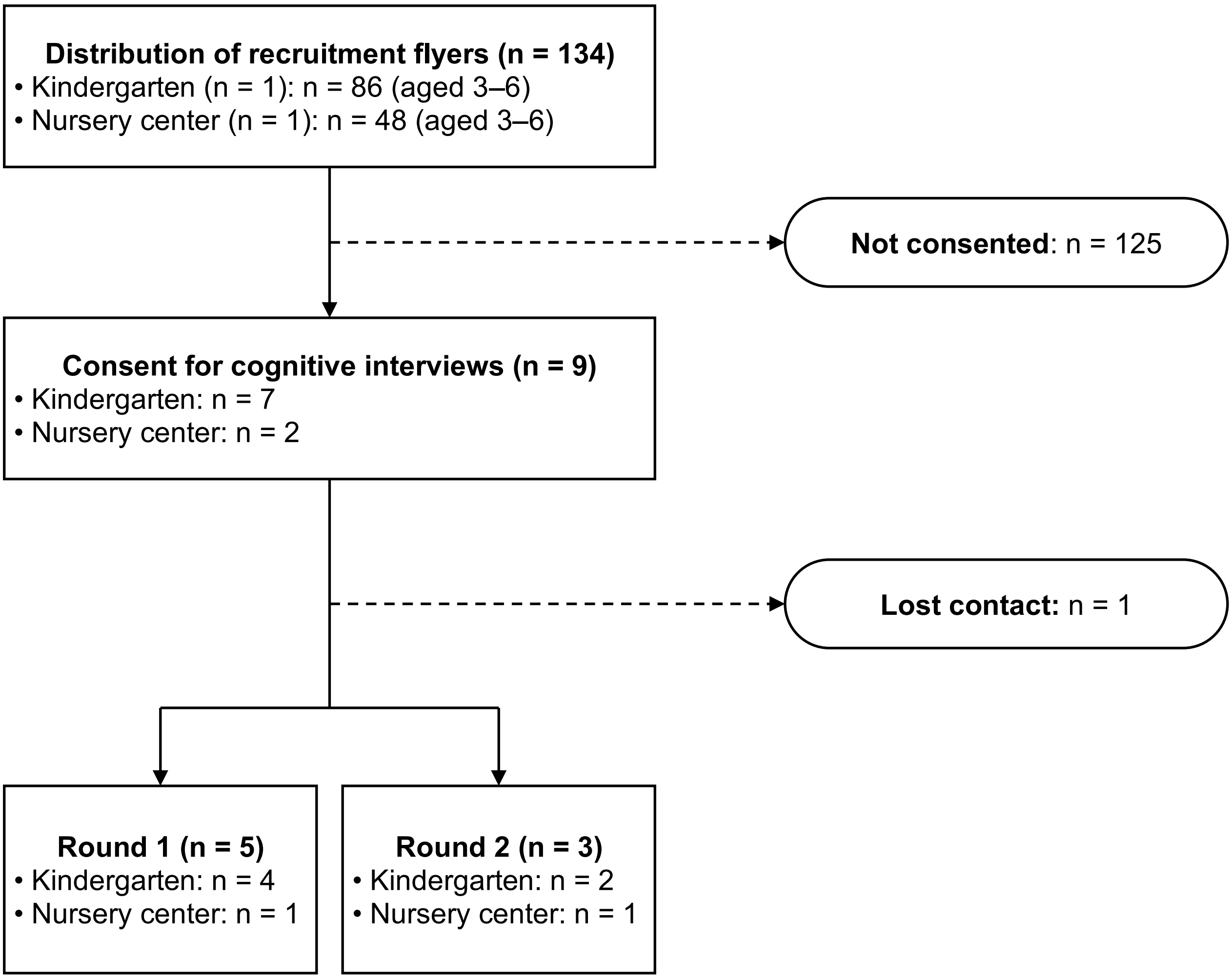

Recruitment flyers were distributed at one kindergarten and one nursery center. Of the 134 eligible children, 9 consented to the cognitive interviews. One child was subsequently lost to contact, and the remaining 8 children participated in either Round 1 (n = 5) or Round 2 (n = 3) of the cognitive interviews.

**Supplementary Table S9. Participant characteristics for cognitive interview rounds 1 and 2**

| | Round 1 (n=5) | | | | | Round 2 (n=3) | | |
|---|---|---|---|---|---|---|---|---|
| Respondent | Mother | Mother | Mother | Mother | Mother | Mother | Mother | Mother |
| Interviews duration (min) | 23.8 | 38.5 | 32.0 | 23.5 | 29.2 | 26.2 | 29.1 | 36.6 |
| Highest education | University | University | University | High school | Junior college | Vocational school | University | High school |
| Household income (yen) | 9–10 million | 5–6 million | 3–4 million | 7–8 million | 6–7 million | 7–8 million | 5–6 million | 5–6 million |
| Occupation | Part-time | Part-time | Office worker | Not working | Not working | Part-time | Self-employed | Not working |
| No. of children in household | 2 | 3 | 2 | 3 | 1 | 1 | 2 | 3 |
| Child age | 5.58 | 5.58 | 3.92 | 6.17 | 4.92 | 6.08 | 6.33 | 4.50 |
| Child gender | Girl | Girl | Girl | Girl | Boy | Girl | Girl | Girl |
| Child height (cm) | 105 | 110 | 93 | 106 | 103 | 118 | 111 | 101 |
| Child weight (kg) | 16 | 18 | 13 | 17 | 16 | 20 | 20 | 14 |
| School status | Kindergarten | Kindergarten | Nursery center | Kindergarten | Kindergarten | Kindergarten | Nursery center | Kindergarten |
| Illness/injury/disability | None | None | None | None | None | None | None | None |
| Childcare attendance (d/week) | 5 | 5 | 5 | 5 | 5 | 5 | 5 | 5 |
| Attendance time (h/d) | 6.4 | 6.4 | 9.5 | 6.2 | 6.3 | 5.5 | 7 | 6.25 |

The percentage of women aged 25 to 34 in Japan who hold a bachelor's degree or an equivalent qualification is 45% (OECD, 2024).
The average household income for households with children is 8.205 million JPY. (Ministry of Health, Labour and Welfare, 2025).
Attendance time refers to the time the child spent at kindergarten or nursery center.

#### Supplementary Table S10. Key revisions made through the cognitive interviews

| Step / phase | Main activities | COSMIN items | Key confirmations and revisions |
|---|---|---|---|
| **Step 1** Forward translation | Two independent forward translations. | 2, 3, 4, 7 | None. |
| **Step 2** Reconciliation and source-text clarification | The two forward translations were reconciled, the source-text intent was clarified with the original author, and the reconciled Japanese draft was prepared. | 8, 9 | **Confirmations with the original author**<br>**1. Specifying the target child for parents with multiple children** — Not adopted (outside MBQ scope; handled in each study).<br>**2. Age categories** — “6 years” may be added; 1- and 2-year categories may not be removed.<br>**3. Closing instruction** — Confirmed to mean “answer all questions to the best of your ability” (not “answer as many as possible”).<br>**4. Passive vs. interactive screen-time note** — Not adopted; only existing examples used.<br>**5. Scope of “game”** — Whether alone or with others not specified; interpret broadly.<br>**6. Video-chat examples** — LINE may be added and Skype etc. removed for the Japanese context.<br>**7. “nap”/“bath” wording** — “nap” intentionally not used; “bath” is one example, changeable (e.g., “bath and story”). |
| **Step 3** Back translation and equivalence review | The reconciled Japanese draft was back-translated, and the source version and back-translation were reviewed to identify discrepancies requiring revision. | 2, 5, 6, 7, 8, 9 | The source version and back-translation were reviewed by the review team. Issues requiring confirmation by the original author or revision were carried forward to Step 4. |
| **Step 4** Author-review revision | The Japanese version was revised based on the original author’s comments. The revised wording was back-translated and confirmed by the original author. | 5, 6, 7, 8, 9 | **1. Japanese rendering of “how much (time)”** — Back-translation read as “approximately how much time”; “approximately” was removed from the back-translation and the Japanese “どのくらい” was changed to “どれだけ” (“how much” without the sense of “about”). |
| **Step 5** Pilot testing — cognitive interview (Round 1) & revision | Cognitive interview (Round 1, n = 5); revise; back-translate; author review. | 5, 6, 7, 8, 9, 11, 12 | **1. Translation of “movement behaviours”** — Back-translation “daily activities” (1日の行動) inappropriate; revised to “1日の生活行動” (author approved).<br>**2. Note to include physical activity at preschool** — Added within the question (Q1A), not the opening directions.<br>**3. “Tablet-based learning” example; “looking at photos”** — Added “tablet learning” to interactive screen-time examples (Q5, Q6); “写真の閲覧” → “写真を見る.”<br>**4. Interpretation of “walk” in active play** — No note added on whether everyday walking is included.<br>**5. Definition of “a typical day”** — No note added to report an average/most-frequent value; parents conceptualize a single typical day. |
| **Step 6** Pilot testing — cognitive interview (Round 2) & revision | Cognitive interview (Round 2, n = 3); revise; back-translate; author review. | 5, 6, 7, 8, 9, 11, 12 | **1. Visual emphasis of “a typical day”** — “A typical weekday/weekend day” emphasized with brackets (「」) and bold (author approved).<br>**2. Response unit for Q9 (times/week → days/week)** — “times/week” unnatural in Japanese; author recommended “nights/week,” but “夜/週” unnatural, so changed to “日/週 (days/week).” |

Step numbers correspond to the steps shown in Figure 1. Numbers in the “COSMIN items” column refer to the item numbers in the Translation process design requirements of the COSMIN Study Design Checklist for Patient-reported outcome measurement instruments. COSMIN items 1 and 10 are addressed by the description of the source and target languages in the manuscript and by this table, which summarises the feedback report of the translation process. Abbreviations: MBQ-C, Movement Behaviour Questionnaire-Child; COSMIN, COnsensus-based Standards for the selection of health Measurement INstruments; Q, question item of the questionnaire.

**Supplementary Material S11. Japanese version of the MBQ-C, Open Version**

## 説明

この調査では、お子様の普段の 1 日の生活行動（身体活動、スクリーンタイム、睡眠）についておたずねします。
普段の 1 日とは、お子様がいつもしていることをする日です。
お子様がこれらの行動をどれだけの時間行っているかについての質問には、時間と分の両方を回答してください。例：2 時間 0 分、0 時間 30 分など。
すべての質問にできる限りご回答ください。

**お子様は何歳ですか？**

[1] 1 歳　[2] 2 歳　[3] 3 歳　[4] 4 歳　[5] 5 歳　[6] 6 歳

#### ここでは、お子様の身体活動についておたずねします

**質問 1A.** 過去 1 週間を振り返ってください。「普段の**平日の 1 日**」に、お子様は体を動かす遊びをどれだけの時間行いましたか？
体を動かす遊びとは、歩く、走る、ダンス、登る、ボールで遊ぶ、自転車やキックボードに乗る、泳ぐなどの活動を含みます。お子様が普段、幼稚園や保育園に通っている場合は、そこでの体を動かす遊びも含めてお答えください。あなたが把握している範囲でかまいません。

[　　]時間[　　]分　　**0 時間 0 分と回答した方 → 質問 2A へ**

**質問 1B.** この時間のうち、強度の高い活動（走る、ジャンプ、ダンス、自転車やキックボードに乗る、など）はどれだけの時間行いましたか？
ただし、ここで回答する時間は、前の回答よりも短くなるようにしてください。

[　　]時間[　　]分

**質問 2A.** 過去 1 週間を振り返ってください。「普段の**週末の 1 日**」に、お子様は体を動かす遊びをどれだけの時間行いましたか？
体を動かす遊びとは、歩く、走る、ダンス、登る、ボールで遊ぶ、自転車やキックボードに乗る、泳ぐなどの活動を含みます。

[　　]時間[　　]分　　**0 時間 0 分と回答した方 → 質問 3A へ**

**質問 2B.** この時間のうち、強度の高い活動（走る、ジャンプ、ダンス、自転車やキックボードに乗る、など）はどれだけの時間行いましたか？
ただし、ここで回答する時間は、前の回答よりも短くなるようにしてください。

[　　]時間[　　]分

#### ここでは、お子様のスクリーンタイムについておたずねします

**質問 3A.** 過去 1 週間を振り返ってください。「普段の**平日の 1 日**」に、お子様は**テレビ番組やビデオ、インターネット動画、映画**を、テレビやパソコン、iPad、タブレット、スマートフォンなどでどれだけの時間見ましたか？

[ ]時間[ ]分　　**0 時間 0 分と回答した方 → 質問 4A へ**

**質問 3B.** この時間のうち、テレビ番組やビデオ、インターネット動画、映画を、立って見ていた時間はどれだけありましたか？
ただし、ここで回答する時間は、前の回答よりも短くなるようにしてください。

[ ]時間[ ]分

**質問 4A.** 過去 1 週間を振り返ってください。「普段の**週末の 1 日**」に、お子様は**テレビ番組やビデオ、インターネット動画、映画**を、テレビやパソコン、iPad、タブレット、スマートフォンなどでどれだけの時間見ましたか？

[ ]時間[ ]分　　**0 時間 0 分と回答した方 → 質問 5A へ**

**質問 4B.** この時間のうち、テレビ番組やビデオ、インターネット動画、映画を、立って見ていた時間はどれだけありましたか？
ただし、ここで回答する時間は、前の回答よりも短くなるようにしてください。

[ ]時間[ ]分

**質問 5A.** 過去 1 週間を振り返ってください。「普段の**平日の 1 日**」に、お子様は**ゲーム、タブレット学習、写真を見る、ビデオ通話（LINE、Zoom など）**を、コンピューターやノートパソコン、ビデオゲーム機、iPad、タブレット、スマートフォンなどの画面付きの機器でどれだけの時間していましたか？

[ ]時間[ ]分　　**0 時間 0 分と回答した方 → 質問 6A へ**

**質問 5B.** この時間のうち、ゲーム、タブレット学習、写真を見る、ビデオ通話（LINE、Zoom など）を、立ってしていた時間はどれだけありましたか？
ただし、ここで回答する時間は、前の回答よりも短くなるようにしてください。

[ ]時間[ ]分

**質問 6A.** 過去 1 週間を振り返ってください。「普段の**週末の 1 日**」に、お子様は**ゲーム、タブレット学習、写真を見る、ビデオ通話（LINE、Zoom など）**を、コンピューターやノートパソコン、ビデオゲーム機、iPad、タブレット、スマートフォンなどの画面付きの機器でどれだけの時間していましたか？

[ ]時間[ ]分　　**0 時間 0 分と回答した方 → 質問 7 へ**

**質問 6B.** この時間のうち、ゲーム、タブレット学習、写真を見る、ビデオ通話（LINE、Zoom など）を、立ってしていた時間はどれだけありましたか？
ただし、ここで回答する時間は、前の回答よりも短くなるようにしてください。

[ ]時間[ ]分

#### ここでは、お子様の睡眠についておたずねします

**質問 7.** 過去 1 週間を振り返ってください。普段、**夜間に**、お子様は 1 日あたりどれだけの時間寝ていましたか？

[ ]時間[ ]分

**質問 8.** 過去 1 週間を振り返ってください。普段、**日中に**、お子様は 1 日あたりどれだけの時間寝ていましたか？

[ ]時間[ ]分

**質問 9.** 普段の 1 週間を振り返ってください。お子様はどれだけの頻度で、決まった寝る前の習慣（お風呂に入って読み聞かせをしてから寝るなど）を行いますか？

[ ]日/週

ご回答、ありがとうございました。

Note: Items 質問 1A to 質問 9 in this Japanese version correspond one-to-one to items Q1A to Q9 of the original English MBQ-C (Open Version), which is publicly available at: https://research.qut.edu.au/peng/wp-content/uploads/sites/85/2022/05/MBQ-Child-Open_hard-copy-survey-and-calculation-of-outcome-variables_v1_25.05.22.pdf

**Supplementary Material S12. Japanese version of the MBQ-C, Closed-ended Version**

## 説明

この調査では、お子様の普段の 1 日の生活行動（身体活動、スクリーンタイム、睡眠）についておたずねします。
普段の 1 日とは、お子様がいつもしていることをする日です。
すべての質問にできる限りご回答ください。

**お子様は何歳ですか？**

[1] 1 歳　[2] 2 歳　[3] 3 歳　[4] 4 歳　[5] 5 歳　[6] 6 歳

#### ここでは、お子様の身体活動についておたずねします

**質問 1A.** 過去 1 週間を振り返ってください。「普段の**平日の 1 日**」に、お子様は体を動かす遊びをどれだけの時間行いましたか？
体を動かす遊びとは、歩く、走る、ダンス、登る、ボールで遊ぶ、自転車やキックボードに乗る、泳ぐなどの活動を含みます。お子様が普段、幼稚園や保育園に通っている場合は、そこでの体を動かす遊びも含めてお答えください。あなたが把握している範囲でかまいません。

[0] 0 分/日　→ **質問 2A へ**
[1] 1 分以上 30 分未満/日
[2] 30 分以上 60 分未満/日
[3] 1 時間以上 2 時間未満/日
[4] 2 時間以上 3 時間未満/日
[5] 3 時間以上 4 時間未満/日
[6] 4 時間以上/日

**質問 1B.** この時間のうち、強度の高い活動（走る、ジャンプ、ダンス、自転車やキックボードに乗る、など）はどれだけの時間行いましたか？
ただし、ここで回答する時間は、前の回答よりも短くなるようにしてください。

[0] 0 分/日
[1] 1 分以上 15 分未満/日
[2] 15 分以上 30 分未満/日
[3] 30 分以上 60 分未満/日
[4] 1 時間以上 1 時間 30 分未満/日
[5] 1 時間 30 分以上 2 時間未満/日
[6] 2 時間以上/日

**質問 2A.** 過去 1 週間を振り返ってください。「普段の**週末の 1 日**」に、お子様は体を動かす遊びをどれだけの時間行いましたか？
体を動かす遊びとは、歩く、走る、ダンス、登る、ボールで遊ぶ、自転車やキックボードに乗る、泳ぐなどの活動を含みます。

[0] 0 分/日　→ **質問 3A へ**
[1] 1 分以上 30 分未満/日
[2] 30 分以上 60 分未満/日
[3] 1 時間以上 2 時間未満/日
[4] 2 時間以上 3 時間未満/日
[5] 3 時間以上 4 時間未満/日
[6] 4 時間以上/日

**質問 2B.** この時間のうち、強度の高い活動（走る、ジャンプ、ダンス、自転車やキックボードに乗る、など）はどれだけの時間行いましたか？
ただし、ここで回答する時間は、前の回答よりも短くなるようにしてください。

[0] 0 分/日
[1] 1 分以上 15 分未満/日
[2] 15 分以上 30 分未満/日
[3] 30 分以上 60 分未満/日
[4] 1 時間以上 1 時間 30 分未満/日
[5] 1 時間 30 分以上 2 時間未満/日
[6] 2 時間以上/日

#### ここでは、お子様のスクリーンタイムについておたずねします

**質問 3A.** 過去 1 週間を振り返ってください。「普段の**平日の 1 日**」に、お子様は**テレビ番組やビデオ、インターネット動画、映画**を、テレビやパソコン、iPad、タブレット、スマートフォンなどでどれだけの時間見ましたか？

[0] 0 分/日 → **質問 4A へ**
[1] 1 分以上 15 分未満/日
[2] 15 分以上 30 分未満/日
[3] 30 分以上 60 分未満/日
[4] 1 時間以上 1 時間 30 分未満/日
[5] 1 時間 30 分以上 2 時間未満/日
[6] 2 時間以上 3 時間未満/日
[7] 3 時間以上/日

**質問 3B.** この時間のうち、テレビ番組やビデオ、インターネット動画、映画を、立って見ていた時間はどれだけありましたか？
ただし、ここで回答する時間は、前の回答よりも短くなるようにしてください。

[0] 0 分/日
[1] 1 分以上 15 分未満/日
[2] 15 分以上 30 分未満/日
[3] 30 分以上 60 分未満/日
[4] 1 時間以上 1 時間 30 分未満/日
[5] 1 時間 30 分以上 2 時間未満/日
[6] 2 時間以上 3 時間未満/日
[7] 3 時間以上/日

**質問 4A.** 過去 1 週間を振り返ってください。「普段の**週末の 1 日**」に、お子様は**テレビ番組やビデオ、インターネット動画、映画**を、テレビやパソコン、iPad、タブレット、スマートフォンなどでどれだけの時間見ましたか？

[0] 0 分/日 → **質問 5A へ**
[1] 1 分以上 15 分未満/日
[2] 15 分以上 30 分未満/日
[3] 30 分以上 60 分未満/日
[4] 1 時間以上 1 時間 30 分未満/日
[5] 1 時間 30 分以上 2 時間未満/日
[6] 2 時間以上 3 時間未満/日
[7] 3 時間以上/日

**質問 4B.** この時間のうち、テレビ番組やビデオ、インターネット動画、映画を、立って見ていた時間はどれだけありましたか？
ただし、ここで回答する時間は、前の回答よりも短くなるようにしてください。

[0] 0 分/日
[1] 1 分以上 15 分未満/日
[2] 15 分以上 30 分未満/日
[3] 30 分以上 60 分未満/日
[4] 1 時間以上 1 時間 30 分未満/日
[5] 1 時間 30 分以上 2 時間未満/日
[6] 2 時間以上 3 時間未満/日
[7] 3 時間以上/日

**質問 5A.** 過去 1 週間を振り返ってください。「普段の**平日の 1 日**」に、お子様は**ゲーム、タブレット学習、写真を見る、ビデオ通話（LINE、Zoom など）**を、コンピューターやノートパソコン、ビデオゲーム機、iPad、タブレット、スマートフォンなどの画面付きの機器でどれだけの時間していましたか？

[0] 0 分/日 → **質問 6A へ**
[1] 1 分以上 15 分未満/日
[2] 15 分以上 30 分未満/日
[3] 30 分以上 60 分未満/日
[4] 1 時間以上 1 時間 30 分未満/日
[5] 1 時間 30 分以上 2 時間未満/日
[6] 2 時間以上 3 時間未満/日
[7] 3 時間以上/日

**質問 5B.** この時間のうち、ゲーム、タブレット学習、写真を見る、ビデオ通話（LINE、Zoom など）を、立ってしていた時間はどれだけありましたか？
ただし、ここで回答する時間は、前の回答よりも短くなるようにしてください。

[0] 0 分/日
[1] 1 分以上 15 分未満/日
[2] 15 分以上 30 分未満/日
[3] 30 分以上 60 分未満/日
[4] 1 時間以上 1 時間 30 分未満/日
[5] 1 時間 30 分以上 2 時間未満/日
[6] 2 時間以上 3 時間未満/日
[7] 3 時間以上/日

**質問 6A.** 過去 1 週間を振り返ってください。「普段の**週末の 1 日**」に、お子様は**ゲーム、タブレット学習、写真を見る、ビデオ通話（LINE、Zoom など）**を、コンピューターやノートパソコン、ビデオゲーム機、iPad、タブレット、スマートフォンなどの画面付きの機器でどれだけの時間していましたか？

[0] 0 分/日　→ **質問 7 へ**
[1] 1 分以上 15 分未満/日
[2] 15 分以上 30 分未満/日
[3] 30 分以上 60 分未満/日
[4] 1 時間以上 1 時間 30 分未満/日
[5] 1 時間 30 分以上 2 時間未満/日
[6] 2 時間以上 3 時間未満/日
[7] 3 時間以上/日

**質問 6B.** この時間のうち、ゲーム、タブレット学習、写真を見る、ビデオ通話（LINE、Zoom など）を、立ってしていた時間はどれだけありましたか？
ただし、ここで回答する時間は、前の回答よりも短くなるようにしてください。

[0] 0 分/日
[1] 1 分以上 15 分未満/日
[2] 15 分以上 30 分未満/日
[3] 30 分以上 60 分未満/日
[4] 1 時間以上 1 時間 30 分未満/日
[5] 1 時間 30 分以上 2 時間未満/日
[6] 2 時間以上 3 時間未満/日
[7] 3 時間以上/日

#### ここでは、お子様の睡眠についておたずねします

**質問 7.** 過去 1 週間を振り返ってください。普段、**夜間に**、お子様は 1 日あたりどれだけの時間寝ていましたか？

[1] 6 時間未満/日
[2] 6 時間以上 8 時間未満/日
[3] 8 時間以上 10 時間未満/日
[4] 10 時間以上 12 時間未満/日
[5] 12 時間以上 14 時間未満/日
[6] 14 時間以上/日

**質問 8.** 過去 1 週間を振り返ってください。普段、**日中に**、お子様は 1 日あたりどれだけの時間寝ていましたか？

[0] 0 時間/日
[1] 1 時間未満/日
[2] 1 時間以上 2 時間未満/日
[3] 2 時間以上 3 時間未満/日
[4] 3 時間以上 4 時間未満/日
[5] 4 時間以上/日

**質問 9.** 普段の 1 週間を振り返ってください。お子様はどれだけの頻度で、決まった寝る前の習慣（お風呂に入って読み聞かせをしてから寝るなど）を行いますか？

[0] 全くしない
[1] 週に 1～2 日
[2] 週に 3～4 日
[3] 週に 5～6 日
[4] 毎日

ご回答、ありがとうございました。

Note: Items 質問 1A to 質問 9 in this Japanese version correspond one-to-one to items Q1A to Q9 of the original English MBQ-C (Closed Version), which is publicly available at: https://research.qut.edu.au/peng/wp-content/uploads/sites/85/2022/05/MBQ-Child-Closed_hard-copy-survey-and-calculation-of-outcome-variables_v1_25.05.22.pdf

#### Supplementary Figure S13. Participant flow for the validation study

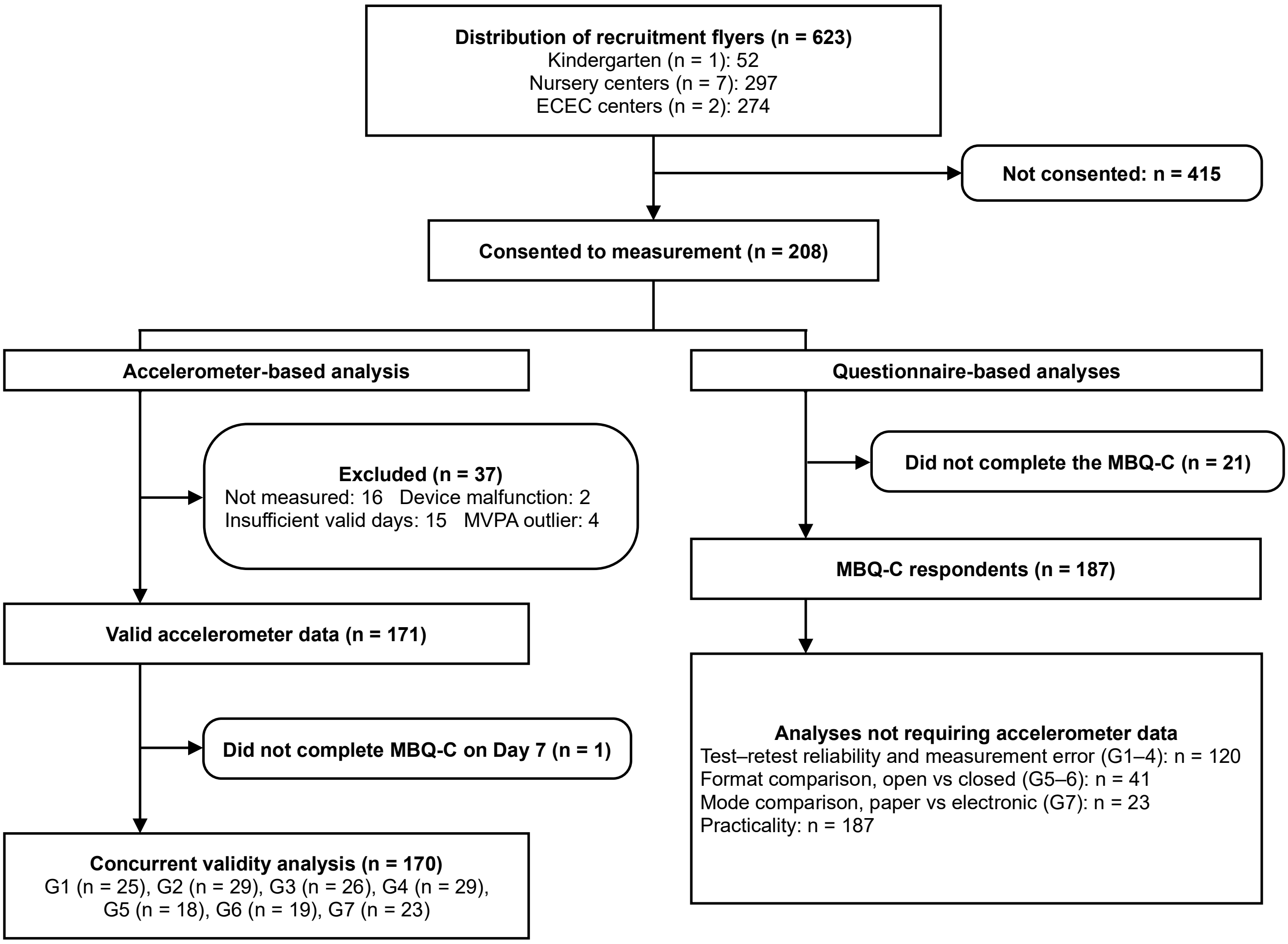

Of the 623 eligible children, 208 consented. From this point the study followed two overlapping streams. In the accelerometer-based stream, 37 children were excluded, leaving 171 with valid accelerometer data; after excluding one child who did not complete the MBQ-C on Day 7, 170 were included in the concurrent-validity analysis. In the questionnaire-based stream, which does not require accelerometer data, 187 children completed the MBQ-C and were used for test–retest reliability and measurement error, format and mode comparisons, and practicality. Practicality (completion time and burden, n = 186; clarity, n = 187): one respondent did not complete the time/burden items on Day 7. The two streams overlap: 170 of the respondents also had valid accelerometer data. The group breakdown in the concurrent-validity box (n = 169) excludes one child who deviated from the allocated protocol (open-ended paper version on Day 7 and closed-ended paper version on Day 10); this child was analysed according to the versions actually completed and is included in the open-ended paper counts (Table 2) and in the format comparison (n = 41). ECEC, early childhood education and care; MVPA, moderate-to-vigorous physical activity.

**Supplementary Table S14. Characteristics of the participants included in each analysis**

| Characteristic | Concurrent validity (n = 170) | Test–retest reliability and measurement error (n = 120) | Format comparison (open vs closed) (n = 41) | Mode comparison (paper vs electronic) (n = 23) | Practicality (n = 187) |
|---|---|---|---|---|---|
| Child age (yr), mean (SD) | 4.95 (0.84) | 4.94 (0.86) | 4.85 (0.75) | 4.98 (0.93) | 4.94 (0.85) |
| Age category, n (%) | | | | | |
| 3 yr | 30 (17.6) | 24 (20.0) | 7 (17.1) | 4 (17.4) | 35 (18.7) |
| 4 yr | 57 (33.5) | 38 (31.7) | 16 (39.0) | 8 (34.8) | 62 (33.2) |
| 5 yr | 65 (38.2) | 43 (35.8) | 17 (41.5) | 8 (34.8) | 70 (37.4) |
| 6 yr | 18 (10.6) | 15 (12.5) | 1 (2.4) | 3 (13.0) | 20 (10.7) |
| Sex, n (%) | | | | | |
| Boy | 80 (47.1) | 55 (46.2) | 22 (53.7) | 10 (43.5) | 88 (47.3) |
| Girl | 90 (52.9) | 64 (53.8) | 19 (46.3) | 13 (56.5) | 98 (52.7) |
| Missing | 0 | 1 | 0 | 0 | 1 |
| Type of preschool, n (%) | | | | | |
| Kindergarten | 5 (2.9) | 3 (2.5) | 2 (4.9) | 0 (0.0) | 5 (2.7) |
| Nursery center | 61 (35.9) | 50 (41.7) | 8 (19.5) | 6 (26.1) | 65 (34.8) |
| ECEC center | 104 (61.2) | 67 (55.8) | 31 (75.6) | 17 (73.9) | 117 (62.6) |
| Respondent, n (%) | | | | | |
| Mother | 151 (89.3) | 104 (88.9) | 36 (87.8) | 22 (95.7) | 163 (88.6) |
| Father | 16 (9.5) | 12 (10.3) | 4 (9.8) | 1 (4.3) | 19 (10.3) |
| Grandparent | 2 (1.2) | 1 (0.9) | 1 (2.4) | 0 (0.0) | 2 (1.1) |
| Missing | 1 | 3 | 0 | 0 | 3 |
| Respondent age (yr), mean (SD) | 37.3 (5.8) | 37.0 (6.0) | 37.9 (6.2) | 37.1 (3.4) | 37.2 (5.7) |
| Parent respondent education, n (%) | | | | | |
| Junior high or high school | 33 (20.6) | 27 (23.9) | 6 (16.2) | 2 (9.1) | 38 (21.7) |
| Technical/junior college or vocational school | 49 (30.6) | 37 (32.7) | 15 (40.5) | 3 (13.6) | 55 (31.4) |
| University or graduate school | 78 (48.8) | 49 (43.4) | 16 (43.2) | 17 (77.3) | 82 (46.9) |
| Missing | 10 | 7 | 4 | 1 | 12 |
| Household income (million JPY), n (%) | | | | | |
| <3 | 5 (4.3) | 5 (6.0) | 0 (0.0) | 0 (0.0) | 5 (3.9) |
| 3–<5 | 17 (14.5) | 12 (14.3) | 7 (26.9) | 1 (5.9) | 21 (16.3) |
| 5–<8 | 44 (37.6) | 36 (42.9) | 11 (42.3) | 4 (23.5) | 52 (40.3) |
| 8–<10 | 27 (23.1) | 14 (16.7) | 6 (23.1) | 7 (41.2) | 27 (20.9) |
| ≥10 | 24 (20.5) | 17 (20.2) | 2 (7.7) | 5 (29.4) | 24 (18.6) |
| Missing | 53 | 36 | 15 | 6 | 58 |

Values are mean (SD) for continuous variables and n (%) for categorical variables; percentages exclude missing values. Samples overlap (e.g., the concurrent-validity sample is largely a subset of the questionnaire respondents). Concurrent validity required valid accelerometer data; the other analyses did not. ECEC, early childhood education and care.

#### Supplementary Table S15. Detailed validity and reliability by indicator and day type

**Part A.** Concurrent validity (Spearman ρ), by response format and mode

Open-Overall

| Period | Criterion measure | MBQ-C item | n | ρ [95% CI] | Criterion Med (Q1-Q3) | MBQ-C Med (Q1-Q3) | P |
|---|---|---|---|---|---|---|---|
| Weekday | Total PA (ACC) | Active play | 78 | 0.09 [-0.131, 0.31] | 403.2 (359.0-441.4) | 150.0 (90.0-240.0) | NA |
| | MVPA (ACC) | Energetic play | 78 | 0.20 [-0.025, 0.404] | 65.2 (53.5-89.2) | 60.0 (30.0-60.0) | NA |
| | Total screen time (diary) | Total screen time | 78 | **0.87 [0.805, 0.916]** | 94.9 (37.5-170.3) | 120.0 (60.0-180.0) | <0.001 |
| | Passive screen (diary) | Passive screen | 78 | **0.83 [0.745, 0.888]** | 78.8 (36.8-129.0) | 114.0 (60.0-120.0) | 0.020 |
| | Interactive screen (diary) | Interactive screen | 78 | **0.53 [0.352, 0.675]** | 1.5 (0.0-22.5) | 0.0 (0.0-30.0) | 0.009 |
| | Sedentary screen (diary) | Sedentary screen | 78 | **0.82 [0.724, 0.879]** | 93.4 (36.0-157.5) | 105.0 (60.0-150.0) | 0.020 |
| | Night sleep (diary+ACC) | Night sleep | 78 | **0.64 [0.48, 0.751]** | 585.1 (566.7-607.3) | 570.0 (540.0-600.0) | 0.039 |
| | Day sleep (diary+ACC) | Day sleep | 78 | **0.59 [0.42, 0.717]** | 0.0 (0.0-15.0) | 0.0 (0.0-30.0) | 0.018 |
| Weekend | Total PA (ACC) | Active play | 78 | 0.22 [-0.003, 0.422] | 387.4 (339.4-425.1) | 120.0 (60.0-180.0) | NA |
| | MVPA (ACC) | Energetic play | 78 | **0.27 [0.053, 0.467]** | 76.3 (54.9-100.1) | 60.0 (30.0-90.0) | NA |
| | Total screen time (diary) | Total screen time | 78 | **0.64 [0.484, 0.753]** | 141.3 (84.4-208.8) | 150.0 (90.0-240.0) | 0.061 |
| | Passive screen (diary) | Passive screen | 78 | **0.58 [0.414, 0.713]** | 120.0 (70.0-189.4) | 120.0 (62.5-180.0) | 0.257 |
| | Interactive screen (diary) | Interactive screen | 78 | **0.48 [0.287, 0.634]** | 0.0 (0.0-30.0) | 0.0 (0.0-37.5) | 0.605 |
| | Sedentary screen (diary) | Sedentary screen | 78 | **0.63 [0.473, 0.747]** | 122.5 (82.5-193.1) | 125.0 (82.5-210.0) | 0.504 |
| | Night sleep (diary+ACC) | Night sleep | 78 | **0.28 [0.057, 0.47]** | 594.0 (574.3-618.0) | 570.0 (540.0-600.0) | 0.008 |
| | Day sleep (diary+ACC) | Day sleep | 78 | **0.34 [0.13, 0.525]** | 12.5 (0.0-37.0) | 0.0 (0.0-30.0) | 0.615 |
| Weekly | Total PA (ACC) | Active play | 78 | 0.15 [-0.079, 0.357] | 392.6 (352.5-439.2) | 162.9 (92.1-204.6) | NA |
| | MVPA (ACC) | Energetic play | 78 | **0.22 [0.001, 0.425]** | 66.7 (56.3-91.5) | 51.4 (28.0-77.1) | NA |
| | Total screen time (diary) | Total screen time | 78 | **0.85 [0.771, 0.901]** | 109.0 (52.0-188.5) | 124.3 (69.6-192.9) | <0.001 |
| | Passive screen (diary) | Passive screen | 78 | **0.85 [0.773, 0.901]** | 91.8 (47.8-156.0) | 106.1 (60.2-150.0) | 0.021 |
| | Interactive screen (diary) | Interactive screen | 78 | **0.53 [0.345, 0.671]** | 4.3 (0.0-20.9) | 7.9 (0.0-33.2) | 0.056 |
| | Sedentary screen (diary) | Sedentary screen | 78 | **0.81 [0.715, 0.874]** | 97.2 (49.8-178.8) | 120.0 (60.0-172.5) | 0.028 |
| | Total sleep (diary+ACC) | Total sleep | 78 | **0.58 [0.404, 0.707]** | 604.5 (580.7-622.9) | 600.0 (570.0-630.0) | 0.317 |
| | Night sleep (diary+ACC) | Night sleep | 78 | **0.56 [0.382, 0.694]** | 590.5 (570.1-610.7) | 570.0 (540.0-600.0) | 0.022 |
| | Day sleep (diary+ACC) | Day sleep | 78 | **0.60 [0.441, 0.729]** | 6.4 (0.0-22.6) | 0.0 (0.0-30.0) | 0.235 |

Open-Paper

| Period | Criterion measure | MBQ-C item | n | ρ [95% CI] | Criterion Med (Q1-Q3) | MBQ-C Med (Q1-Q3) | P |
|---|---|---|---|---|---|---|---|
| Weekday | Total PA (ACC) | Active play | 49 | 0.20 [-0.082, 0.459] | 396.7 (368.0-437.3) | 150.0 (90.0-225.0) | NA |
| | MVPA (ACC) | Energetic play | 49 | 0.18 [-0.103, 0.442] | 64.8 (55.0-85.8) | 40.0 (20.0-60.0) | NA |
| | Total screen time (diary) | Total screen time | 49 | **0.86 [0.765, 0.92]** | 97.5 (57.0-168.0) | 120.0 (60.0-180.0) | <0.001 |
| | Passive screen (diary) | Passive screen | 49 | **0.84 [0.734, 0.908]** | 82.5 (52.5-129.0) | 120.0 (60.0-150.0) | 0.015 |
| | Interactive screen (diary) | Interactive screen | 49 | **0.44 [0.184, 0.644]** | 3.8 (0.0-22.5) | 10.0 (0.0-30.0) | 0.038 |
| | Sedentary screen (diary) | Sedentary screen | 49 | **0.82 [0.702, 0.896]** | 96.0 (57.0-153.0) | 120.0 (60.0-180.0) | 0.007 |
| | Night sleep (diary+ACC) | Night sleep | 49 | **0.66 [0.467, 0.795]** | 583.8 (567.3-608.4) | 570.0 (550.0-600.0) | 0.102 |
| | Day sleep (diary+ACC) | Day sleep | 49 | **0.56 [0.332, 0.727]** | 0.0 (0.0-15.0) | 0.0 (0.0-40.0) | 0.013 |
| Weekend | Total PA (ACC) | Active play | 49 | **0.31 [0.029, 0.542]** | 384.1 (338.3-426.0) | 120.0 (60.0-180.0) | NA |
| | MVPA (ACC) | Energetic play | 49 | 0.18 [-0.103, 0.442] | 67.3 (49.3-92.1) | 45.0 (20.0-60.0) | NA |
| | Total screen time (diary) | Total screen time | 49 | **0.51 [0.265, 0.691]** | 157.5 (90.0-225.0) | 150.0 (90.0-240.0) | 0.772 |
| | Passive screen (diary) | Passive screen | 49 | **0.54 [0.3, 0.71]** | 130.0 (75.0-195.0) | 150.0 (70.0-180.0) | 0.915 |
| | Interactive screen (diary) | Interactive screen | 49 | **0.48 [0.232, 0.672]** | 0.0 (0.0-25.0) | 0.0 (0.0-60.0) | 0.534 |
| | Sedentary screen (diary) | Sedentary screen | 49 | **0.53 [0.293, 0.706]** | 145.0 (90.0-195.0) | 135.0 (90.0-220.0) | 0.907 |
| | Night sleep (diary+ACC) | Night sleep | 49 | **0.33 [0.051, 0.557]** | 594.3 (568.8-623.8) | 570.0 (550.0-600.0) | 0.080 |
| | Day sleep (diary+ACC) | Day sleep | 49 | **0.40 [0.135, 0.613]** | 7.5 (0.0-32.9) | 0.0 (0.0-40.0) | 0.596 |
| Weekly | Total PA (ACC) | Active play | 49 | 0.27 [-0.012, 0.512] | 391.2 (358.0-425.1) | 141.4 (81.4-197.1) | NA |
| | MVPA (ACC) | Energetic play | 49 | 0.17 [-0.115, 0.432] | 64.6 (56.3-84.2) | 38.6 (24.3-68.6) | NA |
| | Total screen time (diary) | Total screen time | 49 | **0.77 [0.626, 0.865]** | 122.1 (64.3-188.4) | 137.1 (98.6-192.9) | 0.005 |
| | Passive screen (diary) | Passive screen | 49 | **0.82 [0.694, 0.892]** | 103.9 (55.7-156.4) | 120.0 (68.6-150.0) | 0.051 |
| | Interactive screen (diary) | Interactive screen | 49 | **0.49 [0.243, 0.678]** | 5.5 (0.0-21.4) | 10.7 (0.0-34.3) | 0.101 |
| | Sedentary screen (diary) | Sedentary screen | 49 | **0.77 [0.625, 0.865]** | 112.7 (64.3-171.8) | 128.6 (68.6-188.6) | 0.033 |
| | Total sleep (diary+ACC) | Total sleep | 49 | **0.58 [0.352, 0.738]** | 599.4 (575.1-622.5) | 600.0 (570.0-630.0) | 0.701 |
| | Night sleep (diary+ACC) | Night sleep | 49 | **0.55 [0.322, 0.722]** | 586.8 (569.8-610.8) | 570.0 (550.0-600.0) | 0.087 |
| | Day sleep (diary+ACC) | Day sleep | 49 | **0.62 [0.405, 0.765]** | 6.2 (0.0-17.1) | 0.0 (0.0-40.0) | 0.056 |

Supplementary Table S15. Continued.

Open-Electronic

| Period | Criterion measure | MBQ-C item | n | ρ [95% CI] | Criterion Med (Q1-Q3) | MBQ-C Med (Q1-Q3) | P |
|---|---|---|---|---|---|---|---|
| Weekday | Total PA (ACC) | Active play | 29 | -0.03 [-0.391, 0.342] | 405.4 (351.1-448.2) | 180.0 (90.0-240.0) | NA |
| | MVPA (ACC) | Energetic play | 29 | 0.27 [-0.11, 0.577] | 68.4 (50.9-94.8) | 60.0 (30.0-120.0) | NA |
| | Total screen time (diary) | Total screen time | 29 | **0.84 [0.692, 0.925]** | 86.2 (22.5-171.0) | 90.0 (50.0-150.0) | 0.151 |
| | Passive screen (diary) | Passive screen | 29 | **0.80 [0.605, 0.9]** | 60.0 (22.5-123.0) | 60.0 (50.0-120.0) | 0.516 |
| | Interactive screen (diary) | Interactive screen | 29 | **0.64 [0.356, 0.815]** | 0.0 (0.0-11.2) | 0.0 (0.0-30.0) | 0.078 |
| | Sedentary screen (diary) | Sedentary screen | 29 | **0.78 [0.573, 0.89]** | 63.8 (22.5-171.0) | 75.0 (40.0-120.0) | 0.682 |
| | Night sleep (diary+ACC) | Night sleep | 29 | **0.58 [0.27, 0.78]** | 591.1 (565.4-606.0) | 570.0 (540.0-600.0) | 0.176 |
| | Day sleep (diary+ACC) | Day sleep | 29 | **0.65 [0.375, 0.822]** | 0.0 (0.0-18.0) | 0.0 (0.0-30.0) | 0.726 |
| Weekend | Total PA (ACC) | Active play | 29 | 0.07 [-0.302, 0.427] | 400.0 (341.2-423.6) | 180.0 (120.0-240.0) | NA |
| | MVPA (ACC) | Energetic play | 29 | **0.44 [0.083, 0.692]** | 91.2 (56.8-111.0) | 60.0 (40.0-120.0) | NA |
| | Total screen time (diary) | Total screen time | 29 | **0.85 [0.697, 0.926]** | 127.5 (82.5-187.5) | 120.0 (90.0-210.0) | 0.007 |
| | Passive screen (diary) | Passive screen | 29 | **0.66 [0.387, 0.827]** | 100.0 (67.5-135.0) | 120.0 (60.0-180.0) | 0.043 |
| | Interactive screen (diary) | Interactive screen | 29 | **0.39 [0.032, 0.664]** | 0.0 (0.0-37.5) | 0.0 (0.0-30.0) | 0.900 |
| | Sedentary screen (diary) | Sedentary screen | 29 | **0.78 [0.579, 0.892]** | 110.0 (75.0-185.0) | 120.0 (60.0-180.0) | 0.264 |
| | Night sleep (diary+ACC) | Night sleep | 29 | 0.20 [-0.176, 0.531] | 593.5 (581.4-610.2) | 570.0 (540.0-600.0) | 0.042 |
| | Day sleep (diary+ACC) | Day sleep | 29 | 0.23 [-0.152, 0.548] | 15.0 (0.0-40.0) | 0.0 (0.0-30.0) | 0.140 |
| Weekly | Total PA (ACC) | Active play | 29 | 0.01 [-0.358, 0.375] | 394.6 (346.0-443.8) | 167.1 (120.0-274.3) | NA |
| | MVPA (ACC) | Energetic play | 29 | 0.24 [-0.142, 0.555] | 72.1 (54.6-103.7) | 60.0 (42.9-102.9) | NA |
| | Total screen time (diary) | Total screen time | 29 | **0.91 [0.819, 0.958]** | 95.5 (45.0-195.0) | 98.6 (60.0-192.9) | 0.003 |
| | Passive screen (diary) | Passive screen | 29 | **0.87 [0.731, 0.935]** | 77.9 (36.4-110.9) | 90.0 (60.0-137.1) | 0.110 |
| | Interactive screen (diary) | Interactive screen | 29 | **0.53 [0.198, 0.749]** | 0.0 (0.0-17.1) | 2.1 (0.0-30.0) | 0.344 |
| | Sedentary screen (diary) | Sedentary screen | 29 | **0.86 [0.713, 0.93]** | 77.9 (42.1-195.0) | 90.0 (51.4-138.6) | 0.358 |
| | Total sleep (diary+ACC) | Total sleep | 29 | **0.60 [0.297, 0.791]** | 610.2 (589.4-625.7) | 600.0 (570.0-630.0) | 0.247 |
| | Night sleep (diary+ACC) | Night sleep | 29 | **0.54 [0.212, 0.755]** | 593.7 (571.7-608.6) | 570.0 (540.0-600.0) | 0.132 |
| | Day sleep (diary+ACC) | Day sleep | 29 | **0.65 [0.372, 0.821]** | 12.9 (4.3-30.0) | 0.0 (0.0-30.0) | 0.511 |

Closed-Overall

| Period | Criterion measure | MBQ-C item | n | ρ [95% CI] | Criterion Med (Q1-Q3) | MBQ-C Med (Q1-Q3) | P |
|---|---|---|---|---|---|---|---|
| Weekday | Total PA (ACC) | Active play | 92 | **0.25 [0.045, 0.43]** | 405.8 (360.8-440.3) | 150.0 (90.0-210.0) | NA |
| | MVPA (ACC) | Energetic play | 92 | 0.20 [-0.002, 0.391] | 76.4 (57.4-100.6) | 45.0 (22.5-75.0) | NA |
| | Total screen time (diary) | Total screen time | 92 | **0.77 [0.668, 0.84]** | 84.0 (37.5-136.9) | 93.8 (52.5-150.0) | <0.001 |
| | Passive screen (diary) | Passive screen | 91 | **0.72 [0.608, 0.809]** | 72.0 (30.0-117.0) | 75.0 (45.0-150.0) | 0.002 |
| | Interactive screen (diary) | Interactive screen | 91 | **0.56 [0.395, 0.684]** | 0.0 (0.0-20.2) | 7.5 (0.0-45.0) | <0.001 |
| | Sedentary screen (diary) | Sedentary screen | 91 | **0.76 [0.65, 0.831]** | 75.0 (38.2-121.5) | 82.5 (45.0-142.5) | <0.001 |
| | Night sleep (diary+ACC) | Night sleep | 92 | **0.61 [0.464, 0.725]** | 596.1 (564.2-613.3) | 540.0 (540.0-660.0) | 0.046 |
| | Day sleep (diary+ACC) | Day sleep | 92 | **0.62 [0.476, 0.732]** | 0.0 (0.0-7.5) | 0.0 (0.0-30.0) | <0.001 |
| Weekend | Total PA (ACC) | Active play | 92 | **0.24 [0.034, 0.422]** | 394.1 (327.4-444.9) | 90.0 (90.0-150.0) | NA |
| | MVPA (ACC) | Energetic play | 92 | 0.12 [-0.086, 0.318] | 83.8 (54.7-108.6) | 45.0 (22.5-75.0) | NA |
| | Total screen time (diary) | Total screen time | 92 | **0.74 [0.625, 0.817]** | 127.5 (73.8-216.2) | 127.5 (75.0-240.0) | 0.450 |
| | Passive screen (diary) | Passive screen | 92 | **0.68 [0.557, 0.779]** | 101.2 (60.0-173.1) | 105.0 (75.0-150.0) | 0.902 |
| | Interactive screen (diary) | Interactive screen | 92 | **0.66 [0.531, 0.765]** | 0.0 (0.0-38.1) | 7.5 (0.0-45.0) | 0.018 |
| | Sedentary screen (diary) | Sedentary screen | 92 | **0.73 [0.621, 0.815]** | 107.5 (65.0-186.2) | 120.0 (67.5-195.0) | 0.907 |
| | Night sleep (diary+ACC) | Night sleep | 92 | 0.20 [-0.008, 0.386] | 597.3 (571.8-627.8) | 540.0 (540.0-660.0) | 0.013 |
| | Day sleep (diary+ACC) | Day sleep | 92 | **0.51 [0.346, 0.651]** | 0.0 (0.0-30.0) | 0.0 (0.0-30.0) | 0.100 |
| Weekly | Total PA (ACC) | Active play | 92 | **0.26 [0.056, 0.44]** | 398.2 (356.2-434.8) | 132.9 (76.6-197.1) | NA |
| | MVPA (ACC) | Energetic play | 92 | 0.12 [-0.087, 0.317] | 76.4 (62.6-99.7) | 38.6 (22.5-75.0) | NA |
| | Total screen time (diary) | Total screen time | 92 | **0.78 [0.689, 0.851]** | 100.1 (46.6-158.0) | 112.5 (65.1-168.5) | <0.001 |
| | Passive screen (diary) | Passive screen | 91 | **0.76 [0.654, 0.834]** | 81.4 (42.9-132.9) | 83.6 (53.6-140.4) | 0.008 |
| | Interactive screen (diary) | Interactive screen | 91 | **0.65 [0.508, 0.752]** | 2.9 (0.0-20.9) | 11.8 (0.0-45.0) | <0.001 |
| | Sedentary screen (diary) | Sedentary screen | 91 | **0.78 [0.678, 0.846]** | 93.0 (43.4-134.6) | 90.0 (54.6-154.3) | 0.002 |
| | Total sleep (diary+ACC) | Total sleep | 92 | **0.48 [0.311, 0.627]** | 602.3 (586.4-632.8) | 600.0 (540.0-660.0) | 0.415 |
| | Night sleep (diary+ACC) | Night sleep | 92 | **0.56 [0.403, 0.687]** | 592.3 (569.9-611.9) | 540.0 (540.0-660.0) | 0.027 |
| | Day sleep (diary+ACC) | Day sleep | 92 | **0.74 [0.635, 0.823]** | 5.4 (0.0-17.1) | 0.0 (0.0-30.0) | <0.001 |

Supplementary Table S15. Continued.
Closed-Paper

| Period | Criterion measure | MBQ-C item | n | ρ [95% CI] | Criterion Med (Q1-Q3) | MBQ-C Med (Q1-Q3) | P |
|---|---|---|---|---|---|---|---|
| Weekday | Total PA (ACC) | Active play | 44 | **0.32 [0.029, 0.566]** | 410.5 (360.9-444.3) | 150.0 (90.0-210.0) | NA |
| | MVPA (ACC) | Energetic play | 44 | 0.07 [-0.233, 0.358] | 81.9 (61.2-104.8) | 45.0 (22.5-75.0) | NA |
| | Total screen time (diary) | Total screen time | 44 | **0.76 [0.591, 0.859]** | 80.6 (29.2-96.0) | 75.0 (52.5-150.0) | <0.001 |
| | Passive screen (diary) | Passive screen | 44 | **0.79 [0.649, 0.882]** | 58.9 (22.1-86.2) | 75.0 (45.0-82.5) | 0.004 |
| | Interactive screen (diary) | Interactive screen | 44 | **0.62 [0.395, 0.774]** | 0.0 (0.0-22.9) | 7.5 (0.0-45.0) | 0.006 |
| | Sedentary screen (diary) | Sedentary screen | 44 | **0.78 [0.621, 0.872]** | 70.1 (26.8-86.2) | 67.5 (45.0-136.9) | 0.004 |
| | Night sleep (diary+ACC) | Night sleep | 44 | **0.64 [0.425, 0.788]** | 595.3 (566.7-617.2) | 540.0 (540.0-660.0) | 0.513 |
| | Day sleep (diary+ACC) | Day sleep | 44 | **0.59 [0.36, 0.757]** | 0.0 (0.0-8.5) | 15.0 (0.0-30.0) | 0.001 |
| Weekend | Total PA (ACC) | Active play | 44 | 0.24 [-0.064, 0.499] | 406.6 (348.1-459.9) | 90.0 (90.0-165.0) | NA |
| | MVPA (ACC) | Energetic play | 44 | 0.05 [-0.25, 0.343] | 90.5 (56.7-123.2) | 45.0 (22.5-75.0) | NA |
| | Total screen time (diary) | Total screen time | 44 | **0.82 [0.687, 0.897]** | 101.2 (60.0-202.5) | 127.5 (75.0-202.5) | 0.199 |
| | Passive screen (diary) | Passive screen | 44 | **0.77 [0.609, 0.867]** | 90.0 (45.0-165.0) | 105.0 (45.0-150.0) | 0.429 |
| | Interactive screen (diary) | Interactive screen | 44 | **0.69 [0.494, 0.819]** | 0.0 (0.0-41.2) | 22.5 (0.0-45.0) | 0.081 |
| | Sedentary screen (diary) | Sedentary screen | 44 | **0.83 [0.708, 0.904]** | 101.2 (60.0-176.2) | 112.5 (52.5-168.8) | 0.915 |
| | Night sleep (diary+ACC) | Night sleep | 44 | 0.21 [-0.097, 0.474] | 597.6 (567.0-628.1) | 540.0 (540.0-660.0) | 0.385 |
| | Day sleep (diary+ACC) | Day sleep | 44 | **0.52 [0.257, 0.704]** | 0.0 (0.0-30.0) | 15.0 (0.0-30.0) | 0.037 |
| Weekly | Total PA (ACC) | Active play | 44 | **0.30 [0.005, 0.549]** | 399.8 (375.0-437.1) | 132.9 (90.0-197.1) | NA |
| | MVPA (ACC) | Energetic play | 44 | 0.04 [-0.262, 0.331] | 82.3 (67.0-101.8) | 45.0 (22.5-75.0) | NA |
| | Total screen time (diary) | Total screen time | 44 | **0.80 [0.667, 0.889]** | 91.3 (42.9-132.3) | 98.0 (67.8-150.0) | 0.001 |
| | Passive screen (diary) | Passive screen | 44 | **0.82 [0.698, 0.901]** | 63.2 (37.4-120.4) | 79.3 (45.0-117.9) | 0.020 |
| | Interactive screen (diary) | Interactive screen | 44 | **0.65 [0.438, 0.794]** | 1.1 (0.0-25.2) | 12.3 (0.0-45.0) | 0.010 |
| | Sedentary screen (diary) | Sedentary screen | 44 | **0.84 [0.732, 0.913]** | 76.0 (42.2-120.2) | 83.0 (53.3-138.2) | 0.028 |
| | Total sleep (diary+ACC) | Total sleep | 44 | **0.47 [0.205, 0.675]** | 601.4 (588.0-634.2) | 630.0 (540.0-660.0) | 0.599 |
| | Night sleep (diary+ACC) | Night sleep | 44 | **0.59 [0.361, 0.757]** | 590.1 (571.1-614.4) | 540.0 (540.0-660.0) | 0.470 |
| | Day sleep (diary+ACC) | Day sleep | 44 | **0.77 [0.618, 0.87]** | 5.4 (0.0-16.5) | 15.0 (0.0-30.0) | 0.001 |

Closed-Electronic

| Period | Criterion measure | MBQ-C item | n | ρ [95% CI] | Criterion Med (Q1-Q3) | MBQ-C Med (Q1-Q3) | P |
|---|---|---|---|---|---|---|---|
| Weekday | Total PA (ACC) | Active play | 48 | 0.23 [-0.058, 0.482] | 403.8 (360.8-439.1) | 150.0 (90.0-217.5) | NA |
| | MVPA (ACC) | Energetic play | 48 | **0.32 [0.044, 0.557]** | 70.3 (55.1-97.8) | 45.0 (7.5-75.0) | NA |
| | Total screen time (diary) | Total screen time | 48 | **0.75 [0.589, 0.851]** | 94.9 (45.0-146.4) | 123.8 (50.6-157.5) | <0.001 |
| | Passive screen (diary) | Passive screen | 47 | **0.62 [0.404, 0.77]** | 78.0 (36.8-144.0) | 105.0 (45.0-150.0) | 0.100 |
| | Interactive screen (diary) | Interactive screen | 47 | **0.47 [0.211, 0.667]** | 0.0 (0.0-11.2) | 7.5 (0.0-45.0) | 0.001 |
| | Sedentary screen (diary) | Sedentary screen | 47 | **0.71 [0.527, 0.826]** | 86.2 (45.0-128.6) | 97.5 (45.0-146.2) | 0.006 |
| | Night sleep (diary+ACC) | Night sleep | 48 | **0.57 [0.345, 0.737]** | 599.1 (558.4-610.4) | 540.0 (540.0-660.0) | 0.030 |
| | Day sleep (diary+ACC) | Day sleep | 48 | **0.66 [0.456, 0.792]** | 0.0 (0.0-6.0) | 0.0 (0.0-30.0) | 0.027 |
| Weekend | Total PA (ACC) | Active play | 48 | 0.24 [-0.046, 0.492] | 377.2 (327.4-430.8) | 90.0 (45.0-150.0) | NA |
| | MVPA (ACC) | Energetic play | 48 | 0.07 [-0.216, 0.35] | 80.2 (52.3-99.9) | 22.5 (7.5-75.0) | NA |
| | Total screen time (diary) | Total screen time | 48 | **0.65 [0.452, 0.79]** | 137.5 (80.6-221.2) | 138.8 (80.6-240.0) | 0.841 |
| | Passive screen (diary) | Passive screen | 48 | **0.60 [0.384, 0.757]** | 112.5 (73.1-175.0) | 105.0 (75.0-150.0) | 0.548 |
| | Interactive screen (diary) | Interactive screen | 48 | **0.64 [0.437, 0.783]** | 2.5 (0.0-31.9) | 7.5 (0.0-45.0) | 0.103 |
| | Sedentary screen (diary) | Sedentary screen | 48 | **0.65 [0.447, 0.787]** | 125.0 (69.4-207.5) | 123.8 (75.0-211.9) | 0.764 |
| | Night sleep (diary+ACC) | Night sleep | 48 | 0.17 [-0.117, 0.435] | 595.7 (576.5-624.2) | 540.0 (540.0-660.0) | 0.012 |
| | Day sleep (diary+ACC) | Day sleep | 48 | **0.53 [0.29, 0.708]** | 0.0 (0.0-28.0) | 0.0 (0.0-30.0) | 0.909 |
| Weekly | Total PA (ACC) | Active play | 48 | 0.26 [-0.026, 0.507] | 390.1 (346.5-432.8) | 141.4 (68.6-197.1) | NA |
| | MVPA (ACC) | Energetic play | 48 | 0.17 [-0.121, 0.432] | 70.3 (61.1-91.5) | 38.6 (16.6-68.6) | NA |
| | Total screen time (diary) | Total screen time | 48 | **0.72 [0.546, 0.833]** | 106.1 (56.8-169.8) | 135.0 (63.8-178.7) | 0.006 |
| | Passive screen (diary) | Passive screen | 47 | **0.67 [0.477, 0.804]** | 96.1 (44.2-144.3) | 105.0 (53.6-150.0) | 0.158 |
| | Interactive screen (diary) | Interactive screen | 47 | **0.64 [0.425, 0.78]** | 4.3 (0.0-18.2) | 7.5 (0.0-45.0) | 0.006 |
| | Sedentary screen (diary) | Sedentary screen | 47 | **0.69 [0.504, 0.816]** | 102.9 (51.8-147.1) | 114.6 (58.9-168.2) | 0.015 |
| | Total sleep (diary+ACC) | Total sleep | 48 | **0.50 [0.255, 0.689]** | 603.4 (585.9-623.8) | 570.0 (540.0-660.0) | 0.098 |
| | Night sleep (diary+ACC) | Night sleep | 48 | **0.53 [0.289, 0.707]** | 594.1 (566.6-611.4) | 540.0 (540.0-660.0) | 0.020 |
| | Day sleep (diary+ACC) | Day sleep | 48 | **0.71 [0.538, 0.829]** | 4.3 (0.0-17.1) | 0.0 (0.0-30.0) | 0.218 |

Abbreviations: ACC, accelerometer; CI, confidence interval; MBQ-C, Movement Behaviour Questionnaire-Child; Q1, first quartile; Q3, third quartile. Values are in min/day. Weekly values were calculated as (weekday×5+weekend/holiday×2)/7. Bold ρ values indicate 95% CIs excluding zero. P values are from Wilcoxon signed-rank tests; NA = not applicable for active and energetic play because questionnaire play time and accelerometer-derived activity intensity are not equivalent measures.

Supplementary Table S15. Continued.

**Part B.** Test-retest reliability (ICC), by response format and mode

| Response format-mode | MBQ-C item | n | ICC [95% CI] | Day 7 Med (Q1-Q3) | Day 10 Med (Q1-Q3) |
|---|---|---|---|---|---|
| Open-Overall | Active play | 59 | **0.79 [0.67, 0.87]** | 171.4 (105.0-229.3) | 145.7 (86.4-235.7) |
| | Energetic play | 59 | **0.66 [0.49, 0.78]** | 57.1 (29.3-81.4) | 55.7 (25.7-100.7) |
| | Total screen time | 59 | **0.79 [0.67, 0.87]** | 137.1 (73.6-207.9) | 141.4 (68.6-218.6) |
| | Passive screen | 59 | **0.85 [0.75, 0.91]** | 115.7 (61.8-154.3) | 98.6 (60.0-152.1) |
| | Interactive screen | 59 | **0.59 [0.39, 0.74]** | 8.6 (0.0-36.4) | 15.7 (0.0-49.3) |
| | Sedentary screen | 59 | **0.80 [0.69, 0.88]** | 122.1 (62.9-182.9) | 111.4 (68.6-191.8) |
| | Day sleep | 59 | **0.81 [0.70, 0.88]** | 0.0 (0.0-35.0) | 0.0 (0.0-45.0) |
| | Night sleep | 59 | **0.70 [0.55, 0.81]** | 570.0 (540.0-600.0) | 570.0 (540.0-600.0) |
| | Total sleep | 59 | **0.68 [0.51, 0.79]** | 600.0 (560.0-630.0) | 600.0 (570.0-630.0) |
| | Sleep routine | 59 | **0.81 [0.70, 0.88]** | 6.0 (4.0-7.0) | 6.0 (4.0-7.0) |
| Open-Paper | Active play | 29 | **0.76 [0.54, 0.88]** | 158.6 (77.1-205.7) | 128.6 (62.9-205.7) |
| | Energetic play | 29 | 0.47 [0.12, 0.71] | 51.4 (24.3-68.6) | 38.6 (16.9-77.1) |
| | Total screen time | 29 | **0.82 [0.65, 0.91]** | 169.3 (137.1-214.3) | 160.7 (118.6-218.6) |
| | Passive screen | 29 | **0.85 [0.69, 0.93]** | 137.1 (115.7-154.3) | 137.1 (77.1-154.3) |
| | Interactive screen | 29 | **0.82 [0.65, 0.91]** | 21.4 (0.0-38.6) | 30.0 (10.7-60.0) |
| | Sedentary screen | 29 | **0.81 [0.64, 0.91]** | 145.7 (122.1-199.3) | 152.1 (95.7-195.0) |
| | Day sleep | 29 | **0.74 [0.51, 0.87]** | 0.0 (0.0-60.0) | 0.0 (0.0-60.0) |
| | Night sleep | 29 | **0.85 [0.71, 0.93]** | 570.0 (540.0-600.0) | 600.0 (540.0-600.0) |
| | Total sleep | 29 | **0.74 [0.52, 0.87]** | 600.0 (570.0-650.0) | 600.0 (570.0-630.0) |
| | Sleep routine | 29 | **0.77 [0.57, 0.89]** | 6.0 (3.0-7.0) | 6.0 (3.0-7.0) |
| Open-Electronic | Active play | 30 | **0.81 [0.64, 0.91]** | 175.7 (121.1-274.3) | 160.7 (121.1-240.0) |
| | Energetic play | 30 | **0.75 [0.54, 0.87]** | 60.0 (42.9-97.5) | 61.1 (32.1-102.9) |
| | Total screen time | 30 | **0.76 [0.55, 0.88]** | 90.0 (56.8-174.6) | 86.8 (47.1-212.1) |
| | Passive screen | 30 | **0.83 [0.67, 0.91]** | 77.1 (56.8-115.7) | 77.1 (40.7-120.0) |
| | Interactive screen | 30 | 0.42 [0.09, 0.67] | 1.1 (0.0-19.8) | 6.8 (0.0-33.2) |
| | Sedentary screen | 30 | **0.77 [0.57, 0.88]** | 78.6 (43.9-125.9) | 78.2 (39.5-178.9) |
| | Day sleep | 30 | **0.90 [0.80, 0.95]** | 0.0 (0.0-26.2) | 0.0 (0.0-30.0) |
| | Night sleep | 30 | **0.58 [0.28, 0.78]** | 570.0 (540.0-600.0) | 570.0 (540.0-600.0) |
| | Total sleep | 30 | **0.63 [0.35, 0.80]** | 600.0 (547.5-630.0) | 600.0 (570.0-630.0) |
| | Sleep routine | 30 | **0.87 [0.75, 0.94]** | 7.0 (4.0-7.0) | 6.5 (5.0-7.0) |
| Closed-Overall | Active play | 61 | **0.71 [0.53, 0.82]** | 132.9 (64.3-197.1) | 90.0 (57.9-150.0) |
| | Energetic play | 61 | **0.69 [0.49, 0.82]** | 45.0 (22.5-75.0) | 28.9 (18.2-53.6) |
| | Total screen time | 61 | **0.81 [0.70, 0.88]** | 132.9 (75.0-175.7) | 117.9 (68.6-166.1) |
| | Passive screen | 61 | **0.80 [0.69, 0.88]** | 96.4 (53.6-150.0) | 96.4 (46.1-143.6) |
| | Interactive screen | 61 | **0.85 [0.77, 0.91]** | 16.1 (0.0-45.0) | 11.8 (0.0-45.0) |
| | Sedentary screen | 61 | **0.83 [0.73, 0.89]** | 98.6 (62.1-165.0) | 90.0 (54.6-138.2) |
| | Day sleep | 61 | **0.89 [0.82, 0.93]** | 0.0 (0.0-30.0) | 0.0 (0.0-30.0) |
| | Night sleep | 61 | **0.75 [0.62, 0.84]** | 540.0 (540.0-660.0) | 540.0 (540.0-660.0) |
| | Total sleep | 61 | **0.72 [0.58, 0.82]** | 630.0 (540.0-660.0) | 630.0 (540.0-660.0) |
| | Sleep routine | 61 | **0.83 [0.73, 0.90]** | 4.0 (2.0-4.0) | 3.0 (2.0-4.0) |
| Closed-Paper | Active play | 30 | **0.63 [0.32, 0.81]** | 141.4 (90.0-197.1) | 107.1 (48.2-145.7) |
| | Energetic play | 30 | **0.58 [0.26, 0.78]** | 52.0 (24.1-79.8) | 41.8 (19.3-60.0) |
| | Total screen time | 30 | **0.78 [0.59, 0.89]** | 114.1 (86.8-162.9) | 137.1 (83.6-159.6) |
| | Passive screen | 30 | **0.83 [0.67, 0.92]** | 90.0 (56.8-121.1) | 90.0 (45.3-143.6) |
| | Interactive screen | 30 | **0.83 [0.68, 0.92]** | 25.7 (0.5-58.4) | 22.5 (1.3-45.0) |
| | Sedentary screen | 30 | **0.82 [0.66, 0.91]** | 93.8 (67.8-162.9) | 94.3 (65.4-132.9) |
| | Day sleep | 30 | **0.82 [0.66, 0.91]** | 30.0 (0.0-30.0) | 0.0 (0.0-30.0) |
| | Night sleep | 30 | **0.80 [0.62, 0.90]** | 540.0 (540.0-660.0) | 540.0 (540.0-660.0) |
| | Total sleep | 30 | **0.76 [0.56, 0.88]** | 630.0 (540.0-660.0) | 630.0 (540.0-660.0) |
| | Sleep routine | 30 | **0.86 [0.73, 0.93]** | 4.0 (2.2-4.0) | 3.5 (1.0-4.0) |
| Closed-Electronic | Active play | 31 | **0.79 [0.61, 0.89]** | 132.9 (54.6-195.0) | 90.0 (57.9-162.9) |
| | Energetic play | 31 | **0.80 [0.60, 0.90]** | 32.1 (9.6-63.2) | 22.5 (13.4-38.6) |
| | Total screen time | 31 | **0.85 [0.67, 0.93]** | 141.4 (69.6-181.6) | 112.5 (57.9-166.1) |
| | Passive screen | 31 | **0.78 [0.57, 0.89]** | 105.0 (49.8-162.9) | 105.0 (49.8-136.1) |
| | Interactive screen | 31 | **0.88 [0.76, 0.94]** | 11.8 (0.0-38.6) | 7.5 (0.0-20.4) |
| | Sedentary screen | 31 | **0.84 [0.68, 0.92]** | 118.9 (58.9-167.1) | 83.6 (46.6-158.0) |
| | Day sleep | 31 | **0.92 [0.84, 0.96]** | 0.0 (0.0-30.0) | 0.0 (0.0-30.0) |
| | Night sleep | 31 | **0.68 [0.44, 0.83]** | 540.0 (540.0-660.0) | 540.0 (540.0-660.0) |
| | Total sleep | 31 | **0.67 [0.42, 0.83]** | 570.0 (540.0-660.0) | 630.0 (540.0-660.0) |
| | Sleep routine | 31 | **0.80 [0.62, 0.90]** | 3.0 (2.0-4.0) | 3.0 (2.0-4.0) |

ICC, intraclass correlation coefficient; other abbreviations as in Part A. Values in min/day (weighted weekly average). Bold ICC values indicate ICC ≥ 0.5 (single-measurement, absolute-agreement, two-way mixed-effects model).

**Supplementary Figure S16. Bland–Altman analysis for total screen time and total sleep**

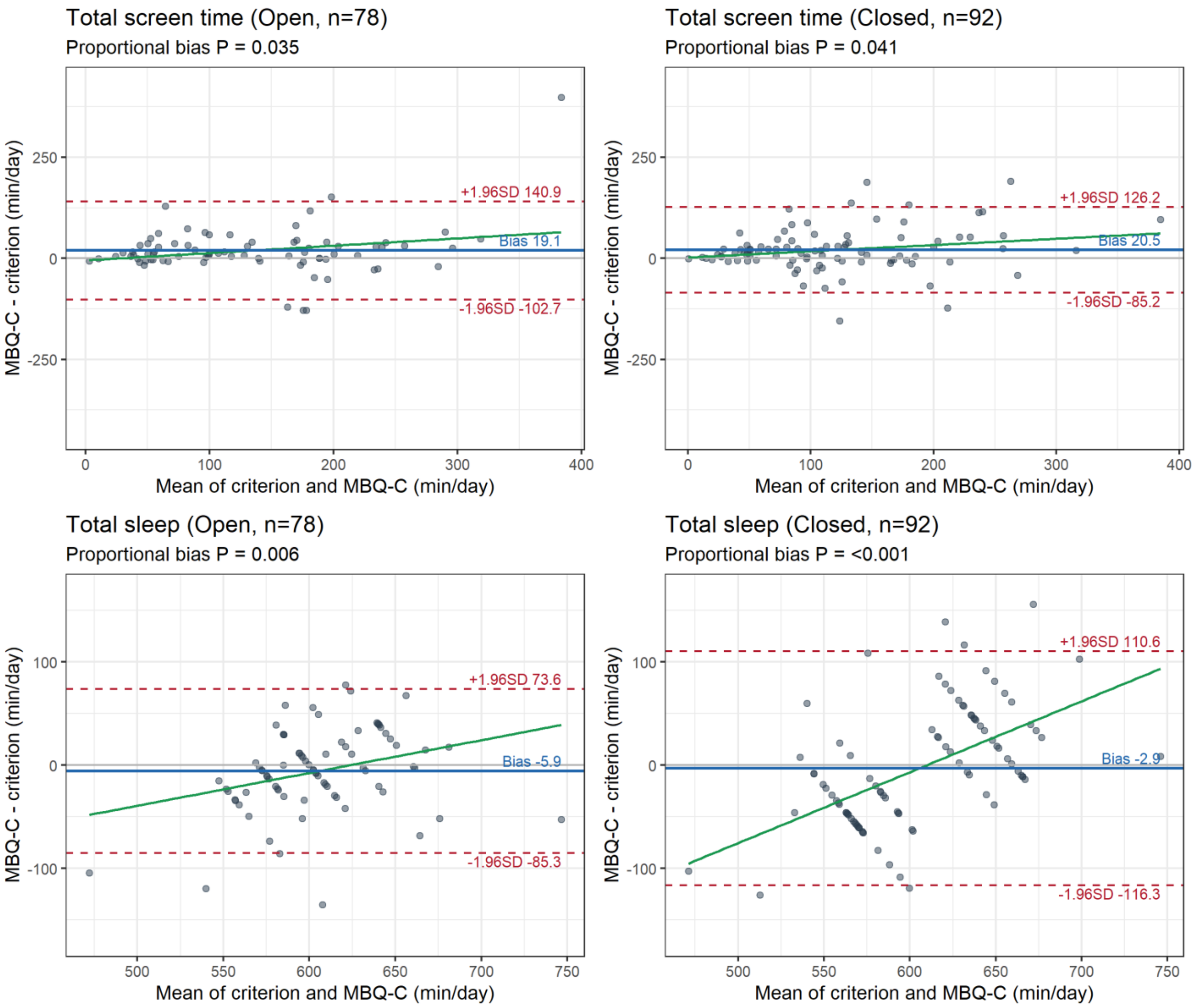

Bland–Altman plots comparing the Japanese version of the Movement Behaviour Questionnaire-Child (MBQ-C) with criterion measures for total screen time and total sleep. The four panels show total screen time and total sleep by response format (open and closed). The x-axis shows the mean of the MBQ-C and criterion values (min/day), and the y-axis shows the difference between them (MBQ-C − criterion, min/day); positive values indicate overestimation by the MBQ-C. The blue solid line represents the mean bias, the red dashed lines represent the 95% limits of agreement, and the green line represents the regression line used to assess proportional bias. The P value for proportional bias is shown below each panel title. The criterion for total screen time was the parent-reported 24-h time-use diary, and the criterion for total sleep was derived by integrating the diary with accelerometer data.

#### Supplementary Table S17. Sensitivity analysis restricted to children aged 3–5 years

##### A. Concurrent validity

| Response format-mode | Criterion measure | MBQ-C item | n | ρ [95% CI] | Criterion Med (Q1-Q3) | MBQ-C Med (Q1-Q3) | P |
|---|---|---|---|---|---|---|---|
| Open-Overall | Total PA (ACC) | Active play | 69 | 0.20 [-0.039, 0.417] | 391.2 (355.6-433.0) | 162.9 (98.6-205.7) | NA |
| | MVPA (ACC) | Energetic play | 69 | 0.20 [-0.039, 0.416] | 66.1 (54.6-83.8) | 51.4 (27.9-77.1) | NA |
| | Total screen time (diary) | Total screen time | 69 | **0.83 [0.744, 0.894]** | 100.4 (49.3-181.8) | 118.6 (63.6-188.6) | <0.001 |
| | Total sleep (diary+ACC) | Total sleep | 69 | **0.57 [0.382, 0.709]** | 607.4 (582.0-629.0) | 600.0 (570.0-645.0) | 0.788 |
| Open-Paper | Total PA (ACC) | Active play | 43 | 0.26 [-0.044, 0.52] | 391.2 (360.9-429.0) | 141.4 (79.3-197.1) | NA |
| | MVPA (ACC) | Energetic play | 43 | 0.03 [-0.272, 0.328] | 64.1 (56.3-78.0) | 34.3 (23.6-64.3) | NA |
| | Total screen time (diary) | Total screen time | 43 | **0.75 [0.583, 0.858]** | 114.1 (53.6-177.2) | 128.6 (70.7-188.6) | 0.015 |
| | Total sleep (diary+ACC) | Total sleep | 43 | **0.61 [0.383, 0.771]** | 599.7 (574.9-626.1) | 600.0 (575.0-655.0) | 0.948 |
| Open-Electronic | Total PA (ACC) | Active play | 26 | 0.17 [-0.231, 0.524] | 389.9 (346.5-438.6) | 169.3 (121.1-272.1) | NA |
| | MVPA (ACC) | Energetic play | 26 | 0.37 [-0.023, 0.661] | 69.9 (52.7-100.0) | 68.6 (43.4-97.5) | NA |
| | Total screen time (diary) | Total screen time | 26 | **0.91 [0.803, 0.958]** | 86.7 (37.5-192.3) | 98.6 (60.9-190.7) | <0.001 |
| | Total sleep (diary+ACC) | Total sleep | 26 | **0.51 [0.151, 0.749]** | 611.7 (590.9-629.9) | 600.0 (570.0-630.0) | 0.515 |
| Closed-Overall | Total PA (ACC) | Active play | 83 | **0.27 [0.061, 0.462]** | 398.9 (355.3-433.3) | 132.9 (90.0-197.1) | NA |
| | MVPA (ACC) | Energetic play | 83 | 0.14 [-0.08, 0.343] | 74.9 (61.0-95.0) | 38.6 (22.5-75.0) | NA |
| | Total screen time (diary) | Total screen time | 83 | **0.80 [0.711, 0.868]** | 98.0 (43.9-151.3) | 112.5 (61.6-160.7) | <0.001 |
| | Total sleep (diary+ACC) | Total sleep | 83 | **0.50 [0.314, 0.643]** | 605.7 (587.8-636.0) | 630.0 (540.0-660.0) | 0.410 |
| Closed-Paper | Total PA (ACC) | Active play | 40 | 0.21 [-0.108, 0.49] | 399.8 (377.2-434.8) | 132.9 (90.0-197.1) | NA |
| | MVPA (ACC) | Energetic play | 40 | 0.00 [-0.308, 0.315] | 82.1 (66.8-101.8) | 45.0 (22.5-75.0) | NA |
| | Total screen time (diary) | Total screen time | 40 | **0.83 [0.702, 0.908]** | 86.5 (42.3-145.0) | 98.0 (64.3-150.0) | 0.001 |
| | Total sleep (diary+ACC) | Total sleep | 40 | **0.45 [0.157, 0.665]** | 605.6 (588.0-636.5) | 630.0 (570.0-660.0) | 0.468 |
| Closed-Electronic | Total PA (ACC) | Active play | 43 | **0.39 [0.1, 0.617]** | 393.2 (346.4-431.4) | 150.0 (71.8-197.1) | NA |
| | MVPA (ACC) | Energetic play | 43 | 0.23 [-0.077, 0.495] | 69.6 (59.5-88.1) | 38.6 (20.4-75.0) | NA |
| | Total screen time (diary) | Total screen time | 43 | **0.74 [0.561, 0.849]** | 101.6 (52.9-158.6) | 127.5 (61.1-170.9) | 0.008 |
| | Total sleep (diary+ACC) | Total sleep | 43 | **0.55 [0.304, 0.732]** | 605.7 (589.5-631.9) | 570.0 (540.0-660.0) | 0.057 |

Supplementary Table S17. Continued.
**B. Test-retest reliability**

| Response format-mode | MBQ-C item | n | ICC [95% CI] | Day 7 Med (Q1-Q3) | Day 10 Med (Q1-Q3) |
|---|---|---|---|---|---|
| Open-Overall | Active play | 52 | **0.79 [0.657, 0.872]** | 175.7 (114.6-237.9) | 162.9 (93.6-240.0) |
| | Energetic play | 52 | **0.65 [0.462, 0.785]** | 58.6 (29.6-81.4) | 60.0 (28.9-102.9) |
| | Total screen time | 52 | **0.76 [0.615, 0.854]** | 134.3 (70.0-196.1) | 130.7 (68.6-212.1) |
| | Total sleep | 52 | **0.61 [0.405, 0.756]** | 600.0 (570.0-646.3) | 600.0 (570.0-632.5) |
| | Sleep routine | 52 | **0.77 [0.624, 0.858]** | 6.5 (4.0-7.0) | 6.0 (4.8-7.0) |
| Open-Paper | Active play | 25 | **0.75 [0.506, 0.882]** | 171.4 (77.1-205.7) | 128.6 (62.9-231.4) |
| | Energetic play | 25 | 0.46 [0.081, 0.72] | 51.4 (24.3-68.6) | 40.0 (17.1-87.1) |
| | Total screen time | 25 | **0.81 [0.625, 0.913]** | 169.3 (131.4-210.0) | 154.3 (115.7-210.0) |
| | Total sleep | 25 | **0.72 [0.463, 0.867]** | 600.0 (570.0-660.0) | 600.0 (570.0-630.0) |
| | Sleep routine | 25 | **0.74 [0.483, 0.874]** | 6.0 (3.0-7.0) | 6.0 (3.0-7.0) |
| Open-Electronic | Active play | 27 | **0.81 [0.638, 0.911]** | 180.0 (143.6-274.3) | 162.9 (126.4-252.9) |
| | Energetic play | 27 | **0.75 [0.522, 0.877]** | 60.0 (43.9-92.1) | 62.1 (38.6-102.9) |
| | Total screen time | 27 | **0.73 [0.493, 0.867]** | 90.0 (56.4-165.0) | 96.4 (47.1-201.4) |
| | Total sleep | 27 | **0.51 [0.166, 0.741]** | 600.0 (570.0-630.0) | 600.0 (585.0-640.0) |
| | Sleep routine | 27 | **0.81 [0.622, 0.908]** | 7.0 (4.5-7.0) | 7.0 (5.0-7.0) |
| Closed-Overall | Active play | 53 | **0.68 [0.487, 0.81]** | 132.9 (68.6-197.1) | 107.1 (57.9-150.0) |
| | Energetic play | 53 | **0.71 [0.506, 0.828]** | 38.6 (22.5-75.0) | 28.9 (16.1-53.6) |
| | Total screen time | 53 | **0.82 [0.704, 0.89]** | 132.9 (75.0-166.1) | 117.9 (64.3-162.9) |
| | Total sleep | 53 | **0.71 [0.552, 0.824]** | 630.0 (540.0-660.0) | 630.0 (540.0-660.0) |
| | Sleep routine | 53 | **0.86 [0.777, 0.92]** | 4.0 (2.0-4.0) | 4.0 (2.0-4.0) |
| Closed-Paper | Active play | 26 | **0.60 [0.264, 0.799]** | 141.4 (94.3-197.1) | 107.1 (59.5-145.7) |
| | Energetic play | 26 | **0.60 [0.271, 0.801]** | 57.9 (24.1-79.8) | 41.8 (18.2-64.8) |
| | Total screen time | 26 | **0.78 [0.574, 0.897]** | 124.3 (86.8-162.9) | 137.1 (85.2-159.6) |
| | Total sleep | 26 | **0.73 [0.486, 0.869]** | 630.0 (547.5-660.0) | 630.0 (570.0-660.0) |
| | Sleep routine | 26 | **0.83 [0.659, 0.923]** | 4.0 (3.0-4.0) | 4.0 (1.2-4.0) |
| Closed-Electronic | Active play | 27 | **0.77 [0.566, 0.89]** | 132.9 (66.4-190.7) | 90.0 (66.4-162.9) |
| | Energetic play | 27 | **0.80 [0.608, 0.907]** | 32.1 (11.8-56.8) | 22.5 (13.4-38.6) |
| | Total screen time | 27 | **0.87 [0.701, 0.94]** | 137.1 (60.0-170.9) | 112.5 (50.4-158.0) |
| | Total sleep | 27 | **0.70 [0.436, 0.85]** | 630.0 (555.0-660.0) | 630.0 (540.0-660.0) |
| | Sleep routine | 27 | **0.91 [0.816, 0.959]** | 4.0 (2.0-4.0) | 3.0 (2.0-4.0) |

*Restricted to children aged 3-5 years (<72 months). Abbreviations and formatting as in Table 2 and Table 3. Bold ρ values indicate a 95% CI that does not include zero. Bold ICC values indicate ICC ≥ 0.5.*
*P values are from Wilcoxon signed-rank tests; NA = not applicable for active and energetic play because questionnaire play time and accelerometer-derived activity intensity are not equivalent measures.*

#### Supplementary Table S18. Sensitivity analysis varying the wear-time criterion

| Response format-mode | MBQ-C item | n 360 min/d | ρ [95% CI] 360 min/d | n 600 min/d | ρ [95% CI] 600 min/d | n 960 min/d | ρ [95% CI] 960 min/d |
|---|---|---|---|---|---|---|---|
| Open-Overall | Active play | 79 | 0.16 [-0.059, 0.372] | 78 | 0.15 [-0.079, 0.357] | 66 | 0.23 [-0.01, 0.449] |
| | Energetic play | 79 | **0.25 [0.025, 0.442]** | 78 | **0.22 [0.001, 0.425]** | 66 | **0.24 [0.001, 0.458]** |
| | Total screen time | 79 | **0.84 [0.764, 0.897]** | 78 | **0.85 [0.771, 0.901]** | 66 | **0.83 [0.742, 0.895]** |
| | Total sleep | 79 | **0.59 [0.421, 0.716]** | 78 | **0.58 [0.404, 0.707]** | 66 | **0.60 [0.424, 0.738]** |
| Open-Paper | Active play | 50 | 0.27 [-0.008, 0.511] | 49 | 0.27 [-0.012, 0.512] | 43 | 0.30 [-0.002, 0.549] |
| | Energetic play | 50 | 0.17 [-0.114, 0.428] | 49 | 0.17 [-0.115, 0.432] | 43 | 0.21 [-0.099, 0.478] |
| | Total screen time | 50 | **0.76 [0.608, 0.856]** | 49 | **0.77 [0.626, 0.865]** | 43 | **0.74 [0.566, 0.851]** |
| | Total sleep | 50 | **0.59 [0.37, 0.745]** | 49 | **0.58 [0.352, 0.738]** | 43 | **0.59 [0.355, 0.758]** |
| Open-Electronic | Active play | 29 | 0.03 [-0.342, 0.39] | 29 | 0.01 [-0.358, 0.375] | 23 | 0.11 [-0.319, 0.498] |
| | Energetic play | 29 | 0.27 [-0.112, 0.576] | 29 | 0.24 [-0.142, 0.555] | 23 | 0.22 [-0.212, 0.579] |
| | Total screen time | 29 | **0.91 [0.819, 0.958]** | 29 | **0.91 [0.819, 0.958]** | 23 | **0.94 [0.864, 0.975]** |
| | Total sleep | 29 | **0.63 [0.338, 0.808]** | 29 | **0.60 [0.297, 0.791]** | 23 | **0.66 [0.336, 0.841]** |
| Closed-Overall | Active play | 94 | **0.23 [0.026, 0.411]** | 92 | **0.26 [0.056, 0.44]** | 79 | 0.19 [-0.033, 0.394] |
| | Energetic play | 94 | 0.12 [-0.085, 0.315] | 92 | 0.12 [-0.087, 0.317] | 79 | 0.04 [-0.186, 0.255] |
| | Total screen time | 94 | **0.79 [0.702, 0.857]** | 92 | **0.78 [0.689, 0.851]** | 79 | **0.75 [0.64, 0.836]** |
| | Total sleep | 94 | **0.47 [0.302, 0.618]** | 92 | **0.48 [0.311, 0.627]** | 79 | **0.48 [0.295, 0.638]** |
| Closed-Paper | Active play | 45 | 0.26 [-0.042, 0.51] | 44 | **0.30 [0.005, 0.549]** | 38 | 0.22 [-0.11, 0.502] |
| | Energetic play | 45 | 0.02 [-0.278, 0.309] | 44 | 0.04 [-0.262, 0.331] | 38 | -0.04 [-0.358, 0.28] |
| | Total screen time | 45 | **0.81 [0.682, 0.893]** | 44 | **0.80 [0.667, 0.889]** | 38 | **0.82 [0.672, 0.901]** |
| | Total sleep | 45 | **0.45 [0.18, 0.657]** | 44 | **0.47 [0.205, 0.675]** | 38 | **0.49 [0.204, 0.701]** |
| Closed-Electronic | Active play | 49 | 0.24 [-0.039, 0.493] | 48 | 0.26 [-0.026, 0.507] | 41 | 0.17 [-0.148, 0.451] |
| | Energetic play | 49 | 0.18 [-0.108, 0.438] | 48 | 0.17 [-0.121, 0.432] | 41 | 0.09 [-0.226, 0.385] |
| | Total screen time | 49 | **0.73 [0.573, 0.842]** | 48 | **0.72 [0.546, 0.833]** | 41 | **0.66 [0.44, 0.804]** |
| | Total sleep | 49 | **0.51 [0.264, 0.69]** | 48 | **0.50 [0.255, 0.689]** | 41 | **0.47 [0.183, 0.676]** |

Abbreviations: CI, confidence interval; MBQ-C, Movement Behaviour Questionnaire-Child; MVPA, moderate-to-vigorous physical activity; ρ, Spearman's rank correlation coefficient.

Values are weighted weekly averages in min/day, calculated as (weekday×5+weekend/holiday×2)/7. The main analysis used the 600 min/day criterion. The 360 and 600 min/day criteria refer to wear time within the 07:00–21:00 waking window, corresponding to 6 h and 10 h, respectively. The 960 min/day criterion refers to all-day wear time, including sleep, corresponding to 16 h in 24 h.

The number of included participants may be identical across wear-time criteria, while ρ may differ. This is because the wear-time criterion determines which days are treated as valid for each child; accelerometer-derived criterion values are calculated from each child's valid-day set. Therefore, ρ may change even when the number of participants is unchanged. n is shown for each wear-time criterion. Bold ρ values indicate 95% CIs that do not include zero.

#### Supplementary Table S19. Sensitivity analysis by diary-recording quality

| Response format–mode | MBQ-C item | Main analysis | (1) Usual days | (2) Accurate diary days | (3) Usual & accurate days |
|---|---|---|---|---|---|
| Open–Overall | Active play | 0.15 [-0.079, 0.357] (n=78) | 0.21 [-0.012, 0.414] (n=78) | 0.21 [-0.012, 0.417] (n=77) | **0.25 [0.026, 0.448]** (n=77) |
| | Energetic play | **0.22 [0.001, 0.425]** (n=78) | 0.24 [-0.004, 0.451] (n=67) | 0.19 [-0.038, 0.406] (n=73) | **0.30 [0.050, 0.508]** (n=62) |
| | Total screen time | **0.85 [0.771, 0.901]** (n=78) | **0.85 [0.774, 0.907]** (n=69) | **0.83 [0.735, 0.887]** (n=74) | **0.85 [0.758, 0.904]** (n=64) |
| | Total sleep | **0.58 [0.404, 0.707]** (n=78) | **0.60 [0.426, 0.734]** (n=69) | **0.55 [0.365, 0.690]** (n=74) | **0.63 [0.460, 0.761]** (n=64) |
| Closed–Overall | Active play | **0.26 [0.056, 0.440]** (n=92) | **0.23 [0.031, 0.419]** (n=92) | **0.26 [0.058, 0.443]** (n=91) | **0.25 [0.051, 0.437]** (n=91) |
| | Energetic play | 0.12 [-0.087, 0.317] (n=92) | **0.23 [0.010, 0.421]** (n=83) | 0.13 [-0.078, 0.336] (n=87) | **0.25 [0.025, 0.447]** (n=77) |
| | Total screen time | **0.78 [0.689, 0.851]** (n=92) | **0.77** [0.669, 0.847] (n=84) | **0.76 [0.655, 0.837]** (n=87) | **0.79 [0.682, 0.859]** (n=77) |
| | Total sleep | **0.48 [0.311, 0.627]** (n=92) | **0.51 [0.327, 0.650]** (n=84) | **0.47 [0.283, 0.616]** (n=87) | **0.53 [0.350, 0.676]** (n=77) |

Abbreviations: CI, confidence interval; MBQ-C, Movement Behaviour Questionnaire–Child; ρ, Spearman's rank correlation coefficient.

Values are Spearman ρ [95% CI] between MBQ-C estimates and accelerometer/diary criterion measures, with n shown for each analysis. Analyses were restricted to the 170 children in the main analytic sample who met the accelerometer wear-time criterion of ≥600 min/day. For each sensitivity analysis, weighted weekly criterion means were recomputed using only the specified subset of diary days; all other procedures were identical to the main analysis. (1) Usual days: days on which parents reported that their child had spent the day in the usual way for that day of the week. (2) Accurate diary days: days rated by parents as accurately recorded, defined as the two most accurate of four response categories. (3) Usual & accurate days: days meeting both criteria. Overall combines paper and electronic modes. Bold ρ values indicate 95% CIs that do not include 0. The n may be smaller than in the main analysis because a weighted weekly mean required at least one weekday and one weekend/holiday within the specified day subset.

The Main analysis column corresponds to the primary concurrent-validity results shown in Table 2. Because criterion means were recomputed using restricted diary-day subsets, ρ can differ from the main analysis even when n is unchanged.
